# A multimodal foundation model for emergency head CT interpretation

**DOI:** 10.64898/2026.07.07.26357429

**Authors:** Jingyuan Zheng, Yifei Chen, Beining Wu, Yuanhan Wang, Mingxuan Liu, Lu Li, Shuo Jiang, Weihong Chen, Liaoman Xu, Yueyi Wu, Chang Liu, Lulu Guo, Xuguang Bai, Zihan Li, Hongjia Yang, Feiwei Qin, Jingzhe Liu, Haibo Qu, Qiang Liao, Gang Zhao, Keqin Pan, Jun Guo, Lizhou Chen, Ying Zhou, Huaiqiang Sun, Qiyuan Tian

## Abstract

Non-contrast head CT is the first-line imaging modality for acute neurological emergencies, with demand rising worldwide. However, existing foundation models for head CT interpretation are ill-suited for emergency use because they target general or chronic-disease assessment and optimize reports for lexical overlap rather than the risk-relevant findings central to emergency triage. Here we present CHIEF, a Chinese-language Head CT Interpretation Emergency Foundation model, pretrained on emergency head CT volumes and paired reports with contrastive, generative, and geometry-regularization objectives. Trained and evaluated on 16,563 examinations from seven hospitals, CHIEF achieved an AUROC of 0.9646 for emergency triage and drafted triage-oriented radiology reports, while also supporting image-to-text retrieval for reference-case support and zero-shot abnormality recognition. CHIEF generated reports of substantially higher quality than those from commercial multimodal large language models, which could not be reliably distinguished from human-written ones by radiologists in a blinded *Turing* test. Overall, CHIEF provides a generalizable foundation for emergency head CT interpretation and radiologist-in-the-loop clinical decision support.

## Introduction

Non-contrast head computed tomography (CT) is the first-line imaging modality for evaluating acute neurological emergencies^1^. Acquired within minutes, it enables rapid assessment of a broad spectrum of life-threatening conditions, such as traumatic brain injury, intracranial hemorrhage, and skull fractures^2,3^, enabling timely diagnosis and guiding clinical decision-making^4,5^. Emergency head CT utilization has risen substantially worldwide, driven by population aging, increasing trauma incidence, and progressively lower imaging thresholds^6^. In the United States, Medicare data show an increase from 13.5 to 24.3 examinations per 100 emergency department encounters over the past decade^7^, with similar trends reported across Europe and Asia. Collectively, these developments underscore the growing and accelerating reliance on head CT in acute neurological care^8,9^.

However, the rising volume compounds the challenge of interpreting inherently three-dimensional CT data, which is time-consuming and cognitively demanding. Radiologists must rapidly scroll through large image stacks and integrate information across contiguous slices to extract clinically relevant findings and communicate critical results to clinical teams without delay^100^. In the emergency setting this must happen within minutes, because every delay in interpretation postpones the treatment decisions that depend on it. This process relies heavily on reader experience and sustained attention, increasing the risk of diagnostic errors and communication delays that may compromise clinical decision-making. These risks are particularly pronounced during night shifts, peak emergency surges, and in non-tertiary hospitals lacking dedicated neuroradiology coverage^9^, thereby requiring commensurate growth in radiologist workforce capacity. Yet demand for expert interpretation has outpaced growth in the radiologist workforce, producing a persistent and widening gap in emergency and after-hours coverage^11,12^.

Artificial intelligence (AI) systems have emerged as a promising approach to address these challenges and have advanced rapidly in recent years. Early approaches demonstrated strong performance, but only on narrowly defined clinical tasks, such as detection and subtyping of intracranial hemorrhage^13^, segmentation of traumatic lesions^14^, prediction of hematoma expansion^15^, and estimation of long-term outcomes^16^. Additionally, they exhibit limited cross-institutional generalizability and require large-scale expert annotations. To overcome these limitations, foundation models have been developed to learn task-agnostic and transferable representations through pretraining on large-scale unannotated datasets. Foundation models pretrained through self-supervised learning, such as self-distillation (e.g., DINO-based FM-HCT^17^) and contrastive learning (e.g., SimCLR-based CT-FM^18^), have demonstrated substantial improvements in cross-institutional generalizability, while those pretrained through vision-language alignment between volumetric head CT data and radiology reports exhibit multi-task capability across disease classification, report generation, and cross-modal image–text retrieval^19,20^.

Existing foundation models remain ill-suited to emergency head CT workflows for several reasons^21–24^. First, they are not designed for emergency triage or the recognition of time-critical findings. For example, BrainGPT was trained predominantly on patients with chronic neurodegenerative disease^20^, whereas Brainfound was developed as a general-purpose system rather than for time-critical emergency care^19^. Second, their generalizability has typically been claimed but evaluated only on single public benchmarks or limited external datasets^25–27^, without systematically assessing domain shifts in imaging equipment, acquisition protocols, patient demographics, and disease prevalence^28–31^. Finally, the generated reports of these models often fail to capture diagnostically relevant and clinically actionable information, as model training relies on conventional text-overlap losses^10,32^. In acute settings, however, clinically relevant imaging features such as lesion extent, spatial relationships, mass effect, and midline shift should be prioritized in the learning objective^33^.

In this study, we propose CHIEF, a Chinese-language Head CT Interpretation Emergency Foundation model designed to address these gaps. CHIEF learns unified vision–language representations from emergency head CT volumes and paired radiology reports, with emergency triage classification as its primary clinical endpoint and complementary downstream tasks. By integrating a three-dimensional visual encoder with bidirectional image–text contrastive alignment, image-conditioned generative modeling, and representation geometry regularization, CHIEF captures cross-slice anatomical structure and report-level clinical semantics. We evaluated CHIEF along four clinically grounded dimensions: (i) triage-level decision support, (ii) triage-oriented report generation, (iii) image-to-text retrieval for reference-case support, and (iv) zero-shot abnormality recognition (Extended Data Fig. 1). In addition, we assessed its generalizability across five external hospital datasets, including county-level hospitals, a pediatric specialty hospital, and geographically distinct general hospitals. Overall, CHIEF provides a foundation for emergency head CT interpretation, enabling radiologist-in-the-loop risk communication and actionable decision support across heterogeneous clinical settings.

## Results

### Overview

CHIEF is a self-supervised multimodal pretraining framework designed to learn unified 3D vision-language representations from emergency head CT volumes and their paired radiology reports (West China Hospital and Mianyang Central Hospital, *n* = 13,269). Tailored to the time-critical demands of acute care, CHIEF captures cross-slice anatomical relationships and clinically significant imaging findings, providing a reusable representation for multiple downstream tasks.

To evaluate CHIEF against comparison models, we constructed a multicenter benchmark covering both in-distribution (ID) and external out-of-distribution (OOD) settings (Fig. 1). We performed internal validation on a held-out test set from West China Hospital and Mianyang Central Hospital (*n* = 971), and external validation across five independent hospitals spanning distinct clinical environments. The external sites included county-level primary hospitals introducing variation in equipment and scanning protocols (Santai County People’s Hospital, OOD-1, *n* = 518; Beichuan Qiang Autonomous County People’s Hospital, OOD-2, *n* = 496). They also included a pediatric specialty hospital reflecting shifts in patient age structure and disease spectrum (West China Second University Hospital, Sichuan University, OOD-4, *n* = 500). The remaining sites were general hospitals in a different geographic region, introducing further institutional and demographic heterogeneity (Tsinghua University Hospital, OOD-3, *n* = 477; The First Hospital of Tsinghua University, OOD-5, *n* = 332). Additionally, we further evaluated zero-shot abnormality detection on the internal matched cohort using report-derived labels, and used CQ500^34^ as an external multi-label classification benchmark under a fine-tuning protocol.

**Fig. 1.**
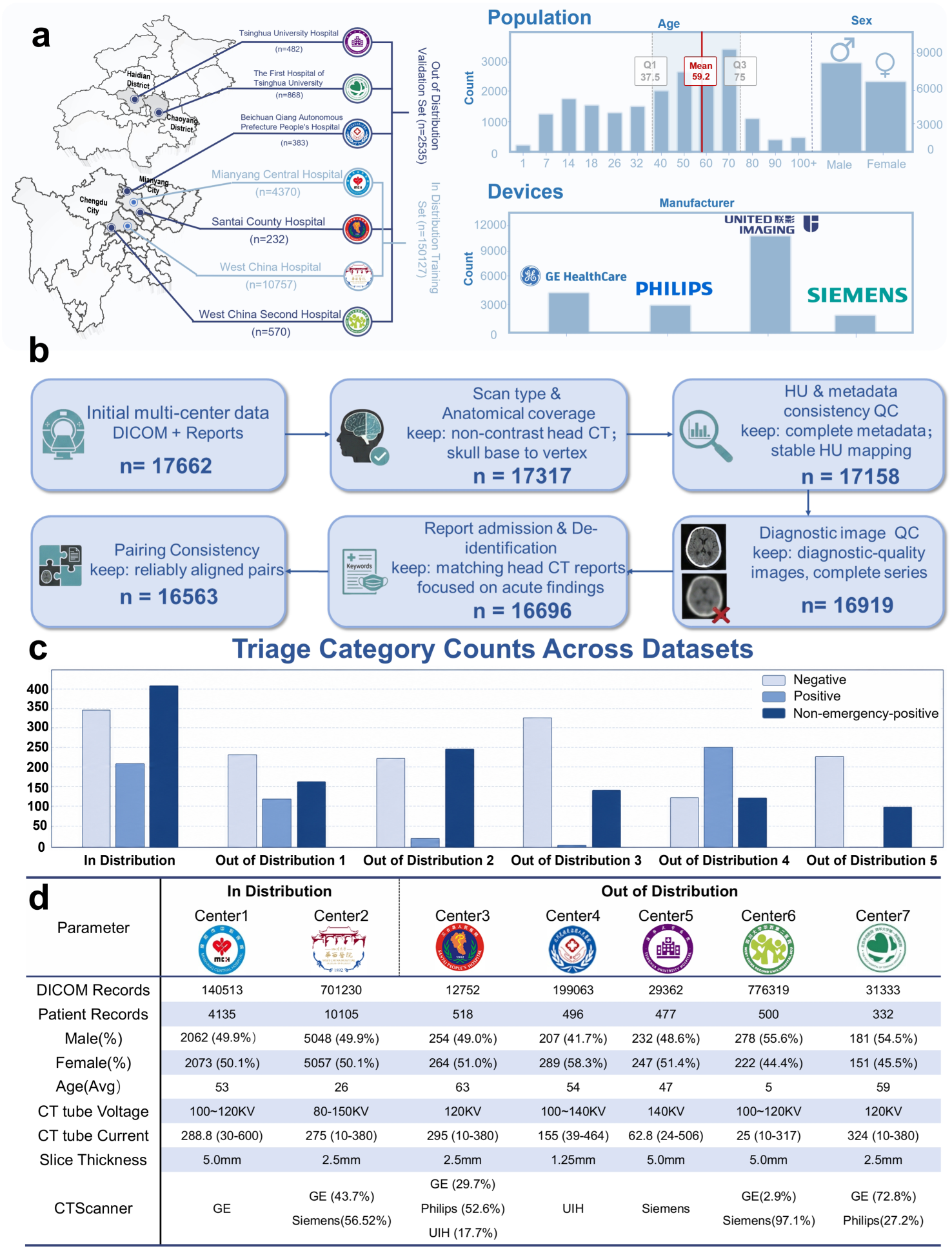
Construction and characterization of the multicenter head CT cohort. a, Geographic distribution of participating hospitals and cohort allocation into development, in-distribution evaluation and out-of-distribution evaluation sets. b, Cohort assembly and quality-control workflow from the initially collected DICOM examinations and reports to the final paired examination-level cohort. c, Distribution of emergency triage categories across the internal cohort and five external cohorts. d, Center-level demographic and acquisition characteristics, including DICOM record counts, patient counts, sex distribution, age, tube voltage, tube current, slice thickness and scanner manufacturer. Together, these panels illustrate the scale, quality-control process and clinically interpretable sources of distribution shift in the multicenter evaluation.

We calculated all performance metrics at the CT-volume level. We compared CHIEF with all publicly available methods: (1) medical vision–language foundation models (CT-CLIP^35^, Med-BLIP^36^, Merlin^37^, and PubMedCLIP^38^); and (2) commercial multimodal large language models (MLLMs; GPT-5, Gemini 3 Pro, Claude 4.5 Sonnet, and Qwen 3 Instruct). Because rapid separation of negative, non-emergency-positive, and positive examinations is the most immediate need in emergency head CT workflows, we treated emergency triage classification as the central clinical task of CHIEF. We evaluated report generation, image-to-text retrieval, and abnormality detection as complementary interfaces that translate the same pretrained representation into report-level communication, reference-case support, and fine-grained recognition. We provide details of the CHIEF architecture, pretraining procedure, and cohort curation in the Methods.

### Emergency triage classification as the central clinical endpoint

We next evaluated whether the unified pretrained representation could be transferred to examination-level emergency triage. In routine emergency head CT workflows, triage enables the rapid prioritization of urgent cases by classifying each examination as negative, non-emergency-positive, or positive. To this end, we trained a lightweight classification head on top of the unified pretrained representation to assign each examination to one of the three triage categories. Examination-level reference labels were assigned by radiologist annotators from the final radiology reports using predefined urgency-based criteria, with these reports used solely as sources of reference labels rather than as model inputs. Because this task requires both abnormality detection and discrimination among urgency categories under substantial class imbalance, we accordingly evaluated performance with an emphasis on sensitivity for positive examinations while maintaining specificity across the remaining categories.

On the ID test dataset, CHIEF achieved the best performance across all metrics (Fig. 2; Supplementary Table 1). Specifically, CHIEF achieved an Accuracy of 0.8688, a balanced accuracy (BACC) of 0.8673, an F1 score of 0.8673, and an AUROC of 0.9646. Compared with CT-CLIP (accuracy = 0.4225), CHIEF more than doubled the Accuracy. Under class-imbalanced conditions, CHIEF maintained a BACC of 0.8673 and a G-mean of 0.8983, demonstrating balanced recognition across categories rather than reliance on the majority class. These performance gains were statistically significant (Supplementary Table 1), supporting the effectiveness of CHIEF for emergency head CT triage..

**Fig. 2.**
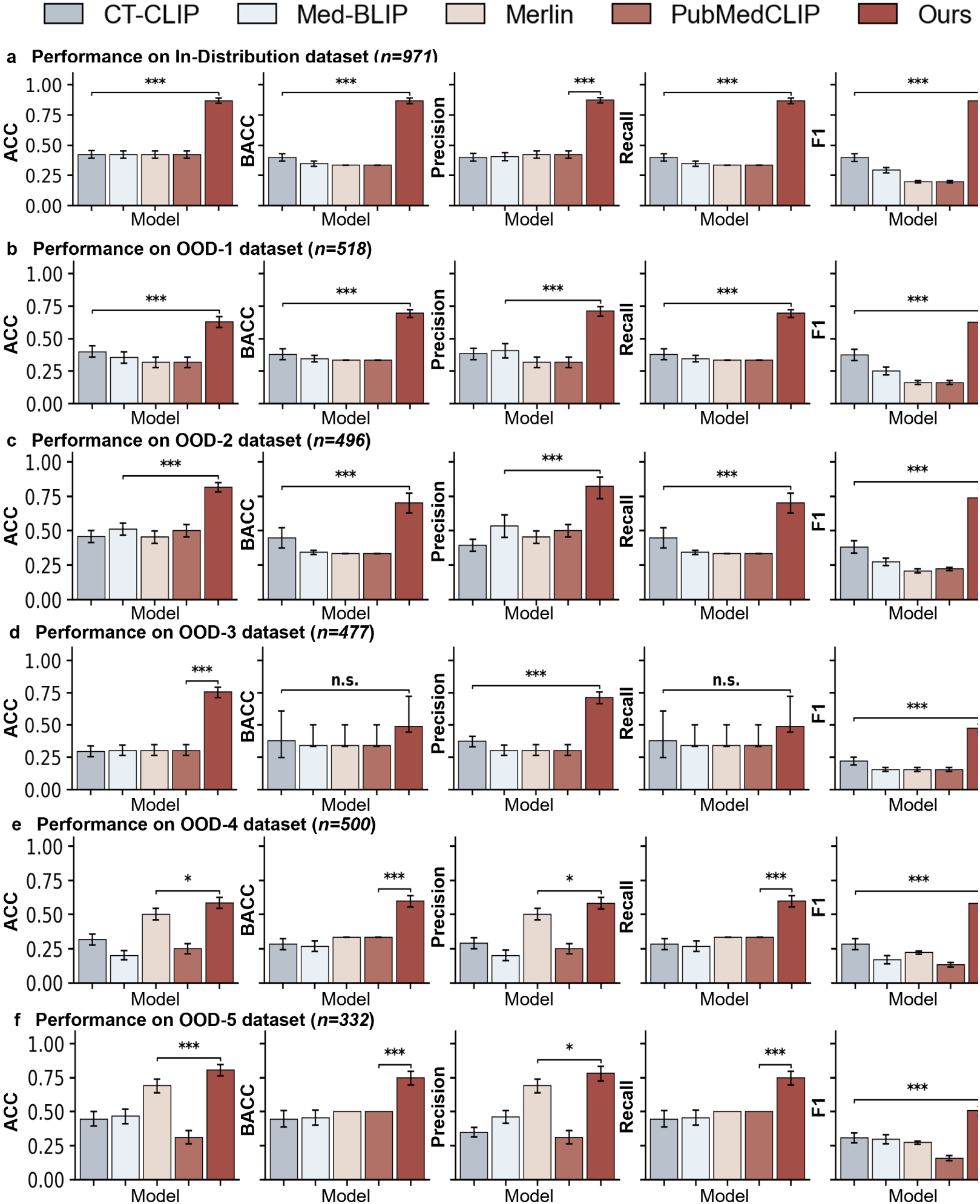
Performance on the in-distribution dataset and five out-of-distribution datasets. a, Performance comparison on the in-distribution dataset. Model performance is shown for ACC, BACC, precision, recall and F1, with comparison to CT-CLIP, Med-BLIP, Merlin and PubMedCLIP. b-f, Performance comparison on five out-of-distribution datasets. Model performance is shown for ACC, BACC, precision, recall and F1 on OOD-1, OOD-2, OOD-3, OOD-4 and OOD-5, respectively, with comparison to CT-CLIP, Med-BLIP, Merlin and PubMedCLIP. Error bars indicate 95% confidence intervals estimated by sample-level bootstrap resampling.

Across the five OOD datasets, CHIEF outperformed the comparator models on most metrics. Specifically, on the OOD-1 dataset, CHIEF achieved an Accuracy of 0.6278 and an AUROC of 0.8517. On the OOD-2 dataset, CHIEF achieved an Accuracy of 0.8167 and an AUROC of 0.9173. On the OOD-3 dataset, CHIEF achieved an AUROC of 0.8319. On the OOD-4 dataset, which featured a markedly different patient population, CHIEF achieved an accuracy of 0.5838 and an AUROC of 0.7753, representing the largest reduction in performance relative to the ID dataset. On the OOD-5 dataset, CHIEF achieved an Accuracy of 0.8065 and a Specificity of 0.8314. Despite variations in imaging equipment, scanning protocols, and patient composition across sites, CHIEF consistently outperformed the comparator models on all external datasets.

### Triage-oriented report generation compared with medical vision-language models

We next evaluated report generation as a complementary interface for communicating imaging findings and risk-relevant information. Compared with general radiology report generation, emergency head CT report generation places greater emphasis on comprehensive reporting of critical findings, including their spatial distribution and clinical significance, to support rapid triage and clinical decision-making. We therefore used 3D head CT volumes, rather than only the 2D slices used by conventional methods, to generate corresponding radiology reports and systematically evaluated report quality using BLEU^39^, ROUGE-L^40^, METEOR^41^, CIDEr^42^, and BERTScore^43^.

On the ID test dataset, CHIEF achieved the best performance across all language generation metrics, with a BLEU of 0.2305, a ROUGE-L of 0.2912, a METEOR of 0.3813, a CIDEr of 0.5032, and a BERTScore of 0.7559 (Supplementary Table 2). Compared with CT-CLIP, the strongest existing medical foundation model, CHIEF improved BLEU by 57.9%, ROUGE-L by 22.1%, METEOR by 15.5%, and CIDEr by 66.4%, and these differences were statistically significant (*p* < 0.05). Notably, the consistent gains on METEOR and CIDEr, which place greater emphasis on semantic consistency, suggest that CHIEF better captures emergency-critical findings and risk-related semantics in the generated reports.

On the OOD datasets, CHIEF remained superior on most metrics under the same report preprocessing, Chinese tokenization and scoring implementation used for all models and cohorts. Its performance, however, decreased relative to the ID dataset and varied across centers (Supplementary Table 2). For example, on the OOD-1 dataset, CHIEF achieved a CIDEr of 0.8649, substantially higher than that of CT-CLIP (0.0363). On the OOD-2 dataset, CHIEF achieved a METEOR of 0.3047, representing an improvement of more than 50% over Merlin. On the OOD-3 dataset, CHIEF achieved BLEU and ROUGE-L scores of 0.2061 and 0.2834, respectively, both of which were the highest among the evaluated methods. On the OOD-5 dataset, CHIEF achieved a BLEU of 0.1116 and a BERTScore of 0.6626.

CHIEF produced more clinically complete and spatially specific reports than existing medical vision-language models, suggested by case-level sentence-by-sentence comparisons (Fig. 3). The comparison models often omitted information or lacked sufficient specificity in describing key abnormalities. For example, in cases of intracranial hemorrhage, these methods often produced vague statements such as “abnormal density shadow” or “no obvious abnormality,” without clearly specifying the location, extent, or associated risk signs of the hemorrhage. By contrast, CHIEF accurately localized abnormal hyperdense foci and described their spatial distribution and morphological characteristics, yielding a more complete diagnostic interpretation. In addition, when potentially dangerous signs were present, CHIEF explicitly reported key findings such as “midline shift,” whereas other methods tended either to omit these signs or to describe them imprecisely. Overall, CHIEF was more consistent in localizing abnormalities, structuring descriptions and reporting high-risk signs, and its reports more closely resembled emergency radiology reports in readability and clinical actionability.

**Fig. 3.**
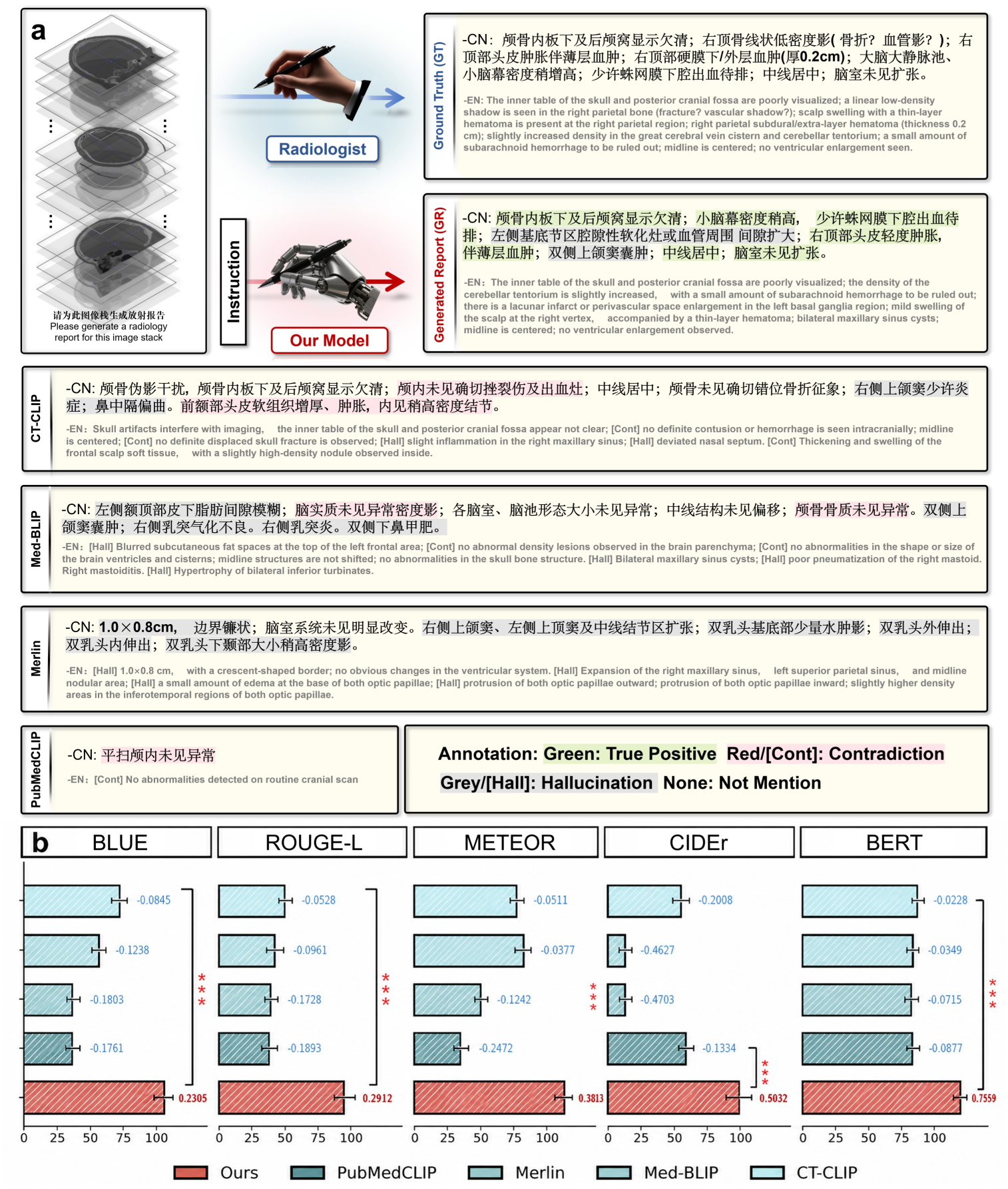
Report generation compared with existing medical vision-language models. a, Representative example of report generation for a head CT examination. The radiologist-written reference report is shown together with the report generated by CHIEF and reports generated by existing medical vision-language models, including CT-CLIP, Med-BLIP, Merlin and PubMedCLIP. Expert reviewers annotated report content to indicate correctly described findings, contradictions to the imaging findings, hallucinated content and abnormalities that were not mentioned. These examples illustrate differences in factual accuracy, hallucination and omission across models. b, Comparison of CHIEF and existing medical vision-language models using conventional automatic report generation metrics, including BLEU, ROUGE-L, METEOR, CIDEr and BERTScore. Although these metrics quantify textual overlap or semantic similarity with the reference report, they do not directly assess clinical correctness. Together, the qualitative expert evaluation in a and the quantitative metric comparison in b highlight the importance of expert review for assessing the clinical reliability of generated radiology reports. For visualization, all automatic metric values in b are multiplied by 100; values in the text and supplementary tables are reported on the original 0-1 scale.

### Report generation compared with commercial multimodal large language models

On the ID test dataset, CHIEF generated substantially higher-quality reports than every commercial MLLM under the same protocol (Supplementary Table 3). Its BLEU score of 0.2305 exceeded the best commercial model by more than ten-fold (GPT-5, 0.0181; Gemini 3 Pro, 0.0172; Claude 4.5 Sonnet, 0.0130; Qwen 3 Instruct, 0.0143). CHIEF likewise led on semantically oriented metrics, reaching 0.3813 METEOR and 0.5032 CIDEr, against a best commercial METEOR of 0.1461 (GPT-5) and near-zero CIDEr for the commercial models.

These results indicate that, although commercial MLLMs produce grammatically coherent text, they struggle to accurately express key abnormal findings, their spatial distribution, and risk-related information in emergency head CT.

CHIEF maintained the same advantage across all five OOD datasets (Supplementary Table 3). Its BLEU scores (0.1803, 0.1419, 0.2061, 0.1249, and 0.1116 for OOD-1 to OOD-5) remained markedly higher than those of the commercial MLLMs, which ranged from 0.004 to 0.065. CHIEF also led on ROUGE-L and METEOR (0.1416 to 0.3052 and 0.2097 to 0.3387 across centers), exceeding the best commercial values on the corresponding datasets (ROUGE-L 0.1803, METEOR 0.2911, both GPT-5), though its performance remained center-dependent. The gap was widest for CIDEr, which rewards semantic consistency: on OOD-1, CHIEF reached 0.8649, whereas GPT-5, Gemini 3 Pro, and Claude 4.5 Sonnet were all near zero.

In case-level analysis, CHIEF described critical findings more completely and specifically than the commercial MLLMs. These models tended to produce generalized or template-like descriptions, omit key abnormal findings, or fail to clearly specify their spatial locations and potential risks. For example, in cases of intracranial hemorrhage, some models produced only statements such as “no obvious abnormality” or similarly vague descriptions, failing to identify hyperdense hemorrhagic foci or associated dangerous signs. Missing or incomplete reporting was also common for critical emergency findings (e.g., midline shift). By contrast, CHIEF more consistently identified key abnormalities (e.g., intracranial hemorrhage and fracture) and described their spatial distribution and associated risk signs, thereby producing reports more consistent with emergency radiology reporting conventions.

### Blinded *Turing* test of CHIEF-generated radiology reports

We designed a blinded *Turing* test to assess how distinguishable CHIEF’s generated reports were from radiologist-written reports under clinical interpretation (Fig. 4a). This was necessary because automatic metrics (e.g., BLEU and ROUGE-L), on which CHIEF already scored highest, capture mainly lexical and syntactic overlap and cannot fully reflect semantic accuracy, diagnostic plausibility, or interpretive value. Specifically, for each case, a paired sample consisting of a CHIEF-generated report and the corresponding ground-truth report was constructed and presented to radiology experts in random order. Under blind conditions, the experts were asked to determine which report had been generated by AI (Q1), provide a confidence score for that judgment (Q2), and record the basis for their decision (Q3). This setting was intended to evaluate, from the perspective of clinical interpretation, how closely the generated reports approximated radiologist-written reports in terms of structural organization, semantic expression, and diagnostic consistency.

**Fig. 4.**
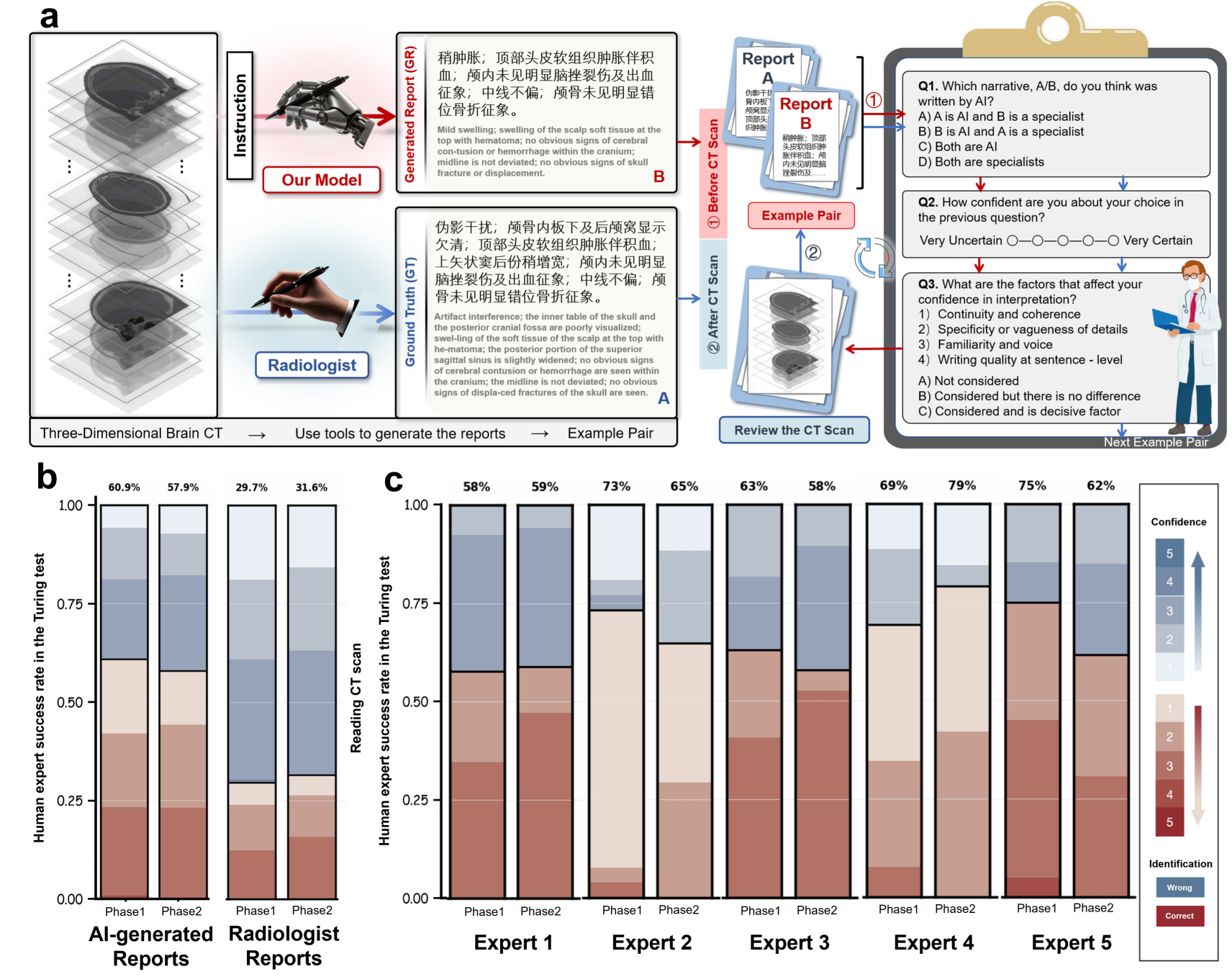
Blinded *Turing* study of CHIEF-generated and radiologist-written head CT reports. a, Study design. Each 3D head CT examination was paired with two anonymized reports, one written by a board-certified radiologist and one generated by CHIEF, and presented in random order. Expert readers reviewed the CT images, identified which report they believed had been written by a radiologist, recorded their confidence and indicated the factors influencing their judgment, including report coherence, finding specificity, writing style and linguistic quality. b, Overall results. The stacked bars show how often CHIEF-generated and radiologist-written reports were judged as human-written or AI-generated across study phases. A high proportion of CHIEF-generated reports judged as human-written indicates limited distinguishability under blinded review. c, Reader-level results. The distribution of judgments is shown for each expert reader and study phase, with values above the bars indicating overall identification accuracy. Together, these results suggest that expert readers had limited ability to distinguish CHIEF-generated reports from radiologist-written reports.

In the blinded *Turing* test, CHIEF-generated reports were largely indistinguishable from radiologist-written reports, with experts’ identification accuracy near chance level. Specifically, CHIEF-generated reports were judged as human-written in approximately 60% of cases (Fig. 4b). In addition, the evaluation results showed a consistent trend across experts (Fig. 4c), suggesting that this phenomenon was relatively stable.

### Cross-modal retrieval between CT volumes and radiology reports

We next examined whether the shared semantic space learned by CHIEF supports clinically meaningful correspondence between CT volumes and radiology reports, using cross-modal retrieval as the probe. We used each 3D head CT volume as an image query to retrieve the paired radiology report from a candidate report set. We adopted this image-to-text setting to simulate the clinically relevant scenario in which a newly acquired emergency head CT examination is matched to prior radiology reports with similar imaging semantics. Within this setting, CHIEF projects the query CT volume and candidate reports into a unified feature space and performs similarity-based matching, retrieving reports by image–text semantic similarity (Fig. 5a). We evaluated ranking quality and semantic matching using Recall@K, MRR, and NDCG (Methods).

**Fig. 5.**
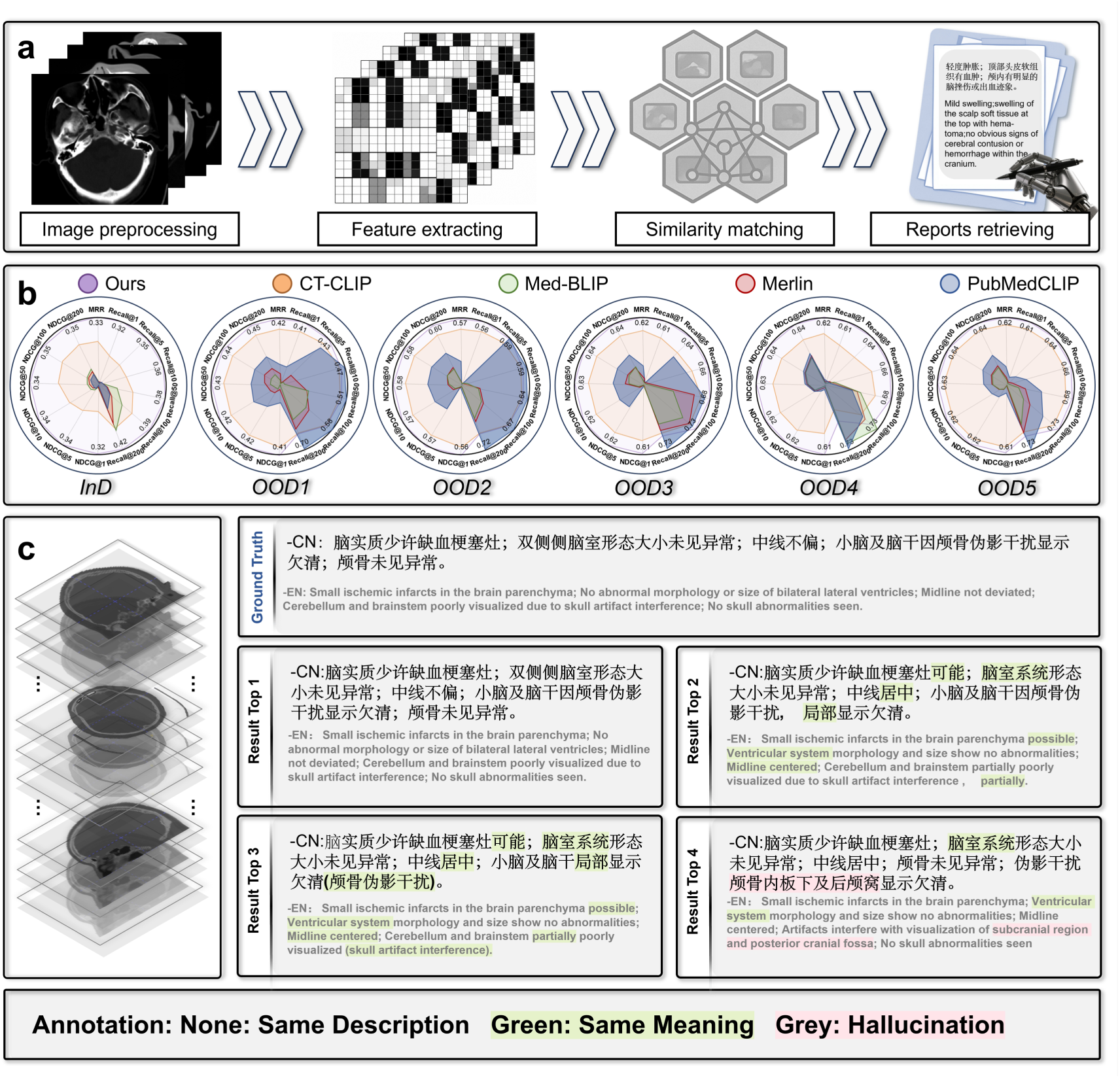
Image-to-text retrieval of clinically similar reports from head CT examinations across centers. a, Schematic overview of the image-to-text retrieval workflow. A query head CT examination is preprocessed and encoded into the shared representation space, where similarity matching is performed against archived report representations to retrieve clinically similar radiology reports. b, Quantitative evaluation of retrieval performance in the in-distribution cohort and five out-of-distribution cohorts. Radar plots summarize Recall@K, mean reciprocal rank and normalized discounted cumulative gain at multiple cutoffs for CHIEF and comparator methods across cohorts. c, Representative retrieval example. A head CT examination was used as the query, and the corresponding radiologist-written report is shown together with the top-ranked retrieved reports. Retrieved reports largely retained the core clinical meaning of the reference report, including small ischemic findings, preserved ventricular morphology, absence of midline shift and limited assessment of the posterior fossa because of skull-related artifact. Text annotations indicate exact or near-exact descriptions, semantically concordant content and unsupported or hallucinated content in retrieved reports.

On the ID test dataset, CHIEF achieved the best performance across all retrieval metrics (Fig. 5b; Supplementary Tables 4 and 5). Specifically, CHIEF attained an MRR of 0.3383, substantially surpassing that of CT-CLIP (0.1421). For the Recall@K metrics, Recall@1, Recall@5, and Recall@10 reached 0.3223, 0.3512, and 0.3553, respectively, all markedly higher than those of CT-CLIP (0.1267, 0.1493, and 0.1627, respectively). These results indicate that CHIEF ranks semantically matched image–text pairs more accurately near the top of the candidate list.

On the OOD datasets, CHIEF maintained strong retrieval performance (Fig. 5b; Supplementary Tables 4 and 5). For example, on the OOD-2 dataset, CHIEF achieved an MRR of 0.5727 and a Recall@1 of 0.5565. On the OOD-3 dataset, CHIEF achieved an MRR of 0.6244 and a Recall@1 of 0.6080. On the OOD-4 dataset, which featured a markedly different patient population, CHIEF still maintained an MRR of 0.2313 and a Recall@1 of 0.2220. CHIEF achieved consistent NDCG performance, remaining superior across multiple OOD datasets. Qualitative examples further illustrate this semantic matching (Fig. 5c). For a given CT query, CHIEF retrieved top-ranked reports that were highly consistent with the reference report, accurately describing key imaging findings with only minor lexical differences and rarely producing overt semantic deviations or hallucinated content. These findings indicate robust image–text alignment across the evaluated cohorts.

### Zero-shot abnormality detection via language-guided reasoning

Beyond the supervised triage, report-generation and retrieval tasks, we evaluated whether CHIEF could identify abnormal findings without task-specific fine-tuning. This analysis was performed on the internal matched cohort using report-derived abnormality labels. Specifically, for each predefined abnormality type, a question template was constructed, and abnormality probabilities were estimated by leveraging the generative preference of the language model for “yes” or “no” responses.

Across 45 report-derived abnormality labels on the internal matched cohort (971 examinations), CHIEF showed strong fine-grained discrimination but limited overall binary separation (Fig. 6a–c; Supplementary Table 6). For the overall binary abnormality-detection task, defined as normal versus any abnormality, CHIEF achieved an AUROC of 0.6255 (95% CI, 0.5880–0.6625) and an AP of 0.7465 (95% CI, 0.7092–0.7860), indicating limited discrimination between normal examinations and those containing any report-derived abnormality. At the fine-grained label level, CHIEF reached a macro-AUROC of 0.9472 and a micro-AUROC of 0.9698, with a macro-AP of 0.3973 and a micro-AP of 0.1600. CHIEF also attained a micro-specificity of 0.9920. Because positive instances were sparse across many labels, these lower AP values reflect class imbalance rather than uniformly weak positive-label detection. CHIEF assigned high predicted probabilities to clinically relevant findings such as subdural hemorrhage, cerebral atrophy, middle ear mastoiditis and hydrocephalus. The accompanying label-distribution panel reports reference positive counts separately from CHIEF-derived prediction scores to avoid conflating disease prevalence with model confidence.

**Fig. 6.**
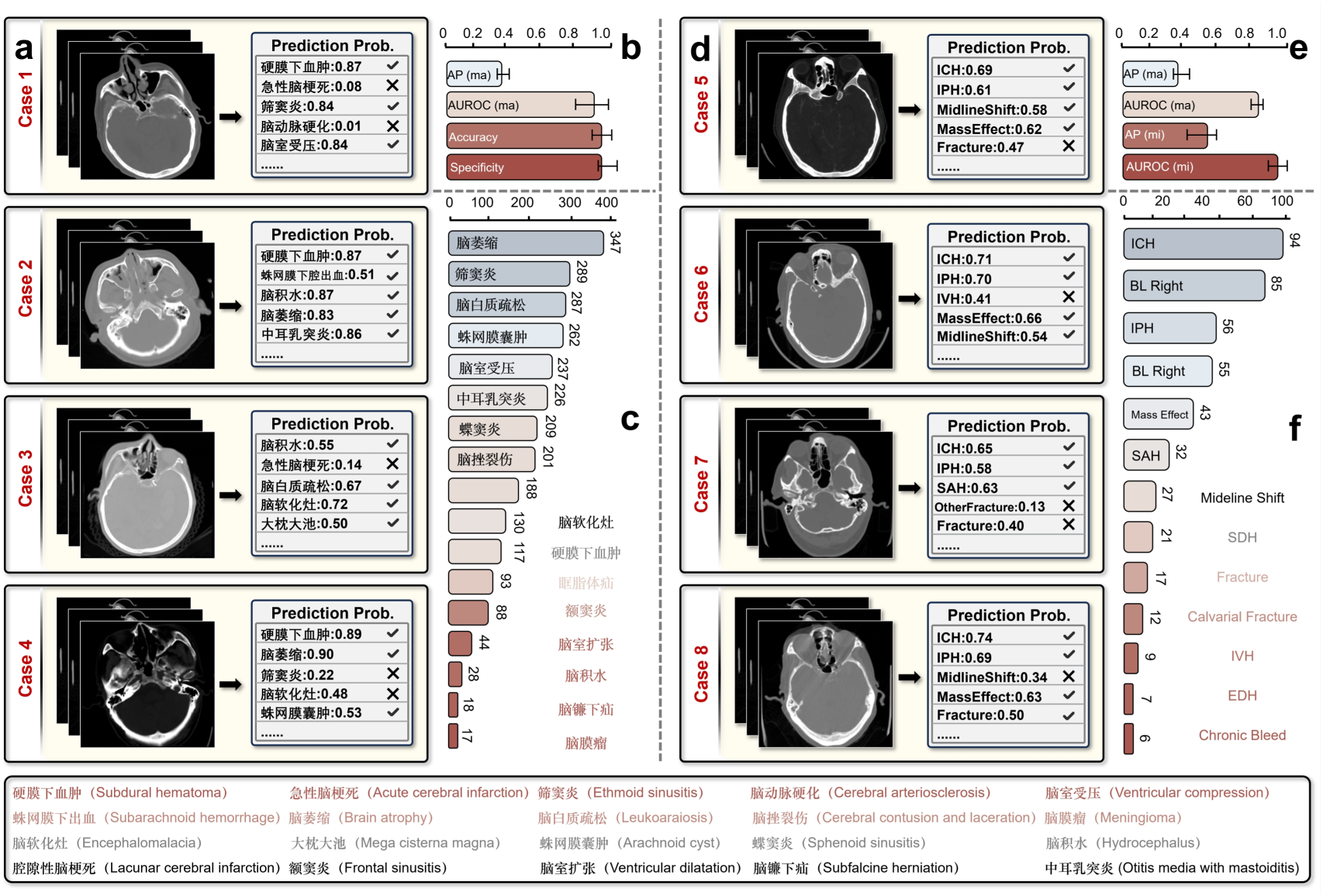
Abnormality detection in the internal matched cohort and CQ500 benchmark. a–c, Zero-shot abnormality detection in the internal matched cohort using report-derived abnormality labels. Representative cases are shown with selected CT slices, predicted abnormality probabilities and agreement markers relative to reference labels. The summary panels report overall abnormality detection and fine-grained multi-label detection performance, with reference positive counts separated from model-derived prediction scores. d–f, Fine-tuned multi-label classification on the CQ500 validation set for hemorrhage and fracture-related abnormalities. Representative cases, macro- and micro-averaged performance, and label-level positive counts are shown. Together, these results indicate that CHIEF supports both language-guided zero-shot abnormality detection on internal report-derived labels and fine-tuned multi-label abnormality classification on an external public benchmark.

### Fine-tuned multi-label abnormality classification on CQ500

To further assess whether the pretrained representations could transfer to an external public benchmark with a structured label system, we evaluated CHIEF on CQ500 using a fine-tuned multi-label classification probe. This task was distinct from the internal zero-shot analysis: the CQ500 probe used supervised fine-tuning and evaluated 14 hemorrhage and fracture-related labels on the validation set. The 491 patients were partitioned into training and validation subsets at the patient level, with no patient contributing CT volumes to both subsets. This yielded 1,019 training CT volumes and 245 validation CT volumes, and all performance metrics were calculated at the CT-volume level.

On the CQ500 validation set, CHIEF achieved a macro-AUROC of 0.8033 (95% CI, 0.7689–0.8425) and a micro-AUROC of 0.8466 (95% CI, 0.8194–0.8750) (Fig. 6d–f; Supplementary Table 6). The corresponding macro-AP and micro-AP were 0.3924 (95% CI, 0.3405–0.4789) and 0.4743 (95% CI, 0.3991–0.5536), respectively, indicating stable transfer to this external benchmark. In label-wise analysis, CHIEF showed stronger discrimination for several clinically important findings, including intraventricular hemorrhage, fracture, calvarial fracture and midline shift, whereas AP was lower for labels with few positive validation cases, such as subdural hemorrhage and chronic bleed (Supplementary Table 7). For representative CQ500 cases, CHIEF assigned high probabilities to intracranial hemorrhage, intraparenchymal hemorrhage, subarachnoid hemorrhage, midline shift, mass effect and fracture when these findings were present, while assigning lower probabilities to absent findings.

## Discussion

In this study, we show that unified 3D vision–language pretraining can turn routine emergency head CT and paired reports into a single, annotation-free model that both triages studies and drafts radiology reports. We developed and evaluated this model, CHIEF, on 16,563 examination-level CT–report pairs from seven institutions. We evaluated it across five emergency tasks, centered on triage classification and complemented by report generation, cross-modal retrieval, zero-shot abnormality detection, and fine-tuned abnormality classification. In a blinded *Turing* test, radiologists mistook its generated reports for human-written ones in approximately 60% of cases. Together, these results show that unified, annotation-free representation learning can provide a scalable, clinically grounded foundation for triage-oriented head CT interpretation in real-world emergency settings.

For emergency triage, the endpoint of most direct clinical consequence, CHIEF preserved balanced discrimination across urgency categories despite marked class imbalance. Rather than merely flagging the presence of an abnormality, CHIEF sorts each examination into three urgency-aligned categories (negative, non-emergency-positive, or positive) that mirror the disposition decision governing how quickly a patient is seen (Fig. 2). The clinical challenge here is class imbalance: the positive examinations that most demand rapid escalation are also the rarest. A model that defaults to the majority class would appear accurate yet fail precisely when it matters most. That CHIEF avoided this failure indicates its prioritization signal reflects genuine pathological urgency rather than the underlying prevalence. Such behavior is a prerequisite for a triage aid that clinicians can act on with confidence.

CHIEF consistently and markedly outperformed all commercial MLLMs across every evaluation cohort. Its BLEU scores were more than ten-fold higher, and its CIDEr far exceeded the near-zero values of even the strongest commercial models (GPT-5, Gemini 3 Pro, Claude 4.5 Sonnet, and Qwen 3 Instruct). This gap challenges the assumption that scale and generality alone can offset the absence of domain-specific visual grounding in clinical imaging^44,45^. Case-level analysis (Supplementary Fig. 7) exposed two complementary failure modes. First, these models hallucinated, generating linguistically fluent but factually unsupported findings^46^. Second, they lost critical information: high-risk features such as midline shift, mass effect and hemorrhage extent were omitted or described with insufficient spatial specificity^32,47^. These failures are not incidental; they reflect a fundamental mismatch between general-purpose model architecture and the problem structure of emergency head CT. Here, key findings depend on continuous spatial relationships across slices rather than local textures within a single image^24,48^. Lacking native 3D volumetric encoding, commercial MLLMs reduce these spatially continuous pathological patterns to fragmented 2D representations, irrecoverably forfeiting the cross-slice context on which accurate emergency diagnosis depends^24,44^.

CHIEF-generated reports proved clinically trustworthy, largely indistinguishable from those written by attending radiologists. In a blinded *Turing* test, attending radiologists misidentified them as human-written in approximately 60% of cases (Fig. 4b). Consistent misidentification across all expert readers indicates that the reports followed established radiology reporting conventions. Conventional natural language generation (NLG) metrics fail to capture this fidelity: BLEU, ROUGE and CIDEr reward surface-level lexical overlap rather than the diagnostic accuracy that clinical validity demands^49–52^. The danger is concrete: a single-word laterality error, substituting right for left temporal hemorrhage, incurs negligible penalty yet reverses the surgical target. These metrics also over-penalize harmless stylistic variation and overlook the hierarchical logic that makes a report actionable. By faithfully conveying the spatial precision and risk stratification that time-sensitive neurosurgical planning depends on, CHIEF-generated reports may support more structured communication in emergency workflows. Realizing this potential will require prospective evaluation.

Beyond triage and report generation, CHIEF functions as a versatile clinical engine rather than a single-task solution. Without any architectural change or retraining of its shared encoder, CHIEF supports cross-modal retrieval, zero-shot abnormality detection across 45 report-derived labels, and fine-tuned multi-label classification on CQ500, implementing each capability as a lightweight downstream probe rather than an independently optimized system (Supplementary Table 8). Unlike the prevailing paradigm of task-specific models, which demand independent architectural design, dense annotation and end-to-end optimization for every clinical endpoint and often converge on dataset-specific shortcuts rather than transferable clinical semantics^44,53^, CHIEF achieves this breadth through three complementary pretraining objectives: contrastive image-text alignment shapes cross-modal discriminative geometry; generative semantic anchoring prevents the collapse of fine-grained semantic distinctions that contrastive learning alone may discard; and decorrelation regularization suppresses protocol-driven dimensional redundancy. CHIEF is especially advantageous for long-tail abnormalities, precisely the categories where specialist models are most vulnerable to data-scarcity-driven underfitting, retaining robust discrimination even for findings present in fewer than 5% of cases (Supplementary Table 7).

CHIEF remained robust across five clinically diverse external hospitals, confirming that heterogeneous multi-institutional pretraining is decisive for overcoming data drift, the single most persistent barrier to clinical AI deployment^54,55^. This barrier is especially acute in emergency head CT^17^, where even moderate acquisition shifts can disrupt the volumetric context that accurate diagnosis relies on^19^. Yet existing models typically validate on only one or two external sites within the same health system, leaving their robustness to genuine acquisition and population heterogeneity untested^25,26^. CHIEF was instead evaluated within an OOD framework that stratifies external sites by clinically interpretable sources of distribution shift. County-level primary hospitals (OOD-1 and OOD-2) introduce acquisition shifts from lower-end scanner hardware and non-standardized protocols. A pediatric specialty hospital (OOD-4) imposes a markedly different age structure and disease spectrum. Independent general hospitals in a geographically distinct region (OOD-3 and OOD-5) contribute further variation in class proportions and reporting conventions. Under these compounding shifts, CHIEF exhibited the smallest performance degradation of all evaluated models across report generation, triage classification and retrieval (Extended Data Fig. 8), whereas comparator medical vision–language models showed substantially larger and less predictable drops.

CHIEF is positioned as a clinical decision-support framework for emergency workflows rather than an autonomous tool intended to replace radiologists^56^. By learning from paired CT-report data without task-specific manual labels, CHIEF could support automated draft generation, triage classification, urgent-finding alerts and abnormality detection across heterogeneous clinical environments, which are especially valuable for resource-limited institutions where subspecialty expertise is scarce^45^. Its integrated cross-modal retrieval further supports reference-based decision-making by surfacing semantically matched prior reports, reducing the cognitive burden of high-volume emergency reading. In scenarios such as night shifts, peak surges, primary hospitals or pre-transfer assessment, CHIEF can translate high-risk three-dimensional findings into structured, management-oriented textual outputs, shortening the pathway from image acquisition to clinical action. CHIEF currently demonstrates foundational capacity for workflow assistance under multitask and multicenter conditions but does not yet establish independent clinical readiness, which will require prospective deployment, physician-interaction studies, and assessment of patient outcomes^57^.

Several limitations warrant consideration. First, CHIEF is pretrained predominantly on non-contrast head CT for acute neurological conditions. Although the underlying cohort is large and multicenter, its transferability to contrast-enhanced scans, broader neuroimaging tasks or other organ systems remains unverified. Future work should therefore expand the pretraining distribution accordingly. Second, both the pretraining supervision and the emergency triage reference labels were derived from real-world radiology reports rather than expert-adjudicated image-based ground truth. As a result, CHIEF may absorb the stylistic omissions, templated phrasing and reporting biases present in the original texts^29^. We consider this a reasonable trade-off for the annotation-free scalability that makes multicenter pretraining feasible at this scale. Future iterations could apply automated text refinement to improve training-signal fidelity. Third, the current framework lacks fine-grained lesion segmentation and temporal disease modeling. Finally, all evaluations were conducted retrospectively, and prospective clinical validation is therefore needed to establish the translational utility of CHIEF for triage efficiency and patient outcomes^30^.

## Online Methods

### Multicenter cohort construction and distribution-shift evaluation design

We built an examination-level cohort of paired non-contrast 3D head CT scans and Chinese radiology reports to train and evaluate CHIEF for emergency head CT interpretation. We treated each emergency examination as the unit of analysis and paired every CT volume one-to-one with the report from the same clinical encounter. We drew cases from tertiary general hospitals, county-level hospitals and a pediatric specialty hospital, so the cohort spans diverse scanner hardware, acquisition protocols, reconstruction parameters, patient populations and disease spectra. To improve pairing reliability and reduce semantic noise, we applied hierarchical quality-control criteria at data entry, requiring physical consistency, diagnostic image quality, report relevance and correct examination-level pairing as joint inclusion constraints (Fig. 1a,b).

For imaging, we kept only complete non-contrast head CT volumes covering the whole head, from the skull base to the vertex. We required complete DICOM metadata and a stable mapping from pixel values to the Hounsfield unit (HU) scale, so intensity calibration stayed comparable across scanners. We then removed series with missing calibration information, abnormal HU mapping, or intensity distortion from truncation or saturation. We further excluded examinations with severe motion or metal artifact, high noise, missing slices, incomplete series or grossly abnormal geometry, so that degraded scans would not inject systematic bias. For text, we kept only reports describing a non-contrast head CT. We required each report to focus on intracranial structures and key emergency findings, and we discarded reports that mixed other body parts, gave uninformative templated statements, or were contaminated by copy-and-paste. At the pairing stage, we kept only report versions that we could reliably align to the same-visit examination on the basis of examination identifiers and timestamps. We excluded any record with an unclear rescan relationship, a combined examination across body parts, or a clear mismatch between image and text. Through this standardized inclusion pipeline, we raised the signal-to-noise ratio and cross-center consistency of the paired data without adding any manual annotation. This gave representation learning more stable supervision while keeping the real distributional spread of multicenter data, so that we improved learning stability and generalization together.

To systematically evaluate cross-institution generalization under real clinical deployment conditions, we established an ID and OOD evaluation framework using medical institutions as the stratification unit, with cross-center transfer as the central methodological axis. The final self-constructed multicenter cohort comprised 16,563 examinations. Cases from West China Hospital (*n* = 10,105) and Mianyang Central Hospital (*n* = 4,135) constituted 14,240 ID examinations, including 13,269 CT–report pairs used for self-supervised pretraining and a held-out ID test set of 971 examinations. Cases from Santai County People’s Hospital (*n* = 518), Beichuan Qiang Autonomous County People’s Hospital (*n* = 496), Tsinghua University Hospital (*n* = 477), Beijing Huaxin Hospital (The First Hospital of Tsinghua University; *n* = 332) and West China Second University Hospital, Sichuan University (West China Women’s and Children’s Hospital; *n* = 500) constituted the OOD data and were used to simulate the cross-center distribution shifts that are unavoidable in real-world deployment (Fig. 1c). Importantly, OOD in this study was not defined simply as data from other hospitals; rather, it was designed to capture distribution shifts with clear clinical interpretability. County-level primary hospitals mainly represented acquisition shifts arising from differences in equipment conditions and scanning protocols. The pediatric specialty hospital introduced a markedly different age structure and disease spectrum. The remaining independent institutions further contributed variation in class proportions and clinical composition, thereby enabling evaluation of transfer stability under both population distribution shift and label distribution shift. This institution-stratified design, with interpretable sources of shift, moved model evaluation beyond comparisons based solely on aggregate metrics and enabled repeatable, attributable quantitative assessment of the transferability and robustness of the pretrained representations under conditions more closely resembling real-world deployment.

In addition to the self-constructed multicenter 3D CT-text cohort, we introduced CQ500 as an external benchmark for downstream multi-label abnormality classification. CQ500 provides radiologist-annotated labels for intracranial hemorrhage, hemorrhage subtypes, skull fracture, mass effect and midline shift, and has been widely used for evaluating emergency head CT abnormality detection. In this study, CQ500 was used in a fine-tuned downstream setting rather than as a zero-shot benchmark. The 491 patients were partitioned into training and validation subsets at the patient level, with no patient contributing CT volumes to both subsets. This partition yielded 1,019 training CT volumes and 245 validation CT volumes, and all reported CQ500 performance metrics were computed at the CT-volume level on the validation subset. Fourteen hemorrhage and fracture-related labels were treated as independent binary classification endpoints. By combining multicenter real-world evaluation with public-benchmark validation, this study further strengthened the reproducibility and horizontal comparability of the conclusions while preserving clinical representativeness.

### Emergency triage label definition and annotation

We annotated examination-level emergency triage labels by asking radiologists to read the final non-contrast head CT reports for each examination. We used these reports only to define reference labels, not as model inputs for triage classification. We asked annotators to review both the Findings and Impression sections of each report and to classify each examination as Positive, non-emergency-positive or negative. The labels reflected the urgency of the abnormalities documented in the report rather than the overall severity of the patient’s clinical condition, final clinical diagnosis, hospital admission or need for surgery. Positive examinations contained acute, recent, progressive or potentially rapidly deteriorating abnormalities that generally required immediate clinical evaluation or management. Non-emergency-positive examinations contained chronic, stable, mild or incidental abnormalities without acute high-risk features. Negative examinations contained no reportable pathological abnormality.

We determined labels only from information explicitly documented in the report, and did not incorporate symptoms, physical examination findings, laboratory results, treatment information or reinterpretation of the source CT images. When multiple findings were present, we assigned the examination according to the finding with the highest urgency, with Positive taking precedence over non-emergency-positive and negative. Radiologist collaborators reviewed the operational definitions, and we provide detailed criteria for individual abnormalities, temporal qualifiers, negation, uncertain expressions and boundary cases (Supplementary Note 7).

### Image and text preprocessing

We processed all included CT examinations and reports through a fixed pipeline before pretraining. We first reconstructed DICOM series as 3D volumes, resampled them along the z axis to reduce slice-thickness anisotropy, and spatially normalized and reshaped each volume to 32 × 256 × 256 voxels. We removed invalid slices outside the effective head field of view where applicable. We then clipped intensities using fixed head CT windows and mapped them to a common scale, which reduced acquisition-related variation while preserving density information relevant to brain parenchyma, hemorrhage, fracture-adjacent hyperdensity and other emergency findings.

We de-identified and harmonized radiology reports and question-answer texts by text cleaning, terminology normalization, sentence segmentation and structured processing. This procedure removed formatting artifacts and redundant symbols, reduced variation from synonymous medical expressions and abbreviations, and decomposed long reports into more stable semantic units for cross-modal alignment and image-conditioned generation.

### CHIEF architecture and joint pretraining objectives

We designed CHIEF as a unified 3D vision-language pretraining framework for emergency head CT (Extended Data Figs. 2 and 3). The framework learns a shared semantic space by aligning paired CT volumes and radiology reports, while retaining interfaces for image-conditioned report generation and downstream clinical probes. The full pretraining architecture is provided (Extended Data Fig. 2), with additional implementation-level details shown (Extended Data Fig. 3).

This design addresses two challenges that are prominent in emergency head CT. First, radiology reports contain templated phrasing and frequent term co-occurrence, which can encourage superficial image-text matching. Second, multicenter CT data vary in scan protocol, slice thickness, noise level, reconstruction kernel and population structure, creating opportunities for site-specific shortcuts. CHIEF therefore combines four components: bidirectional contrastive alignment with learnable temperature clipping, dual-path latent representations, image-conditioned generation for semantic anchoring and representation-geometry regularization for cross-center robustness.

<u>Bidirectional contrastive alignment based on learnable temperature clipping.</u> Cross-modal alignment serves as the core training signal of the pretraining framework. We adopt a bidirectional symmetric InfoNCE loss with in-batch negative samples to jointly optimize matching in both directions, namely text-to-image retrieval and image-to-text retrieval, within a shared semantic space. Given a batch of paired CT and text samples, the model produces contrastive representations for the image and text modalities, denoted by *u_v_* and *u_t_*, respectively. Both are *d*-dimensional vectors and are L2-normalized before similarity computation, such that the dot product is equivalent to cosine similarity:

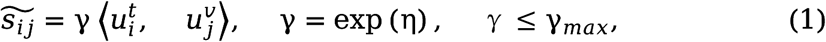

Where <.,.> denotes the vector dot product, η is a learnable scalar parameter, γ is the similarity scaling factor and γ*_m_* is a predefined upper bound.

To improve training stability under multicenter conditions, we use a learnable temperature parameter with upper-bound clipping for similarity scaling. Relative to natural-image data, multicenter clinical batches show greater variation in sample composition and difficulty. If the temperature scale increases without constraint, the similarity distribution may become excessively sharp, leading to undue gradient concentration and training instability. Temperature clipping limits uncontrolled scale amplification while preserving adaptivity, thereby enabling a more stable consistency optimization process on heterogeneous clinical data. On this basis, the contrastive loss is defined as:

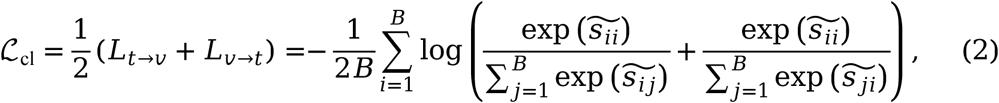

where *L_t_*_→*v*_ denotes the loss in the text-to-image direction and *L_v_*_→*t*_ denotes the loss in the image-to-text direction. This bidirectional alignment mechanism enables the model to establish stable retrieval capability in both directions within the shared semantic space and provides a unified basis for cross-modal alignment in subsequent downstream probe tasks.

<u>Dual-path latent design: decoupling alignment discriminability from downstream usability.</u> During pretraining, although contrastive learning can effectively establish cross-modal consistency, its optimization objective primarily acts on relative similarity structure rather than on the semantic organization required by downstream clinical tasks. If a single latent space is required to simultaneously support alignment, generation and transfer, the resulting representation geometry may become overly optimized for contrastive separability, thereby compromising semantic stability and cross-distribution generalization. To mitigate this conflict, we introduce a dual-path latent design that explicitly decouples alignment discriminability from downstream usability. The visual and text encoders first generate global features, ℎ*_v_* and ℎ*_t_*, respectively, which are then projected through linear mapping and LayerNorm into a base latent space with unified dimensionality:

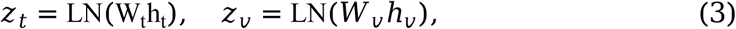

where *z_t_* and *z_v_* serve as general semantic carriers and are used by default for image-conditioned generation and downstream probe tasks. On this basis, we further stack lightweight contrastive heads, *ψ_t_* and *ψ_v_*, on top of the base latent and construct a contrastive latent using nonlinear projection, residual connection, LayerNorm and L2 normalization:

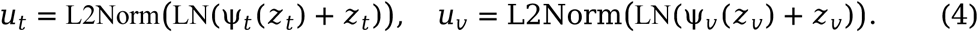

During training, the contrastive loss is applied only to the contrastive branch, (*u_t_*, *u_v_*), whereas downstream tasks and the generation branch operate by default on the base latent, (*z_t_*, *z_v_*). In this way, the framework preserves a smoother, more stable and more transferable semantic structure while maintaining strong alignment capability.

<u>Image-conditioned generation for semantic anchoring.</u> Pure contrastive learning mainly constrains relative relationships among samples, but does not explicitly define the linguistic semantics to which an image should correspond. This limitation is particularly evident in head CT, where radiology reports are highly templated. Under such conditions, the model may achieve pairing by relying on high-frequency phrasing or institution-specific reporting habits, without truly establishing content-level correspondence between image and semantics. To strengthen semantic anchoring, we further introduce an image-conditioned language generation objective during pretraining and use it as an additional anchor for cross-modal representation learning.

Specifically, we map the visual representation *z_v_* of the 3D CT through a projection module *p*_ω_ into a set of continuous prefix embeddings, which are concatenated with the target text token sequence at the input stage. The resulting sequence is then fed into an autoregressive language model to model the conditional probability and compute the generation loss. Let the number of prefix tokens be *K* (with the default implementation setting *K* = 1), and let the hidden dimension of the language model be *H*. The prefix tensor is defined as:

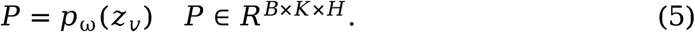

Under this prefix-conditioning mechanism, we define the generation training objective through conditional language modeling. During training, teacher forcing is adopted, and the token-level cross-entropy over the target sequence *y*=(*y*_1_, …,*y_T_*_)_, excluding PAD, is computed as:

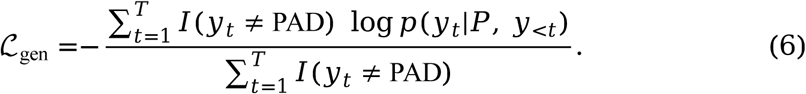

This generation objective provides a complementary constraint to contrastive learning: the model is required not only to retrieve semantically matched text within the shared space, but also to generate condition-consistent language descriptions from the input CT. As a result, the learned representations provide not only cross-modal matching capability, but also stronger linguistic interpretability and semantic groundedness, thereby supporting subsequent report generation and question-answering reasoning.

<u>Representation geometry regularization for cross-center generalization.</u> In addition to alignment and generation, we further improve model robustness under multicenter conditions through representation geometry regularization. In emergency head CT, differences in slice thickness, noise level, reconstruction kernel and scanning settings can readily introduce low-level statistical biases unrelated to clinical semantics into the representation space. These biases may manifest as dimensional redundancy, covariance imbalance and increased embedding anisotropy. As a result, a small number of dominant directions may exert disproportionate influence on similarity computation, thereby weakening cross-institution generalization.

To address this issue, we introduce a decorrelation regularization term to constrain redundant correlations among feature dimensions and encourage different dimensions to encode complementary information. Unlike the contrastive loss, which focuses on pairwise relationships between samples, this regularization acts directly on the statistical geometry of the representation space. By jointly constraining both image and text representations, this design helps reduce reliance on institution-specific acquisition and reporting patterns, while promoting preservation of clinically semantic factors that are more stable across centers.

### Three-dimensional visual and text encoders

<u>Three-dimensional visual encoder.</u> The visual branch uses a 3D encoder to model head CT volumetric data end to end. Compared with approaches that treat CT as a sequence of two-dimensional slices, a 3D encoder can directly capture anatomically continuous structures and lesion morphology across slices in volumetric space, which is better aligned with the imaging characteristics of brain injury. For example, the spatial continuity between hemorrhagic lesions and surrounding edema, the extension of small hemorrhages across adjacent slices, and the geometric consistency between fracture lines and neighboring hyperdense regions are all more appropriately modeled in a 3D setting.

In implementation, we feed the preprocessed 3D CT volumes into CTViT and encode them into dense 3D token representations, which we then globally aggregate to obtain an image-level representation, ℎ_*v*_ This representation serves as the shared visual semantic carrier for subsequent cross-modal alignment, image-conditioned generation and downstream probes. The visual encoder is not designed for any single task; rather, it functions as a shared representation interface within the unified pretraining framework, supporting multiple objectives on the basis of the same visual representations.

<u>Text encoder.</u> The text branch uses a pretrained language model to encode the corresponding radiology reports. Because multicenter clinical texts show systematic variation in writing style, degree of templating, terminology granularity and information density, we adopt a last-four-layer fusion strategy. Specifically, token-level representations are extracted from the final four hidden layers of the language model and fused to achieve a more robust balance between high-level semantic abstraction and word-level detail. The fused token representations are then

pooled to form a sentence-level text representation, ℎ*_t_*, which is subsequently mapped into the shared semantic space for cross-modal loss computation. Compared with single-layer representations, cross-layer fusion is less susceptible to templated language and institution-specific expression patterns, and is better suited to stably capturing entities, attributes and relational information relevant to imaging semantics, thereby reducing the influence of superficial linguistic form on image-text alignment.

<u>Latent space projection and task interfaces.</u> The global representations ℎ*_v_* and ℎ*_t_* produced by the visual and text encoders are first mapped through linear projection and normalization into a base latent space with unified dimensionality, yielding *z_v_*and *z_t_*, respectively. On this basis, we further construct contrastive latents through lightweight contrastive heads, yielding *u_v_* and *u_t_*, respectively. This two-level latent-space interface allows different training objectives to operate on distinct representation branches within a unified framework. Specifically, the contrastive loss is applied only to the contrastive latent to shape the discriminative geometry required for cross-modal alignment. By contrast, the image-conditioned generation branch and downstream probes preferentially use the base latent, thereby preserving a smoother and more robust semantic structure as far as possible. Through this division of task interfaces, the framework balances representation sharing with objective decoupling while maintaining a shared encoder backbone.

<u>Joint pretraining objectives.</u> During pretraining, we adopted a single-stage joint optimization strategy in which three training signals-cross-modal alignment, image-conditioned generation and representation-geometry regularization-were optimized jointly. These objectives shared the same visual and text encoder backbones, but operated on partially distinct latent branches, thereby reducing interference among objectives as far as possible without introducing the complexity of multi-stage training. The overall optimization objective was defined as:

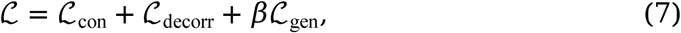

where ℒ_con_ denotes the bidirectional symmetric contrastive learning loss, which learns a shared cross-modal semantic space and provides the core training signal for retrieval capability; ℒ_gen_ denotes the image-conditioned generation loss, which anchors the 3D CT representations to the language generation trajectory; ℒ_decorr_ denotes the decorrelation regularization term, which suppresses dimensional redundancy and improves cross-center robustness from the perspective of representation geometry; and is a hyperparameter controlling the weight of the generation objective. This joint optimization scheme allows contrastive learning, generative constraints and geometric regularization to play complementary roles within a unified framework. As a result, the learned cross-modal representations support not only paired-sample matching, but also the semantic basis required for downstream transfer, while maintaining more consistent performance in real multicenter clinical settings.

### Downstream probe implementation

We formulated all downstream analyses as probes of the same pretrained representation (Extended Data Figs. 4–7 and Supplementary Table 8). This design allowed report generation, triage classification, cross-modal retrieval, CQ500 multi-label classification and zero-shot abnormality detection to be evaluated as reusable interfaces to the pretrained representation rather than as independent task-specific systems.

<u>Image-to-text report generation probe.</u> We used the image-to-text generation probe to evaluate the model’s ability to map imaging semantics into language (Extended Data Fig. 4). This probe reused the image-conditioned generation module introduced during pretraining by projecting the 3D CT representation into a continuous prefix and feeding it into an autoregressive language model to generate text. This setting tested whether the pretrained representation could support stable linguistic expression of imaging semantics without additional architectural redesign.

<u>Emergency triage classification probe.</u> We used the emergency triage classification probe to evaluate the transferability of the pretrained representations in discriminative tasks (Extended Data Fig. 5). In the basic setting, the model used the visual-side base latent as input and performed prediction through a lightweight classification head. We also evaluated generation-feature enhancement, in which hidden states from the image-conditioned language model were projected and fused with the visual representation to support decision-level prediction, particularly for abnormalities with finer semantic granularity.

<u>Cross-modal retrieval probe.</u> We used the cross-modal retrieval probe to evaluate direct transfer of the pretrained representation to image-text semantic matching (Extended Data Fig. 6). Image and text inputs were encoded into base latents, L2-normalized and compared by cosine similarity. No additional generation or classification objective was introduced, so retrieval performance directly reflected the consistency of the learned cross-modal representation space.

<u>CQ500 multi-label classification probe.</u> On the CQ500 dataset, we adopted a fine-tuned multi-label binary classification probe to evaluate the model’s transfer stability under a structured label system involving brain hemorrhage, fracture and related abnormalities. This dataset contains both overall-category labels and multiple fine-grained subtype labels, and each label dimension was modeled as an independent binary classification task. To address class imbalance and long-tail distribution, the classification head used an asymmetric loss, and a hierarchical consistency constraint was introduced during training to reduce medically implausible prediction combinations. To ensure that evaluation focused on transferability of the visual representations themselves, this probe did not introduce label text as input, thereby avoiding prediction through linguistic shortcuts.

<u>Zero-shot abnormality detection probe.</u> We used the zero-shot abnormality detection probe to evaluate abnormality recognition in the absence of explicit task supervision (Extended Data Fig. 7; Supplementary Figs. 13 and 14). For each predefined abnormality category, label-specific question templates were constructed and the image-conditioned language model was used to estimate calibrated yes/no probabilities. Multi-template ensembling, baseline prefix calibration and adaptive thresholding were used to reduce prompt phrasing bias and convert label scores into abnormality predictions.

To clarify how the same pretrained representation was used across downstream tasks, we summarized the input interface, output space and evaluation endpoints for each probe (Supplementary Table 8). The table is intended as an implementation map rather than a performance table; numerical results are reported (Supplementary Tables 1–5), with implementation details, abnormality labels and negation rules summarized (Supplementary Figs. 13 and 14). Abnormality-detection results for the internal zero-shot setting and fine-tuned CQ500 benchmark are reported (Supplementary Tables 6 and 7).

### Evaluation metrics and statistical analysis

We evaluated all downstream tasks using task-appropriate metrics and reported point estimates together with 95% confidence intervals. Unless otherwise stated, we estimated confidence intervals by sample-level bootstrap resampling: in each bootstrap iteration, we resampled the test set with replacement, recomputed the metric, and used the 2.5th and 97.5th percentiles of the bootstrap distribution as confidence bounds.

For image-to-text report generation, we evaluated lexical overlap, sequence-level content preservation and contextual semantic consistency using Corpus-BLEU, ROUGE-L, BERTScore, METEOR and CIDEr. These metrics jointly assess n-gram agreement, longest-common-subsequence overlap, contextual embedding similarity, word-alignment quality and TF-IDF-weighted consensus similarity between generated and reference reports. Full mathematical definitions are provided in Supplementary Note 5.

For emergency triage and other classification probes, we reported the Accuracy, Balanced Accuracy, Precision, Recall, F1-score, Specificity, G-mean, AUROC and Cohen’s Kappa where applicable. For multiclass tasks, class-wise statistics were computed in a one-vs-rest manner and then macro-averaged across classes, which reduced the influence of class-frequency imbalance on aggregate performance.

For cross-modal retrieval, we evaluated ranking quality using Recall@K, NDCG@K and mean reciprocal rank (MRR). Recall@K quantified whether the paired item appeared among the top-ranked candidates, NDCG@K weighted correct matches more strongly when they appeared at higher ranks, and MRR measured the reciprocal rank of the first relevant item. Retrieval metrics were calculated on a per-query basis and summarized across all queries.

For CQ500 multi-label classification and zero-shot abnormality detection, each label was treated as an independent binary endpoint. Threshold-free performance was assessed using AUROC and average precision, while thresholded predictions were summarized using Accuracy, Balanced Accuracy, Precision, Recall, F1-score, Specificity and G-mean. Macro- and micro-averaged summaries were reported where applicable. The abnormality label bank, negation rules and full metric formulae are provided (Supplementary Notes 5 and 6 and Supplementary Figs. 13 and 14).

### Ethics statement

This study received institutional review board approval from all participating centers. Data collection at West China Hospital was approved by its Research Ethics Committee (2025-1612). Data collection at Mianyang Central Hospital was approved by its Research Ethics Committee (S202503234-1). The Research Ethics Committee of Santai County People’s Hospital approved data collection under protocol 2025-KY-0076, while Beichuan Qiang Autonomous County People’s Hospital approved data collection under protocol LL-20250002. Data collection at Tsinghua University Hospital was approved by its Research Ethics Committee (THU-01-2025-1054-R1). Data collection at West China Second University Hospital, Sichuan University was approved by its Medical Research Ethics Committee (2026-029), and data collection at Beijing Huaxin Hospital was approved by its Biomedical Research Ethics Committee (R2026-017-01). All patient data were anonymized and de-identified prior to analysis. Given the retrospective nature of the study, the requirement for informed consent was waived. All procedures were conducted in accordance with the Declaration of Helsinki and relevant ethical guidelines for medical research.

## Data availability

The emergency head CT images, associated Chinese radiology reports, and derived annotations used in this study were collected retrospectively from multiple clinical centers and contain sensitive patient information. Therefore, these data are not publicly available due to patient privacy protection regulations and institutional data governance policies. Researchers seeking access to the data for legitimate scientific and non-commercial research purposes may submit a reasonable request to the corresponding author. All requests will be evaluated on a case-by-case basis and are subject to approval by the Ethics Committees of the participating institutions, including West China Hospital, Mianyang Central Hospital, Santai County People’s Hospital, Beichuan Qiang Autonomous County People’s Hospital, Tsinghua University Hospital, West China Second University Hospital, Sichuan University, and Beijing Huaxin Hospital.

## Code availability

The complete source code for model construction and implementation, including data preprocessing pipelines, model architecture, and evaluation scripts, is available for download through GitHub (https://github.com/JustlfC03/CHIEF). This code repository contains detailed documentation and usage instructions to facilitate reproduction of our results and further development of the models.

## Author contributions

Jingyuan Zheng: Methodology, Writing – original draft. Yifei Chen: Methodology, Conceptualization, Writing – original draft, Writing – review & editing. Beining Wu: Report generation experiments, Visualization. Yuanhan Wang: Methodology, Report generation experiments. Mingxuan Liu: Visualization, Writing – review & editing. Lu Li: Data collection (West China Hospital), Annotation. Shuo Jiang: Retrieval experiments. Weihong Chen: Retrieval experiments. Liaoman Xu: Classification experiments. Yueyi Wu: Classification experiments. Chang Liu: Visualization. Lulu Guo: Visualization. Xuguang Bai: Visualization. Zihan Li: Visualization. Hongjia Yang: Visualization. Feiwei Qin: Writing – review & editing. Jingzhe Liu: Data collection (Beijing Huaxin Hospital), Annotation. Haibo Qu: Data collection (West China Second University Hospital, Sichuan University), Annotation. Qiang Liao: Data collection (Beichuan Qiang Autonomous County People’s Hospital), Annotation. Gang Zhao: Data collection (Santai County People’s Hospital), Annotation. Keqin Pan: Data collection (Tsinghua University Hospital), Annotation. Jun Guo: Data collection (Tsinghua University Hospital), Annotation. Lizhou Chen: *Turing* test. Ying Zhou: Data collection (Mianyang Central Hospital), Annotation. Huaiqiang Sun: Supervision, Writing – review & editing, Supervision, Funding acquisition. Qiyuan Tian: Methodology, Conceptualization, Supervision, Project administration, Funding acquisition, Writing – original draft, Writing – review & editing.

## Acknowledgments

This work was supported by the National Natural Science Foundation of China (grant number 82302166), Dushi Program (grant number 20261080018), and Tsinghua University Startup Fund.

---

**Extended Data Fig. 1.**
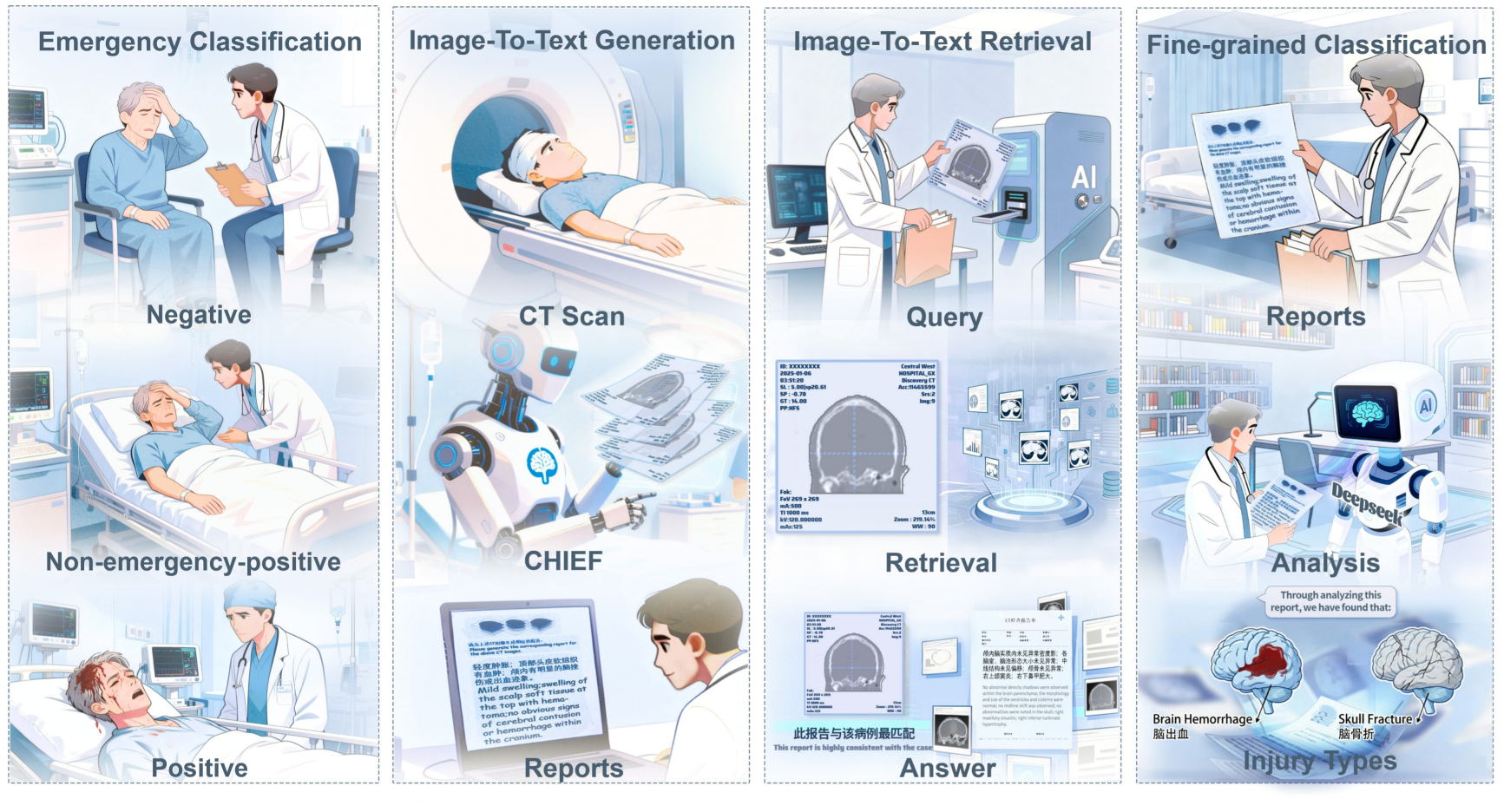
Clinical applications supported by CHIEF across emergency head CT interpretation. CHIEF supports several clinically relevant applications based on head CT examinations and their corresponding radiology reports. In the emergency setting, examinations can be categorized as negative, non-emergency-positive or positive to assist triage and prioritization of care. The same framework can generate radiology-style textual descriptions from 3D head CT scans, retrieve reports from clinically similar cases and support more detailed abnormality-subtype assessment, including intracranial hemorrhage and skull fracture. Together, these applications demonstrate how a single framework can support diagnostic assessment, report generation and case-based retrieval in emergency neuroimaging.

**Extended Data Fig. 2.**
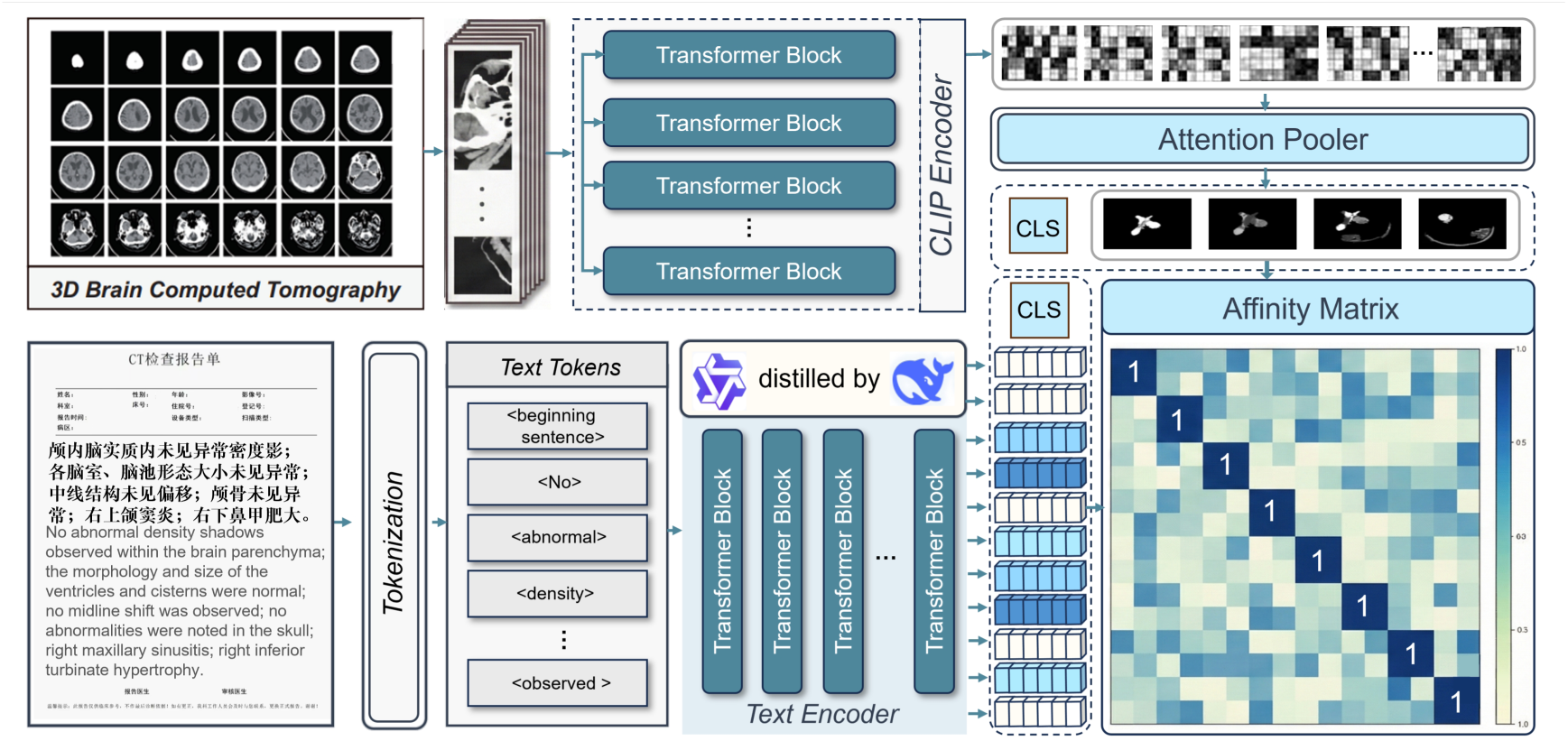
Architecture of CHIEF for joint representation learning from 3D head CT and radiology reports. Three-dimensional non-contrast head CT volumes are encoded by a CTViT visual encoder to produce an image-level visual representation, whereas Chinese radiology reports are encoded by a Chinese BERT text encoder with last-four-layer fusion and report-level pooling. Visual and textual representations are projected into a shared base latent space. Bidirectional contrastive alignment is applied through contrastive heads to shape image-text matching, image-conditioned generative semantic anchoring maps the visual latent into continuous prefix embeddings for report reconstruction, and representation geometry regularization constrains embedding redundancy. This architecture supports report generation, emergency triage classification, cross-modal retrieval and zero-shot abnormality detection through a shared pretrained representation.

**Extended Data Fig. 3.**
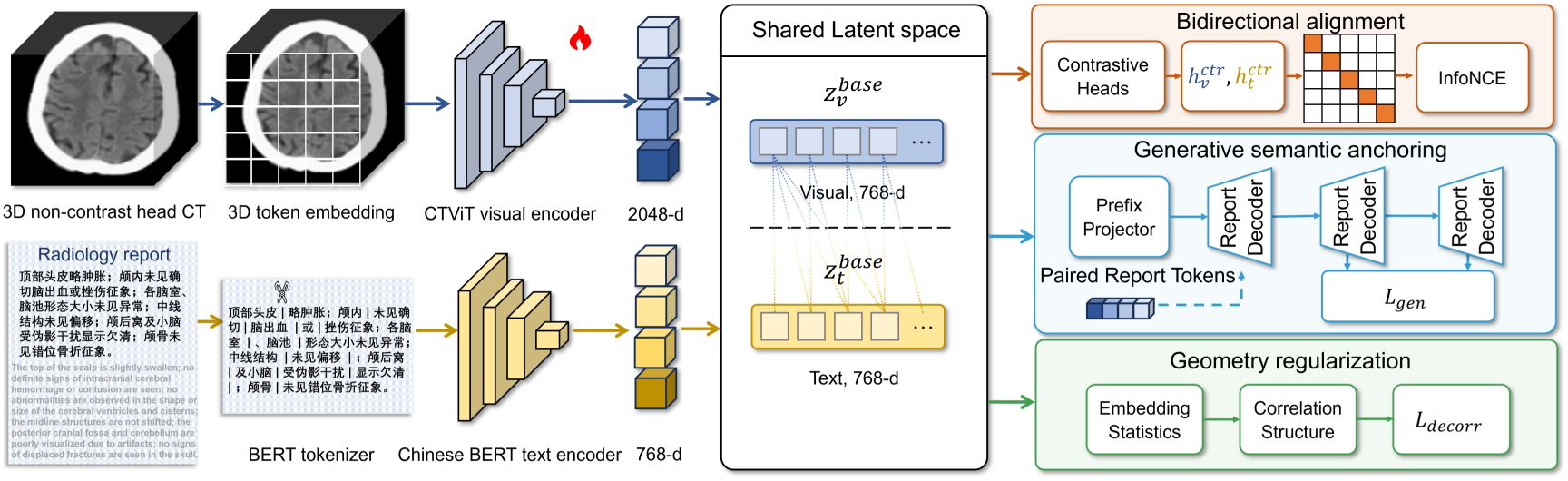
Implementation-level CHIEF architecture for joint representation learning from three-dimensional head CT and radiology reports. Three-dimensional non-contrast head CT volumes are encoded by the CTViT visual encoder to produce image-level visual representations, whereas Chinese radiology reports are encoded by a Chinese BERT text encoder with last-four-layer fusion and report-level pooling. Visual and textual representations are projected into a shared base latent space. Lightweight contrastive heads support bidirectional image-text alignment, the prefix-generation branch supports image-conditioned semantic anchoring, and representation-geometry regularization constrains redundant embedding structure.

**Extended Data Fig. 4.**
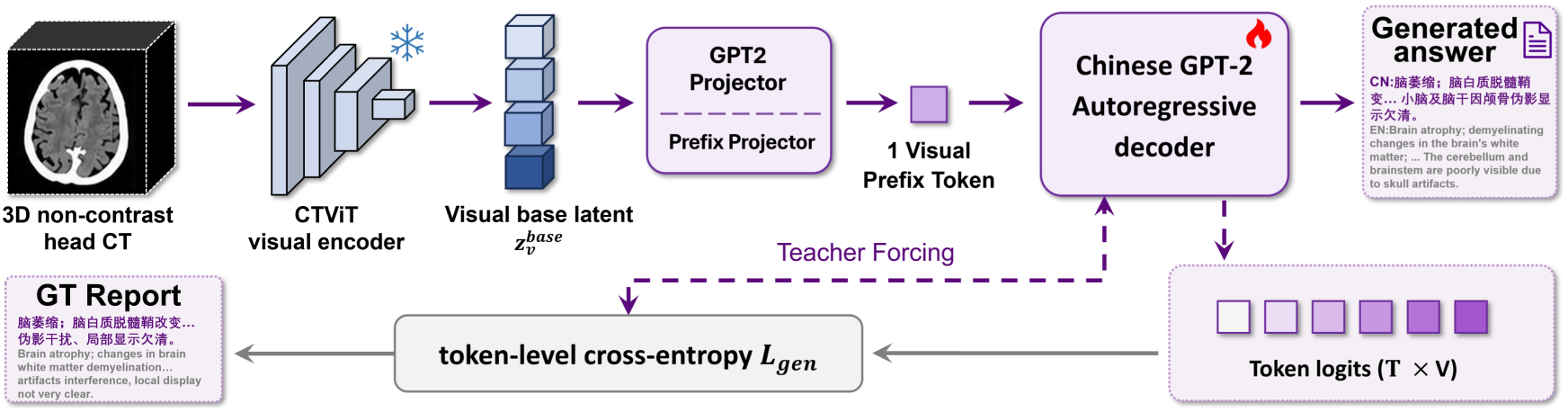
Image-to-text report-generation probe. A three-dimensional head CT volume is encoded by the pretrained CTViT visual encoder, projected into the visual base latent and mapped to continuous prefix embeddings. The Chinese GPT-2 decoder then generates the radiology report autoregressively, with teacher forcing and token-level cross-entropy used during training.

**Extended Data Fig. 5.**
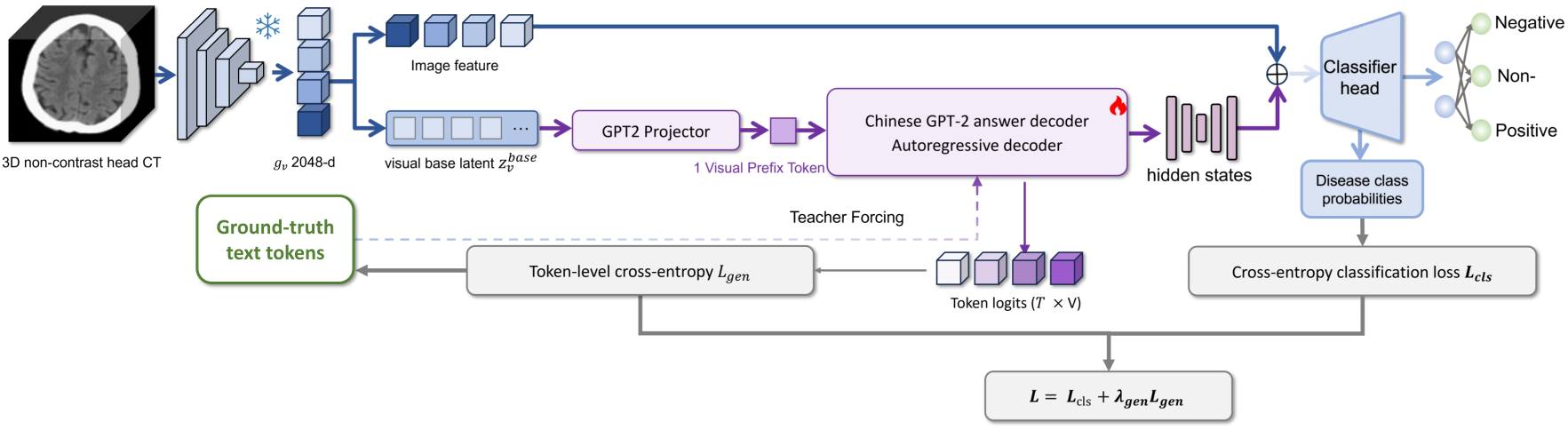
Emergency triage classification probe. The visual base latent from the pretrained CHIEF backbone is used by a lightweight classification head to predict negative, non-emergency-positive and positive examinations. Generation-derived hidden states can be projected and fused with the visual representation as auxiliary semantic features, and the probe is optimized using classification loss with optional generation-feature supervision.

**Extended Data Fig. 6.**
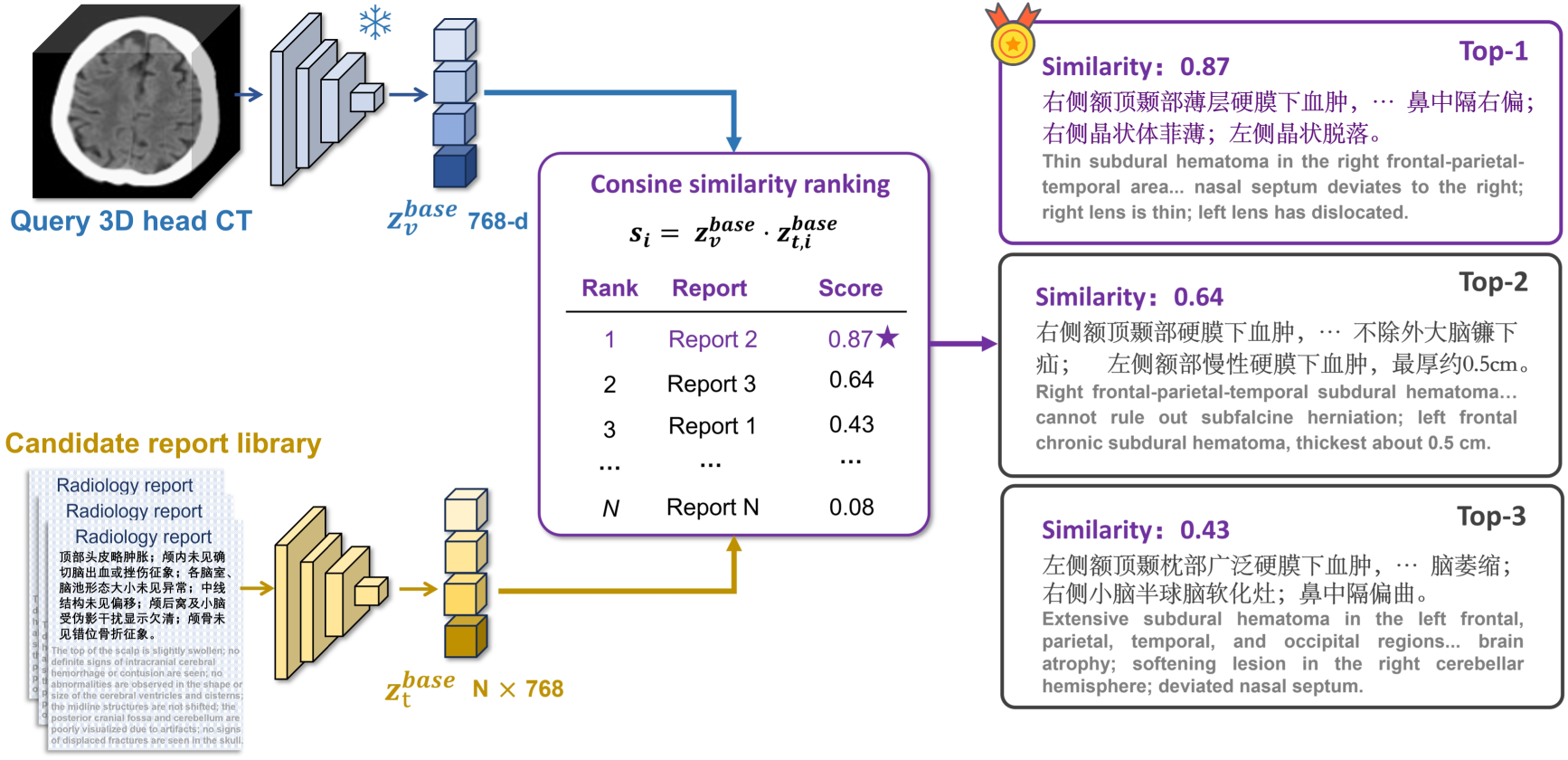
Cross-modal retrieval probe. Query CT volumes and candidate reports are encoded into L2-normalized image and text base latents. Reports are ranked by cosine similarity to the query CT representation, enabling retrieval of clinically similar reports from the candidate report library without adding a separate task-specific training objective.

**Extended Data Fig. 7.**
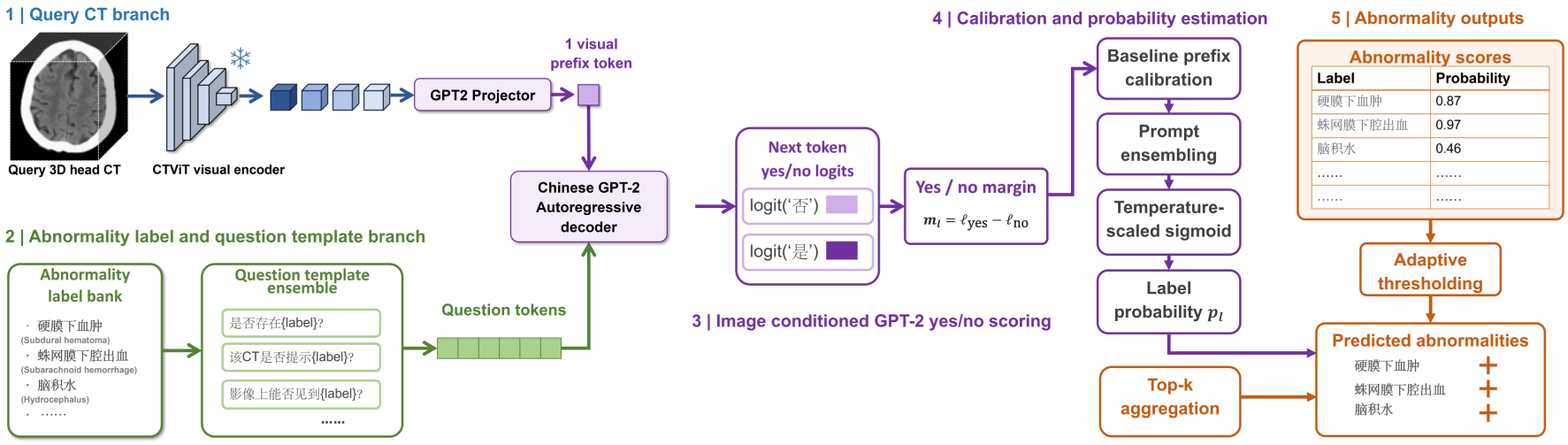
Zero-shot abnormality-detection probe. A query CT volume is encoded into the visual base latent and converted to a visual prefix for the image-conditioned decoder. Abnormality labels are inserted into Chinese question-template ensembles, and next-token yes/no logits are converted into calibrated label probabilities using baseline prefix calibration, prompt ensembling and temperature-scaled sigmoid transformation. Adaptive thresholding and top-k aggregation are then used to obtain abnormality-level outputs.

**Extended Data Fig. 8.**
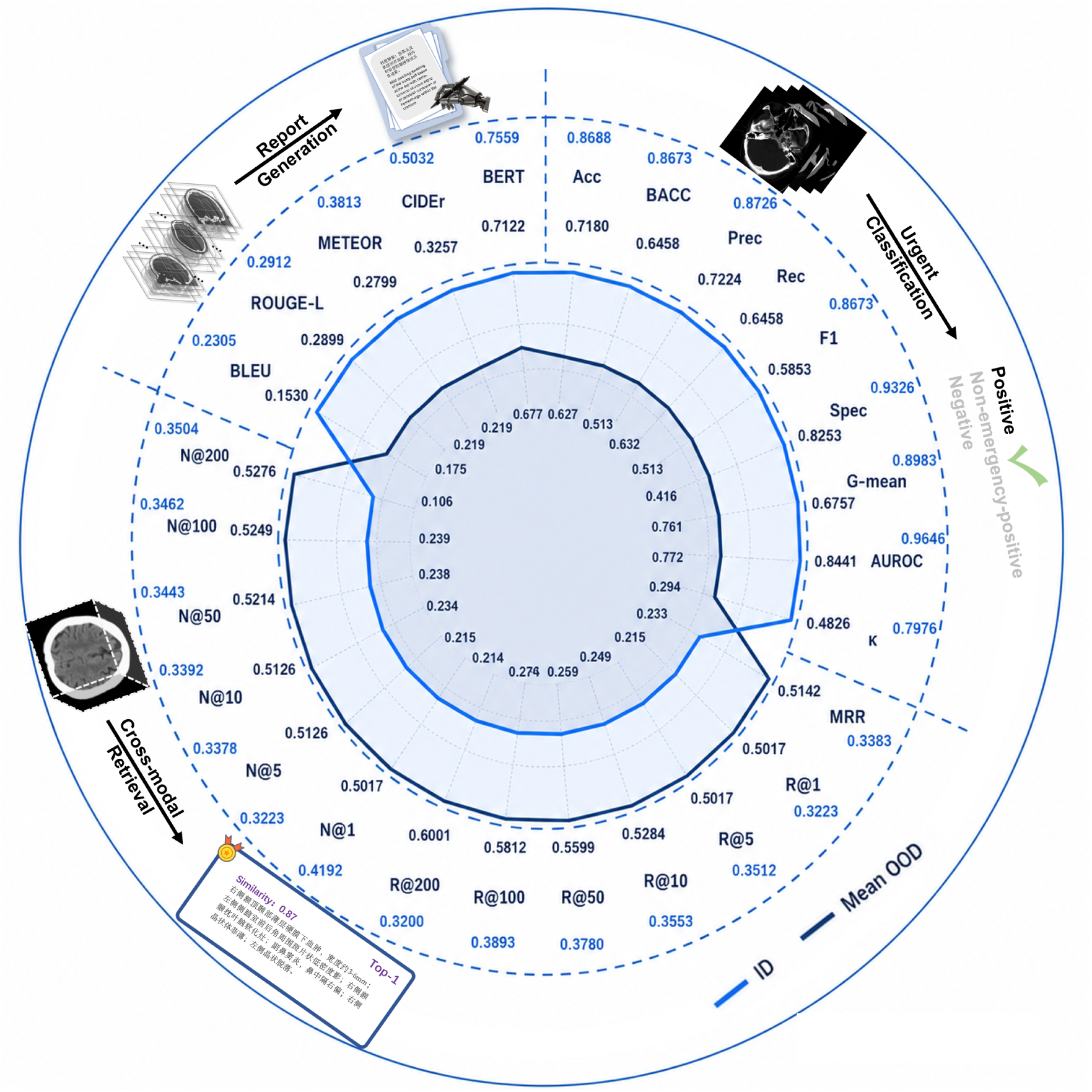
Cross-center performance degradation across downstream tasks. Relative changes from the internal cohort to external cohorts are summarized for report generation, emergency triage classification and image-to-text retrieval. Metrics are normalized within each task to enable comparison across heterogeneous endpoints, and the panels summarize internal versus mean external performance, relative performance drop, worst-center performance and task-level normalized score profiles.

