## Supplementary Information for "A multimodal foundation model for emergency head CT interpretation"

### Supplementary Notes

#### Supplementary Note 1 | Additional cohort and quality-control details

The main cohort construction, institution-stratified in-distribution and out-of-distribution design, and CQ500 validation setting are described in Online Methods. Here, we provide supplementary quality-control information supporting the cohort definition. All included examinations were required to have complete non-contrast head CT volumetric coverage, reliable DICOM metadata, stable HU calibration and one-to-one linkage to the corresponding radiology report from the same clinical encounter. Cases were excluded when imaging quality, metadata integrity or report–image correspondence was insufficient to support examination-level weak supervision. These criteria were used to reduce semantic noise and maintain consistent paired supervision across centres.

#### Supplementary Note 2 | Additional preprocessing details

The primary image and text preprocessing pipeline is described in Online Methods. In brief, all CT volumes were standardised to a fixed volumetric input size and intensity scale before model training, and all reports were de-identified and normalised before cross-modal pairing. Supplementary preprocessing checks were used to remove invalid slices, non-head series, incomplete series and report texts with obvious semantic contamination. These procedures were applied before cohort splitting to ensure consistent image–text supervision across centres.

#### Supplementary Note 3 | Additional architecture implementation details

The CHIEF architecture and joint pretraining objectives are described in Online Methods and summarized (Extended Data Fig. 2). Additional implementation-level schematics of the backbone and downstream task interfaces are provided (Extended Data Figs. 3–7), and cross-centre performance-degradation analyses are provided (Extended Data Fig. 8). Briefly, the visual branch encodes preprocessed three-dimensional head CT volumes into image-level representations, the text branch encodes Chinese radiology reports into report-level representations, and the two modalities are connected through base and contrastive latent interfaces. The implementation separates representation interfaces used for contrastive alignment from those used for generation and downstream probes to reduce interference among objectives. Detailed downstream input–output interfaces are summarized (Supplementary Table 8).

#### Supplementary Note 4 | Downstream probe implementation summary

Downstream analyses were implemented as lightweight probes of the same pretrained CHIEF representation rather than as independent task-specific systems. The input interface, output space and evaluation endpoints for report generation, emergency triage classification, image-to-text retrieval, fine-tuned CQ500 multilabel classification and internal zero-shot abnormality detection are summarized (Supplementary Table 8). Numerical results for report generation, triage classification and retrieval are reported (Supplementary Tables 1–5); abnormality-detection results for the internal zero-shot setting and the fine-tuned CQ500 benchmark are reported (Supplementary Tables 6 and 7).

#### Supplementary Note 5 | Full evaluation metrics and statistical analysis

All downstream tasks were evaluated using task-appropriate metrics, with point estimates reported together with 95% confidence intervals. Unless otherwise stated, confidence intervals were estimated by sample-level bootstrap resampling. For each bootstrap iteration, the test set was sampled with replacement, the metric was recomputed and the 2.5th and 97.5th percentiles of the bootstrap distribution were used as the confidence bounds.

<u>Report generation evaluation metrics.</u> Corpus-BLEU was used to quantify the overall n-gram-level agreement between generated reports and reference reports. Unlike approaches that compute BLEU for each sample and then average across samples, corpus-BLEU first aggregates the modified precision at each n-gram order over the entire corpus, then combines these values using a weighted geometric mean, with a brevity penalty to suppress spuriously inflated scores arising from overly short candidate texts:

$$BLEU_{corpus} = BP \cdot exp\left(\sum_{n=1}^{N} w_n log\, p_n\right), \quad BP = min\big(1, exp(1 - r/\mathrm{c})\big),$$

$$p_n = \frac{\sum_{g \in \mathcal{G}_n} \min\left(count_{cand}(g), count_{ref}(g)\right)}{\sum_{g \in \mathcal{G}_n} count_{cand}(g)}, \tag{1}$$

where $G_n$ denotes the set of all n-grams appearing in the candidate corpus; $\mathrm{count_{cand}}(g)$ denotes the number of occurrences of n-gram $g$ in the candidate corpus; $\mathrm{count_{ref}}(g)$ denotes the number of occurrences of $g$ in the reference corpus, with the maximum count across references used in the multiple-reference setting; and $c$and $r$denote the total token lengths of the candidate corpus and reference corpus, respectively.

ROUGE-L measures the overlap between candidate text and reference text based on the longest common subsequence, with greater emphasis on content preservation and sequence-order consistency. Its F-form is defined as:

$$ROUGE\text{-}L_F = \frac{(1+\beta^2)\, P_{LCS}\, R_{LCS}}{R_{LCS} + \beta^2 P_{LCS}}, \quad P_{LCS} = \frac{LCS(X,Y)}{|X|}, \quad R_{LCS} = \frac{LCS(X,Y)}{|Y|}, \qquad (2)$$

where, $\mathrm{LCS}(X, Y)$ denotes the length of the longest common subsequence between candidate text $X$ and reference text $Y$; $|X|$ and $|Y|$ denote the corresponding sequence lengths; and $\beta$ is typically set to 1 to balance precision and recall.

BERTScore was used to assess semantic consistency between generated text and reference text in contextual semantic space. Let the contextual embeddings of tokens in the candidate text and reference text be denoted by $e_x$ and $e_y$, respectively. Precision, recall, and the F-score are defined as:

$$P = \frac{1}{|X|}\sum_{x\in X} \max_{y\in Y} \cos(e_x, e_y), \quad R = \frac{1}{|Y|}\sum_{y\in Y} \max_{x\in X} \cos(e_x, e_y), \quad F = \frac{2PR}{P+R}. \qquad (3)$$

METEOR jointly considers precision and recall through word-level alignment and further introduces a word-order penalty term to reduce superficially high similarity driven by non-contiguous matches. It is defined as:

$$METEOR = (1 - Pen) \cdot \frac{PR}{\alpha P + (1-\alpha)R}, \quad Pen = \gamma \left(\frac{ch}{m}\right)^{\theta}, \qquad (4)$$

where, Pendenotes the word-order penalty term, which penalizes non-contiguous distributions of matched tokens between candidate text and reference text, thereby reducing scores for generated results with disordered word order but superficially similar content. In this formulation, $m$denotes the number of matched tokens, $ch$denotes the number of contiguous matched chunks, and $\alpha$, $\gamma$, and $\theta$ are hyperparameters.

CIDEr measures the consistency between candidate text and reference text, potentially under a multiple-reference setting, using TF-IDF-weighted n-gram similarity, with greater emphasis on descriptions consistent with corpus-level consensus. For the $n$-th order n-gram, similarity is computed as the cosine similarity between vector representations, and the final CIDEr score is obtained by averaging across n-gram orders:

$$CIDEr = \frac{1}{N}\sum_{n=1}^{N}\frac{g_n(c)\cdot g_n(s)}{|g_n(c)|\,|g_n(s)|}, \quad (5)$$

where, $g_n(\cdot)$ denotes the TF-IDF-weighted n-gram vector representation, $c$ denotes the candidate text, and $s$denotes the set of reference texts.

Evaluation metrics for classification tasks. For classification tasks, we used Precision, Recall, F1-score, Accuracy (ACC), and Balanced Accuracy (BACC) to provide a comprehensive evaluation of model performance from multiple perspectives, including prediction reliability, class coverage, and overall class balance. For multiclass tasks, class-wise statistics were computed in a one-vs-rest manner and then macro-averaged across classes. Specifically:

(1) Macro-Precision. Macro-Precision is calculated by first computing precision for each class and then taking the arithmetic mean across all classes. It reflects the overall reliability of the model's positive predictions across classes without being influenced by class sample distribution. For class $c$, precision and Macro-Precision is defined as:

$$Precision_c = \frac{TP_c}{TP_c + FP_c}, \quad Macro\text{-}P = \frac{1}{C}\sum_{c=1}^{C} Precision_c\,, \quad (6)$$

where $TP_c$ denotes the number of samples correctly predicted as class $c$, and $FP_c$ denotes the number of samples incorrectly predicted as class $c$.

(2) Macro-Recall. Macro-Recall is obtained by averaging recall across classes and reflects the overall ability of the model to detect different classes. For class $c$, recall and Macro-Recall is defined as:

$$Recall_c = \frac{TP_c}{TP_c + FN_c}, \quad \text{Macro-}R = \frac{1}{C}\sum_{c=1}^{C} Recal\,l_c, \quad (7)$$

where $TP_c$ denotes the number of samples correctly predicted as class $c$, and $FN_c$ denotes the number of samples from class $c$ that were incorrectly predicted as other classes.

(3) Macro-F1 score. The Macro-F1 score jointly reflects prediction precision and class coverage across classes. For a given class $c$, the per-class F1 score and overall Macro-F1 score are defined respectively as:

$$F1 = \frac{2PR}{P+R}, \quad \text{Macro-F1} = \frac{1}{C}\sum_{c=1}^{C} F\,1_c. \tag{8}$$

(4) Accuracy (ACC). Accuracy measures the proportion of correctly predicted samples among all samples. It is defined as:

$$ACC = \frac{1}{N}\sum_{i=1}^{N} I(\hat{y}_i = y_i), \tag{9}$$

where $N$denotes the total number of samples, $\hat{y}_i$ and $y_i$ denote the predicted and true labels of the $i$-th sample, respectively, and $I(\cdot)$ is the indicator function.

(5) Balanced Accuracy (BACC). For imbalanced datasets, Balanced Accuracy is defined as the average recall across all classes and can better reflect model performance on each class. In the multiclass setting, Balanced Accuracy is defined as:

$$BACC = \frac{1}{C}\sum_{c=1}^{C} Recall_c. \tag{10}$$

(6) Specificity. Specificity measures the model's ability to correctly identify negative samples. Under the one-vs-rest binary perspective for class $c$, specificity is defined as:

$$Specificity_c = \frac{TN_c}{TN_c + FP_c}, \tag{11}$$

where $TN_c$ denotes the number of samples that truly do not belong to class $c$and are also predicted as not belonging to class $c$.

(7) G-mean. G-mean jointly captures the model's ability to recognize both positive and negative samples. It is defined as the geometric mean of sensitivity (Sensitivity, that is, Recall) and specificity (Specificity), and reflects the overall balance of model performance across classes. For class $c$, the per-class G-mean score and overall Macro-G-mean score are defined respectively as:

$$G\text{-}mean_c = \sqrt{Recall_c \cdot Specificity_c}, \quad \text{Macro-G-mean} = \frac{1}{C}\sum_{c=1}^{C} G\text{-}mean_c. \tag{12}$$

(8) AUROC. For multiclass tasks, we adopted a one-vs-rest strategy by first calculating the AUROC for each class and then taking the macro average across all classes for which AUROC was definable. This metric reflects the overall discriminative ability of the model across classes without being affected by class sample distribution. For class $c$, AUROC and Macro-AUROC is defined as:

$$AUROC_c = \int_0^1 TPR_c\left(FPR_c^{-1}(x)\right) dx, \quad \text{Macro-AUROC} = \frac{1}{|C^*|}\sum_{c\in C^*} AUROC_c, \tag{13}$$

where $\text{TPR}_c$ denotes the true positive rate of class $c$ under different decision thresholds, and $\text{FPR}_c$ denotes the false positive rate of class $c$ under different decision thresholds. $C^*$ denotes the set of classes for which AUROC is definable, that is, classes for which both positive and negative samples are present in the test set, and $|C^*|$ denotes the number of classes in this set. If a class lacks either positive or negative samples in the test set, the AUROC for that class is undefined and is excluded from the macro average.

(9) Cohen's Kappa. Cohen's Kappa was used to measure agreement between predictions and true labels while correcting for chance agreement. It is defined as:

$$\kappa = \frac{p_o - p_e}{1 - p_e}, \tag{14}$$

where $p_o$denotes the observed agreement rate and $p_e$denotes the agreement rate expected by chance.

Evaluation metrics for retrieval tasks. For retrieval tasks, we evaluated the ranking quality of relevant items within the candidate set on a per-query basis and reported Recall@K, NDCG@K, and MRR. Let  denote the rank position of the first relevant item in the candidate list for the -th query. Then:

(1) Recall@K. Recall@K measures the proportion of queries for which at least one relevant item is retrieved within the top $K$ results:

$$Recall@K = \frac{1}{|Q|}\sum_{i=1}^{|Q|} \mathrm{o}I(r_i \leq K).\#(15) \tag{15}$$

(2) NDCG@K. NDCG@K is a ranking metric that assigns greater weight to relevant items appearing at higher ranks. Let DCG@K be defined as:

$$DCG@K = \sum_{j=1}^{K} \frac{rel_j}{log_{2(j+1)}}, \quad NDCG@K = \frac{DCG@K}{IDCG@K}, \tag{16}$$

where $\mathrm{rel}_j$ denotes the relevance of the item at rank $j$ (binary relevance in this study), and $\mathrm{IDCG}@K$ denotes the ideal $\mathrm{DCG}@K$ under the optimal ranking.

(3) MRR (Mean Reciprocal Rank). MRR is defined as the average reciprocal rank of the first relevant item:

$$N = |Q|, \quad MRR = \frac{1}{N}\sum_{i=1}^{N} \frac{1}{r_i}. \tag{17}$$

In this study, retrieval relevance was defined at the examination-pair level: for each CT query, the paired report from the same examination was assigned binary relevance 1 and all other candidate reports were assigned relevance 0. No containment or fuzzy text matching was used for the quantitative retrieval metrics. Retrieval metrics were evaluated using query-level bootstrap resampling to obtain 95% confidence intervals.

<u>Evaluation metrics for CQ500 multilabel classification.</u> CQ500 was formulated as a multilabel classification task. For each label, AUROC and AP were computed on the basis of the ground-truth label and the predicted probability, and both macro- and micro-level summary results were reported. Per-class AUROC denotes the area under the ROC curve obtained by treating each label as an independent binary classification problem; this metric is undefined when a label contains only

positive samples or only negative samples in the test set. Per-class AP denotes the average precision under the corresponding precision-recall curve and is defined as:

$$AP_c = \sum_n (R_n - R_{n-1}) P_n, \tag{18}$$

where $P_n$ and $R_n$ denote the precision and recall at the $n$-th threshold, respectively. Macro-average metrics were defined as follows:

$$\text{Macro-AUROC} = \frac{1}{C}\sum_{c=1}^{C} AURO\,C_c, \quad \text{Macro-AP} = \frac{1}{C}\sum_{c=1}^{C} A\,P_c. \tag{19}$$

For micro-averaging, AUROC and AP were computed after pooling all predictions at the sample-label pair level. All statistics were reported with 95% confidence intervals obtained by bootstrap resampling.

<u>Evaluation metrics for abnormality detection.</u> Abnormality detection was evaluated at two levels: overall abnormality (binary classification) and fine-grained abnormality labels (multilabel classification). In the overall abnormality task, “negative/normal” was coded as 0, and any other abnormality was coded as 1. In the fine-grained task, each abnormality label was evaluated separately in a one-vs-rest binary classification manner.

AUROC and AP were used to evaluate probability-output performance across decision thresholds. After thresholding, Accuracy, Balanced Accuracy, Precision, Recall, F1 score, Specificity and G-mean were reported to summarize overall discrimination, class-balanced performance, abnormality sensitivity and normal-case exclusion. Fine-grained abnormality detection was evaluated independently for each label in a one-vs-rest manner, with macro-average results reported where applicable. Negation rules were applied during report-derived label construction to exclude expressions such as “not seen,” “absent” and “normal,” reducing false-positive labels. All confidence intervals were estimated by bootstrap resampling.

#### Supplementary Note 6 | Additional examples and abnormality-detection reporting

Additional qualitative and robustness displays are provided (Supplementary Figs. 1-15). Class-wise emergency triage performance across the internal and external cohorts is presented in (Supplementary Fig. 1). Additional report-generation examples across representative internal and external clinical scenarios are provided (Supplementary Figs. 2–6), and the comparison with commercial multimodal large language models is shown (Supplementary Fig. 7). Additional image-to-text retrieval examples are shown (Supplementary Figs. 8-12). The abnormality label bank and negation expressions used for report-derived label construction are provided in (Supplementary Figs. 13 and 14), respectively. Representative CT examinations from the internal and external cohorts are illustrated (Supplementary Fig. 15).

#### Supplementary Note 7 | Operational definitions for emergency triage labels

Emergency triage labels were assigned at the examination level from the final non-contrast head CT radiology report. The labels reflected the urgency of the abnormalities documented in the report rather than the overall severity of the patient's clinical condition, final clinical diagnosis, hospital admission or need for surgery. Annotators considered both the imaging findings and diagnostic impression but did not use symptoms, physical examination findings, laboratory results, treatment information or additional findings inferred from the source images. When more than one abnormality was reported, the examination was assigned to the category with the highest urgency: Positive took precedence over non-emergency-positive, which took precedence over negative.

Positive. An examination was labelled Positive when the report described or raised substantial suspicion of an acute, recent, progressive or potentially rapidly deteriorating abnormality that would generally require immediate clinical evaluation, monitoring, additional emergency investigation, specialist consultation or treatment. This category included acute or subacute intracranial haemorrhage; acute or subacute cerebral infarction or other acute stroke signs; acute skull fracture; pathological midline shift; cerebral herniation; substantial mass effect; marked cerebral oedema; and acute or obstructive hydrocephalus. Acute cerebral contusion, traumatic intracranial air, penetrating injury, severe intracranial infection, acute intracranial vascular abnormalities and acute postoperative complications were also assigned to this category. Intracranial masses were classified as Positive when accompanied by haemorrhage, marked oedema, obstructive hydrocephalus, midline shift or risk of herniation.

No fixed threshold of midline displacement was required; a pathological displacement described in the report as mass effect or a high-risk finding was sufficient. New or acute hydrocephalus included ventricular enlargement, progressive ventricular dilatation, periventricular transependymal fluid or shunt dysfunction accompanied by ventricular enlargement or another acute abnormality. Ventricular compression or collapse was treated as a manifestation of mass effect or cerebral oedema rather than as a feature of hydrocephalus.

Expressions such as “suggestive of”, “consistent with”, “indicating” or “highly suspicious for” were treated as indicating that the abnormality was present. Potentially time-sensitive abnormalities described as “cannot be excluded”, “suspicious” or “requiring further exclusion” were also assigned to the Positive category unless the report explicitly favoured artefact or a normal anatomical variant and excluded an acute lesion.

Non-emergency-positive. An examination was labelled non-emergency-positive when the report documented an imaging abnormality that was chronic, old, stable, degenerative, mild or incidental and did not contain acute haemorrhage, acute infarction, acute fracture, substantial mass effect, acute hydrocephalus or another finding requiring immediate management. Typical findings included cerebral atrophy, chronic white-matter ischaemic change, old cerebral infarction, lacunar infarction, encephalomalacia, old haemorrhage or haemosiderin deposition, stable postoperative changes, intracranial atherosclerosis and vascular calcification.

Incidental intracranial lesions without substantial oedema, mass effect, hydrocephalus or acute haemorrhage, including small meningiomas and benign cystic lesions, were also included. Stable chronic subdural fluid collections without mass effect, isolated scalp haematoma or soft-tissue swelling without skull fracture or intracranial injury, and uncomplicated sinus, middle-ear or mastoid inflammatory changes were assigned to this category. A report stating “no acute intracranial abnormality” was not considered negative when a chronic or incidental abnormality was also documented. A lesion otherwise meeting the non-emergency-positive definition was upgraded to Positive when accompanied by acute haemorrhage, marked oedema, pathological midline shift, obstructive hydrocephalus, herniation or another high-risk feature.

Negative. An examination was labelled negative only when the report contained no reportable pathological intracranial, skull or related head abnormality. This category indicated the absence of a pathological abnormality identified on the non-contrast head CT examination and did not imply that the patient was clinically healthy or that abnormalities not detectable on CT had been excluded. Physiological calcification, normal anatomical variants and age-appropriate findings not regarded as pathological by the reporting radiologist were permitted within this category.

Negation, temporal qualifiers and multiple findings. Explicitly negated findings, such as “no intracranial haemorrhage”, “no fracture” or “no midline shift”, were not treated as abnormalities. Temporal qualifiers including “old”, “chronic”, “stable” and “unchanged from the previous examination” were considered when determining urgency. Chronic or stable abnormalities were generally classified as non-emergency-positive, but the presence of any new acute component or high-risk feature resulted in classification as Positive. For example, cerebral atrophy accompanied by an acute subdural haematoma was labelled Positive; an old cerebral infarct accompanied by sinusitis was labelled non-emergency-positive; and a report containing only a normal anatomical variant was labelled negative. These operational definitions were reviewed by radiologist collaborators to confirm consistency with the intended emergency triage categories.

### Supplementary Tables

| Data Source | Method | Accuracy↑ | BACC↑ | Precision↑ | F1↑ | Specificity↑ | G-mean↑ | AUROC↑ | Kappa↑ |
|---|---|---|---|---|---|---|---|---|---|
| | CT-CLIP | 0.4225 [0.391, 0.454] | 0.3972 [0.367, 0.429] | 0.3981 [0.367, 0.430] | 0.3971 [0.366, 0.429] | 0.7005 [0.684, 0.716] | 0.5175 [0.490, 0.546] | 0.5856 [0.558, 0.613] | 0.1009 [0.054, 0.146] |
| | Med-BLIP | 0.4207 [0.390, 0.451] | 0.3469 [0.326, 0.368] | 0.4035 [0.370, 0.438] | 0.2936 [0.272, 0.315] | 0.6764 [0.663, 0.690] | 0.3142 [0.292, 0.336] | 0.5711 [0.544, 0.598] | 0.0285 [-0.012, 0.070] |
| | Merlin | 0.4219 [0.392, 0.452] | 0.3333 [0.333, 0.333] | 0.4219 [0.392, 0.452] | 0.1977 [0.188, 0.208] | 0.6667 [0.667, 0.667] | 0.0000 [0.000, 0.000] | 0.5373 [0.511, 0.562] | 0.0000 [0.000, 0.000] |
| | PubMedCLIP | 0.4219 [0.392, 0.452] | 0.3333 [0.333, 0.333] | 0.4219 [0.392, 0.452] | 0.1977 [0.188, 0.208] | 0.6667 [0.667, 0.667] | 0.0000 [0.000, 0.000] | 0.5008 [0.495, 0.506] | 0.0000 [0.000, 0.000] |
| | **Ours** | **0.8688** [0.846, 0.889] | **0.8673** [0.843, 0.889] | **0.8726** [0.850, 0.893] | **0.8673** [0.844, 0.889] | **0.9326** [0.920, 0.943] | **0.8983** [0.879, 0.915] | **0.9646** [0.955, 0.973] | **0.7970** [0.761, 0.829] |
| | *p-value* | *0.0005* | *0.0005* | *0.0005* | *0.0005* | *0.0005* | *0.0005* | *0.0005* | *0.0005* |
| Out of Distribution 1 | CT-CLIP | 0.3995 [0.357, 0.444] | 0.3793 [0.336, 0.421] | 0.3832 [0.338, 0.426] | 0.3756 [0.332, 0.419] | 0.6922 [0.671, 0.713] | 0.4997 [0.459, 0.538] | 0.5541 [0.514, 0.591] | 0.0732 [0.010, 0.13] |
| | Med-BLIP | 0.3555 [0.312, 0.397] | 0.3448 [0.320, 0.370] | 0.4074 [0.350, 0.461] | 0.2500 [0.220, 0.281] | 0.6739 [0.658, 0.690] | 0.2677 [0.237, 0.298] | 0.5028 [0.464, 0.540] | 0.0201 [-0.021, 0.064] |
| | Merlin | 0.3190 [0.278, 0.359] | 0.3333 [0.333, 0.333] | 0.3190 [0.278, 0.359] | 0.1611 [0.145, 0.176] | 0.6667 [0.667, 0.667] | 0.0000 [0.000, 0.000] | 0.4956 [0.460, 0.534] | 0.0000 [0.000, 0.000] |
| | PubMedCLIP | 0.3190 [0.278, 0.359] | 0.3333 [0.333, 0.333] | 0.3190 [0.278, 0.359] | 0.1611 [0.145, 0.176] | 0.6667 [0.667, 0.667] | 0.0000 [0.000, 0.000] | 0.5027 [0.496, 0.511] | 0.0000 [0.000, 0.000] |
| | **Ours** | **0.6278** [0.584, 0.668] | **0.6945** [0.663, 0.724] | **0.7131** [0.674, 0.748] | **0.6267** [0.587, 0.665] | **0.8206** [0.802, 0.837] | **0.7161** [0.685, 0.745] | **0.8517** [0.827, 0.876] | **0.4578** [0.405, 0.510] |
| | *p-value* | *0.0005* | *0.0005* | *0.0005* | *0.0005* | *0.0005* | *0.0005* | *0.0005* | *0.0005* |
| Out of Distribution 2 | CT-CLIP | 0.4581 [0.415, 0.502] | 0.4484 [0.373, 0.523] | 0.3929 [0.352, 0.437] | 0.3811 [0.336, 0.428] | 0.6990 [0.672, 0.725] | 0.5497 [0.483, 0.606] | 0.5830 [0.530, 0.638] | 0.0933 [0.024, 0.166] |
| | Med-BLIP | 0.5104 [0.467, 0.554] | 0.3429 [0.326, 0.359] | 0.5343 [0.452, 0.616] | 0.2734 [0.248, 0.301] | 0.6751 [0.659, 0.690] | 0.1949 [0.155, 0.235] | 0.5379 [0.486, 0.588] | 0.0267 [-0.021, 0.075] |
| | Merlin | 0.4534 [0.409, 0.498] | 0.3333 [0.333, 0.333] | 0.4534 [0.409, 0.498] | 0.2079 [0.194, 0.222] | 0.6667 [0.667, 0.667] | 0.0000 [0.000, 0.000] | 0.5468 [0.488, 0.604] | 0.0000 [0.000, 0.000] |
| | PubMedCLIP | 0.5006 [0.456, 0.544] | 0.3333 [0.333, 0.333] | 0.5006 [0.456, 0.544] | 0.2223 [0.209, 0.235] | 0.6667 [0.667, 0.667] | 0.0000 [0.000, 0.000] | 0.4885 [0.466, 0.505] | 0.0000 [0.000, 0.000] |
| | **Ours** | **0.8167** [0.782, 0.850] | **0.7021** [0.630, 0.773] | **0.8223** [0.734, 0.889] | **0.7382** [0.658, 0.808] | **0.8846** [0.863, 0.906] | **0.7723** [0.708, 0.828] | **0.9173** [0.894, 0.939] | **0.6558** [0.593, 0.719] |
| | *p-value* | *0.0005* | *0.0005* | *0.0005* | *0.0005* | *0.0005* | *0.0005* | *0.0005* | *0.0005* |
| Out of Distribution 3 | CT-CLIP | 0.2948 [0.255, 0.337] | 0.3783 [0.248, 0.608] | 0.3733 [0.331, 0.411] | 0.2189 [0.189, 0.251] | 0.6815 [0.662, 0.699] | 0.4025 [0.254, 0.573] | 0.5532 [0.390, 0.682] | 0.0227 [0.014, 0.058] |
| | Med-BLIP | 0.3017 [0.262, 0.345] | 0.3409 [0.333, 0.500] | 0.3017 [0.262, 0.345] | 0.1543 [0.138, 0.171] | 0.6667 [0.667, 0.667] | 0.0000 [0.000, 0.000] | 0.4531 [0.293, 0.553] | 0.0000 [0.000, 0.000] |
| | Merlin | 0.3017 [0.262, 0.346] | 0.3410 [0.333, 0.500] | 0.3017 [0.262, 0.346] | 0.1544 [0.138, 0.171] | 0.6667 [0.667, 0.667] | 0.0000 [0.000, 0.000] | 0.6318 [0.493, 0.722] | 0.0000 [0.000, 0.000] |
| | PubMedCLIP | 0.3017 [0.262, 0.346] | 0.3410 [0.333, 0.500] | 0.3017 [0.262, 0.346] | 0.1544 [0.138, 0.171] | 0.6667 [0.667, 0.667] | 0.0000 [0.000, 0.000] | 0.5147 [0.501, 0.530] | 0.0000 [0.000, 0.000] |
| | **Ours** | **0.7550** [0.714, 0.792] | **0.4874** [0.446, 0.723] | **0.7121** [0.666, 0.757] | **0.4751** [0.444, 0.505] | **0.8060** [0.775, 0.836] | **0.4784** [0.432, 0.713] | **0.8319** [0.794, 0.867] | **0.4221** [0.331, 0.511] |
| | *p-value* | *0.0005* | *0.4523* | *0.0005* | *0.0005* | *0.0005* | *0.9051* | *0.0005* | *0.0005* |
| Out of Distribution 4 | CT-CLIP | 0.3163 [0.276, 0.358] | 0.2845 [0.244, 0.325] | 0.2905 [0.251, 0.330] | 0.2829 [0.243, 0.322] | 0.6413 [0.620, 0.663] | 0.4111 [0.370, 0.452] | 0.4495 [0.413, 0.488] | -0.0731 [-0.130, -0.011] |
| | Med-BLIP | 0.2002 [0.168, 0.236] | 0.2673 [0.230, 0.306] | 0.2008 [0.164, 0.240] | 0.1694 [0.140, 0.198] | 0.6446 [0.632, 0.657] | 0.2527 [0.222, 0.283] | 0.4233 [0.388, 0.457] | -0.0658 [-0.103, -0.027] |
| | Merlin | 0.5016 [0.460, 0.544] | 0.3333 [0.333, 0.3333] | 0.5016 [0.460, 0.544] | 0.2226 [0.210, 0.234] | 0.6667 [0.667, 0.667] | 0.0000 [0.000, 0.000] | 0.5000 [0.500, 0.500] | 0.0000 [0.000, 0.000] |
| | PubMedCLIP | 0.2505 [0.214, 0.288] | 0.3333 [0.333, 0.333] | 0.2505 [0.214, 0.288] | 0.1334 [0.117, 0.149] | 0.6667 [0.667, 0.667] | 0.0000 [0.000, 0.000] | NaN [nan, nan] | 0.0000 [0.000, 0.000] |
| | **Ours** | **0.5838** [0.544, 0.624] | **0.5980** [0.556, 0.640] | **0.5827** [0.540, 0.625] | **0.5805** [0.539, 0.621] | **0.7837** [0.762, 0.805] | **0.6818** [0.647, 0.716] | **0.7753** [0.745, 0.804] | **0.3561** [0.295, 0.417] |
| | *p-value* | *0.0115* | *0.0005* | *0.0200* | *0.0005* | *0.0005* | *0.0005* | *0.0005* | *0.0005* |
| Out of Distribution 5 | CT-CLIP | 0.4448 [0.394, 0.500] | 0.4452 [0.388, 0.508] | 0.3482 [0.313, 0.384] | 0.3062 [0.271, 0.344] | 0.6863 [0.651, 0.722] | 0.5152 [0.462, 0.571] | NaN [nan, nan] | 0.0328 [-0.046, 0.114] |
| | Med-BLIP | 0.4660 [0.412, 0.518] | 0.4552 [0.401, 0.510] | 0.4616 [0.414, 0.509] | 0.2950 [0.262, 0.329] | 0.6368 [0.600, 0.673] | 0.4532 [0.396, 0.510] | NaN [nan, nan] | -0.077 [-0.171, 0.018] |
| | Merlin | 0.6902 [0.638, 0.738] | 0.5000 [0.500, 0.500] | 0.6902 [0.638, 0.738] | 0.2721 [0.259, 0.283] | 0.6667 [0.667, 0.667] | 0.0000 [0.000, 0.000] | NaN [nan, nan] | 0.0000 [0.000, 0.000] |
| | PubMedCLIP | 0.3097 [0.262, 0.361] | 0.5000 [0.500, 0.500] | 0.3097 [0.262, 0.361] | 0.1574 [0.138, 0.177] | 0.6667 [0.667, 0.667] | 0.0000 [0.000, 0.000] | NaN [nan, nan] | 0.0000 [0.000, 0.000] |
| | **Ours** | **0.8065** [0.762, 0.846] | **0.7472** [0.694, 0.797] | **0.7818** [0.726, 0.834] | **0.5062** [0.470, 0.539] | **0.8314** [0.796, 0.864] | **0.7301** [0.668, 0.786] | NaN [nan, nan] | **0.5211** [0.417, 0.619] |
| | *p-value* | *0.0005* | *0.0005* | *0.0170* | *0.0005* | *0.0005* | *0.0005* | *0.0005* | *0.0005* |

**Supplementary Table 1 | Comparison of emergency triage classification performance across internal and external cohorts.** Classification performance is summarized for CHIEF and four existing medical vision-language models across one internal cohort and five independent external cohorts. The task assigns each examination to negative, non-emergency-positive or positive categories. Performance was assessed using accuracy, balanced accuracy, precision, recall, F1 score, specificity, G-mean, AUROC and Cohen's kappa, with higher values indicating better performance. Within each cohort, the highest value for a given metric is highlighted, the second-highest value is underlined, and values for CHIEF are shown in bold. Values in brackets indicate 95% confidence intervals. P values indicate the statistical significance of the comparison between CHIEF and the best-performing non-CHIEF comparator for the corresponding metric within each cohort. AUROC is reported as undefined when the relevant class distribution does not contain both positive and negative samples.

| Data Source | Method | BLEU↑ | ROUGE-L↑ | METEOR↑ | CIDEr↑ | BERT↑ |
|---|---|---|---|---|---|---|
| | CT-CLIP | 0.1460 [0.137, 0.155] | 0.2384 [0.229, 0.249] | 0.3302 [0.318, 0.342] | 0.3024 [0.259, 0.347] | 0.7331 [0.729, 0.737] |
| | Med-BLIP | 0.1067 [0.101, 0.112] | 0.1951 [0.189, 0.202] | 0.3436 [0.333, 0.354] | 0.0405 [0.031, 0.052] | 0.7210 [0.717, 0.724] |
| | Merlin | 0.0502 [0.048, 0.052] | 0.1184 [0.116, 0.121] | 0.2571 [0.252, 0.262] | 0.0329 [0.026, 0.040] | 0.6844 [0.682, 0.686] |
| | PubMedCLIP | 0.0544 [0.044, 0.065] | 0.1019 [0.091, 0.113] | 0.1341 [0.121, 0.146] | 0.3698 [0.283, 0.461] | 0.6682 [0.662, 0.674] |
| | **Ours** | **0.2305** [0.217, 0.244] | **0.2912** [0.280, 0.303] | **0.3813** [0.367, 0.395] | **0.5032** [0.428, 0.582] | **0.7559** [0.751, 0.761] |
| | *p-value* | *< 0.0001* | *< 0.0001* | *0.0003* | *< 0.0001* | *< 0.0001* |
| Out of Distribution 1 | CT-CLIP | 0.0589 [0.054, 0.064] | 0.1388 [0.131, 0.146] | 0.2586 [0.245, 0.274] | 0.0363 [0.024, 0.051] | 0.6833 [0.679, 0.688] |
| | Med-BLIP | 0.0342 [0.032, 0.037] | 0.0996 [0.094, 0.105] | 0.2441 [0.233, 0.255] | 0.0082 [0.003, 0.014] | 0.6736 [0.670, 0.678] |
| | Merlin | 0.0229 [0.022, 0.024] | 0.0712 [0.068, 0.075] | 0.2073 [0.201, 0.214] | 0.0081 [0.003, 0.015] | 0.6581 [0.656, 0.661] |
| | PubMedCLIP | 0.0427 [0.039, 0.047] | 0.1060 [0.099, 0.113] | 0.1876 [0.176, 0.199] | 0.0108 [0.005, 0.017] | 0.6607 [0.656, 0.666] |
| | **Ours** | **0.1803** [0.152, 0.209] | **0.3052** [0.280, 0.331] | **0.3387** [0.311, 0.364] | **0.8649** [0.722, 1.018] | **0.7614** [0.751, 0.772] |
| | *p-value* | *< 0.0001* | *< 0.0001* | *0.0013* | *< 0.0001* | *< 0.0001* |
| Out of Distribution 2 | CT-CLIP | 0.0386 [0.035, 0.042] | 0.0566 [0.052, 0.062] | 0.1942 [0.181, 0.207] | 0.0315 [0.024, 0.040] | 0.6439 [0.638, 0.650] |
| | Med-BLIP | 0.0233 [0.021, 0.025] | 0.0430 [0.039, 0.047] | 0.2002 [0.191, 0.211] | 0.0040 [0.001, 0.007] | 0.6393 [0.633, 0.645] |
| | Merlin | 0.0241 [0.023, 0.026] | 0.0431 [0.040, 0.047] | 0.1947 [0.188, 0.202] | 0.0044 [0.002, 0.008] | 0.6365 [0.631, 0.642] |
| | PubMedCLIP | 0.0666 [0.063, 0.070] | 0.0874 [0.084, 0.091] | 0.3096 [0.292, 0.328] | 0.0802 [0.063, 0.097] | 0.6746 [0.671, 0.678] |
| | **Ours** | **0.1419** [0.127, 0.158] | **0.1416** [0.128, 0.156] | 0.3047 [0.282, 0.329] | **0.2065** [0.155, 0.265] | **0.6863** [0.678, 0.693] |
| | *p-value* | *< 0.0001* | *< 0.0001* | *0.4081* | *0.0055* | *0.0120* |
| Out of Distribution 3 | CT-CLIP | 0.0650 [0.059, 0.071] | 0.1623 [0.156, 0.169] | 0.1834 [0.175, 0.192] | 0.0544 [0.043, 0.066] | 0.6867 [0.683, 0.690] |
| | Med-BLIP | 0.0346 [0.032, 0.037] | 0.1203 [0.117, 0.123] | 0.1850 [0.180, 0.190] | 0.0199 [0.013, 0.028] | 0.6731 [0.671, 0.675] |
| | Merlin | 0.1011 [0.099, 0.103] | 0.1515 [0.150, 0.153] | 0.2961 [0.293, 0.299] | 0.0281 [0.021, 0.036] | 0.6988 [0.698, 0.700] |
| | PubMedCLIP | 0.0887 [0.087, 0.090] | 0.2353 [0.230, 0.240] | 0.2865 [0.282, 0.291] | 0.0666 [0.054, 0.080] | 0.7407 [0.739, 0.742] |
| | **Ours** | **0.2061** [0.188, 0.222] | **0.2834** [0.266, 0.300] | **0.3062** [0.289, 0.325] | **0.1239** [0.086, 0.171] | **0.7536** [0.746, 0.761] |
| | *p-value* | *< 0.0001* | *< 0.0001* | *0.0232* | *0.0045* | *0.0075* |
| Out of Distribution 4 | CT-CLIP | 0.0335 [0.030, 0.037] | 0.1105 [0.106, 0.115] | 0.1388 [0.131, 0.147] | 0.0436 [0.035, 0.053] | 0.6531 [0.650, 0.657] |
| | Med-BLIP | 0.0415 [0.039, 0.044] | 0.1243 [0.121, 0.128] | 0.2008 [0.194, 0.208] | 0.0306 [0.023, 0.039] | 0.6704 [0.667, 0.674] |
| | Merlin | 0.0260 [0.024, 0.028] | 0.1003 [0.098, 0.103] | 0.1780 [0.172, 0.184] | 0.0213 [0.016, 0.027] | 0.6463 [0.643, 0.650] |
| | PubMedCLIP | 0.0158 [0.014, 0.018] | 0.0556 [0.052, 0.059] | 0.0894 [0.084, 0.095] | 0.0097 [0.007, 0.013] | 0.6494 [0.647, 0.652] |
| | **Ours** | **0.1249** [0.108, 0.143] | **0.2155** [0.203, 0.229] | **0.2400** [0.223, 0.257] | **0.2722** [0.204, 0.350] | **0.6973** [0.691, 0.703] |
| | *p-value* | *< 0.0001* | *< 0.0001* | *0.0159* | *0.8385* | *< 0.0001* |
| Out of Distribution 5 | CT-CLIP | 0.0297 [0.026, 0.033] | 0.0670 [0.061, 0.073] | 0.1338 [0.123, 0.145] | 0.0392 [0.025, 0.056] | 0.6236 [0.619, 0.628] |
| | Med-BLIP | 0.0150 [0.013, 0.017] | 0.0613 [0.057, 0.066] | 0.1320 [0.124, 0.140] | 0.0006 [0.000, 0.002] | 0.6123 [0.609, 0.615] |
| | Merlin | 0.0169 [0.015, 0.018] | 0.0662 [0.063, 0.070] | 0.1459 [0.139, 0.153] | 0.0023 [0.000, 0.006] | 0.6046 [0.601, 0.608] |
| | PubMedCLIP | 0.0334 [0.030, 0.037] | 0.0596 [0.054, 0.065] | 0.1298 [0.119, 0.141] | 0.0537 [0.034, 0.076] | 0.6475 [0.643, 0.652] |
| | **Ours** | **0.1116** [0.098, 0.124] | **0.1486** [0.136, 0.162] | **0.2097** [0.194, 0.226] | **0.1612** [0.117, 0.210] | **0.6626** [0.656, 0.669] |
| | *p-value* | *< 0.0001* | *< 0.0001* | *< 0.0001* | *< 0.0001* | *0.0004* |

**Supplementary Table 2 | Comparison of report generation performance with existing medical vision-language models across internal and external cohorts.** Report generation performance is summarized for the proposed model and four previously reported medical vision-language models across one internal cohort and five independent external cohorts. The first block represents the internal evaluation cohort, and the remaining blocks represent external cohorts from institutions not used for model development. Performance was assessed using BLEU, ROUGE-L, METEOR, CIDEr and BERTScore, with higher values indicating closer agreement with the reference radiology reports. Within each cohort, the highest value for a given metric is highlighted, the second highest value is underlined, and values for the proposed model are shown in bold. Values in brackets indicate confidence intervals. P values indicate the significance of the comparison between the proposed model and competing methods for the corresponding metric within each cohort. Performance varied across centres and comparator models, with the proposed model maintaining competitive performance across both internal and external evaluations.

| Data Source | Method | BLEU↑ | ROUGE-L↑ | METEOR↑ | CIDEr↑ | BERT↑ |
|---|---|---|---|---|---|---|
| | Gemini 3 pro | 0.0172 [0.016, 0.018] | 0.0906 [0.087, 0.094] | 0.1177 [0.113, 0.122] | 0.0000 [0.000, 0.000] | 0.6535 [0.649, 0.658] |
| | Claude 4.5 sonnet | 0.0130 [0.012, 0.013] | 0.0682 [0.066, 0.070] | 0.1056 [0.102, 0.109] | 0.0000 [0.000, 0.000] | 0.6462 [0.643, 0.650] |
| | GPT 5 | 0.0181 [0.017, 0.019] | 0.0964 [0.094, 0.098] | 0.1461 [0.142, 0.150] | 0.0000 [0.000, 0.000] | 0.6729 [0.670, 0.676] |
| | Qwen 3 Instruct | 0.0143 [0.013, 0.015] | 0.0653 [0.063, 0.067] | 0.1259 [0.122, 0.130] | 0.0000 [0.000, 0.000] | 0.6706 [0.667, 0.674] |
| | **Ours** | **0.2305** [0.217, 0.244] | **0.2912** [0.280, 0.303] | **0.3813** [0.367, 0.395] | **0.5032** [0.428, 0.582] | **0.7559** [0.751, 0.761] |
| | *p-value* | *< 0.0001* | *< 0.0001* | *< 0.0001* | *< 0.0001* | *< 0.0001* |
| Out of Distribution 1 | Gemini 3 pro | 0.0045 [0.004, 0.005] | 0.0558 [0.054, 0.058] | 0.0683 [0.065, 0.072] | 0.0000 [0.000, 0.000] | 0.6082 [0.604, 0.612] |
| | Claude 4.5 sonnet | 0.0050 [0.004, 0.006] | 0.0390 [0.037, 0.042] | 0.0569 [0.052, 0.061] | 0.0000 [0.000, 0.000] | 0.5984 [0.593, 0.604] |
| | GPT 5 | 0.0109 [0.017, 0.019] | 0.0649 [0.094, 0.099] | 0.1094 [0.142, 0.150] | 0.0000 [0.000, 0.000] | 0.6297 [0.670, 0.676] |
| | Qwen 3 Instruct | 0.0080 [0.007, 0.009] | 0.0431 [0.041, 0.045] | 0.0769 [0.073, 0.081] | 0.0000 [0.000, 0.000] | 0.6223 [0.618, 0.626] |
| | **Ours** | **0.1803** [0.152, 0.209] | **0.3052** [0.280, 0.331] | **0.3387** [0.311, 0.364] | **0.8649** [0.722, 1.018] | **0.7614** [0.751, 0.772] |
| | *p-value* | *< 0.0001* | *< 0.0001* | *< 0.0001* | *< 0.0001* | *< 0.0001* |
| Out of Distribution 2 | Gemini 3 pro | 0.0067 [0.006, 0.008] | 0.0503 [0.048, 0.053] | 0.0511 [0.048, 0.055] | 0.0000 [0.000, 0.000] | 0.5862 [0.581, 0.592] |
| | Claude 4.5 sonnet | 0.0085 [0.007, 0.010] | 0.0415 [0.037, 0.047] | 0.0559 [0.047, 0.065] | 0.0000 [0.000, 0.000] | 0.5910 [0.580, 0.602] |
| | GPT 5 | 0.0116 [0.011, 0.013] | 0.0616 [0.058, 0.065] | 0.0714 [0.067, 0.077] | 0.0000 [0.000, 0.000] | 0.6151 [0.610, 0.621] |
| | Qwen 3 Instruct | 0.0091 [0.008, 0.009] | 0.0394 [0.037, 0.042] | 0.0641 [0.060, 0.069] | 0.0000 [0.000, 0.000] | 0.6041 [0.598, 0.611] |
| | **Ours** | **0.1419** [0.127, 0.158] | **0.1416** [0.128, 0.156] | **0.3047** [0.282, 0.329] | **0.2065** [0.155, 0.265] | **0.6863** [0.678, 0.693] |
| | *p-value* | *< 0.0001* | *< 0.0001* | *< 0.0001* | *< 0.0001* | *< 0.0001* |
| Out of Distribution 3 | Gemini 3 pro | 0.0546 [0.052, 0.057] | 0.1714 [0.165, 0.177] | 0.2432 [0.234, 0.252] | 0.0000 [0.000, 0.000] | 0.7132 [0.707, 0.720] |
| | Claude 4.5 sonnet | 0.0386 [0.038, 0.040] | 0.1219 [0.119, 0.124] | 0.2007 [0.196, 0.205] | 0.0000 [0.000, 0.000] | 0.7183 [0.716, 0.720] |
| | GPT 5 | 0.0652 [0.064, 0.067] | 0.1803 [0.177, 0.184] | 0.2911 [0.285, 0.297] | 0.0000 [0.000, 0.000] | 0.7579 [0.756, 0.760] |
| | Qwen 3 Instruct | 0.0435 [0.043, 0.045] | 0.1105 [0.109, 0.113] | 0.2163 [0.213, 0.220] | 0.0000 [0.000, 0.000] | 0.7332 [0.732, 0.735] |
| | **Ours** | **0.2061** [0.188, 0.222] | **0.2834** [0.266, 0.300] | **0.3062** [0.289, 0.325] | **0.1239** [0.086, 0.171] | **0.7536** [0.746, 0.761] |
| | *p-value* | *< 0.0001* | *< 0.0001* | *< 0.0001* | *< 0.0001* | *< 0.0001* |
| Out of Distribution 4 | Gemini 3 pro | 0.0226 [0.021, 0.024] | 0.0858 [0.083, 0.088] | 0.2258 [0.221, 0.230] | 0.0016 [0.001, 0.003] | 0.6298 [0.627, 0.632] |
| | Claude 4.5 sonnet | 0.0170 [0.016, 0.018] | 0.0659 [0.063, 0.124] | 0.2232 [0.217, 0.228] | 0.0000 [0.000, 0.000] | 0.6155 [0.612, 0.618] |
| | GPT 5 | 0.0203 [0.018, 0.021] | 0.0858 [0.083, 0.088] | 0.2377 [0.232, 0.242] | 0.0001 [0.000, 0.000] | 0.6298 [0.627, 0.632] |
| | Qwen 3 Instruct | 0.0173 [0.016, 0.018] | 0.0702 [0.067, 0.072] | 0.2452 [0.239, 0.250] | 0.0000 [0.000, 0.000] | 0.6277 [0.624, 0.631] |
| | **Ours** | **0.1249** [0.108, 0.143] | **0.2155** [0.203, 0.229] | **0.2400** [0.223, 0.257] | **0.2722** [0.204, 0.350] | **0.6973** [0.691, 0.703] |
| | *p-value* | *< 0.0001* | *< 0.0001* | *< 0.0001* | *< 0.0001* | *< 0.0001* |
| Out of Distribution 5 | Gemini 3 pro | 0.0082 [0.007, 0.009] | 0.0388 [0.036, 0.040] | 0.1468 [0.140, 0.154] | 0.0000 [0.000, 0.000] | 0.5717 [0.564, 0.577] |
| | Claude 4.5 sonnet | 0.0064 [0.005, 0.009] | 0.0320 [0.030, 0.033] | 0.1293 [0.123, 0.135] | 0.0000 [0.000, 0.000] | 0.5591 [0.556, 0.562] |
| | GPT 5 | 0.0068 [0.006, 0.007] | 0.0406 [0.039, 0.042] | 0.1400 [0.134, 0.146] | 0.0000 [0.000, 0.000] | 0.5733 [0.570, 0.576] |
| | Qwen 3 Instruct | 0.0058 [0.005, 0.006] | 0.0280 [0.026, 0.029] | 0.1281 [0.123, 0.133] | 0.0000 [0.000, 0.000] | 0.5581 [0.555, 0.560] |
| | **Ours** | **0.1116** [0.098, 0.124] | **0.1486** [0.136, 0.162] | **0.2097** [0.194, 0.226] | **0.1612** [0.117, 0.210] | **0.6626** [0.656, 0.669] |
| | *p-value* | *< 0.0001* | *< 0.0001* | *< 0.0001* | *< 0.0001* | *< 0.0001* |

**Supplementary Table 3 | Comparison of report generation performance between the proposed model and commercial large language models across internal and external cohorts.** Report generation performance is summarized for the proposed model and four commercial large language models across one internal cohort and five independent external cohorts. The first block corresponds to the internal cohort used for in distribution evaluation, whereas the remaining blocks correspond to five external cohorts used for out of centre evaluation. Performance was assessed using BLEU, ROUGE-L, METEOR, CIDEr and BERT Score. Higher values indicate better agreement with the reference radiology reports. For each cohort, the best performing model for a given metric is highlighted, the second-best result is underlined, and results obtained by the proposed model are shown in bold. Bracketed values indicate confidence intervals. P values denote the statistical significance of comparisons between the proposed model and the competing methods for the corresponding metric within each cohort. Across the internal cohort and most external cohorts, the proposed model showed consistently stronger performance than the commercial baselines, supporting more reliable report generation under variation in patient population, acquisition protocol and institution.

| Data Source | Method | MRR | Recall@1 | Recall@5 | Recall@10 | Recall@50 | Recall@100 | Recall@200 |
|---|---|---|---|---|---|---|---|---|
| | CT-CLIP | 0.1421 [0.122, 0.162] | 0.1267 [0.106, 0.147] | 0.1493 [0.128, 0.172] | 0.1627 [0.141, 0.185] | 0.2101 [0.186, 0.235] | 0.2379 [0.211, 0.264] | 0.2657 [0.239, 0.292] |
| | Med-BLIP | 0.0165 [0.013, 0.022] | 0.0041 [0.001, 0.008] | 0.0124 [0.006, 0.020] | 0.0216 [0.013, 0.031] | 0.1030 [0.084, 0.123] | 0.1782 [0.156, 0.201] | 0.2894 [0.262, 0.317] |
| | Merlin | 0.0099 [0.009, 0.012] | 0.0000 [0.000, 0.000] | 0.0041 [0.004, 0.004] | 0.0103 [0.010, 0.010] | 0.0772 [0.061, 0.094] | 0.1411 [0.119, 0.164] | 0.2544 [0.225, 0.281] |
| | PubMedCLIP | 0.0103 [0.008, 0.013] | 0.0010 [0.001, 0.001] | 0.0051 [0.005, 0.005] | 0.0103 [0.010, 0.010] | 0.0618 [0.048, 0.077] | 0.1246 [0.104, 0.145] | 0.2214 [0.196, 0.247] |
| | **Ours** | **0.3383 [0.312, 0.368]** | **0.3223 [0.296, 0.352]** | **0.3512 [0.323, 0.381]** | **0.3553 [0.327, 0.385]** | **0.3780 [0.349, 0.409]** | **0.3893 [0.359, 0.420]** | **0.4192 [0.389, 0.449]** |
| | *p-value* | 0.0005 | 0.0005 | 0.0005 | 0.0005 | 0.0005 | 0.0005 | 0.0025 |
| Out of Distribution 1 | CT-CLIP | 0.3927 [0.354, 0.433] | 0.3764 [0.338, 0.417] | 0.3958 [0.355, 0.436] | 0.4170 [0.375, 0.459] | 0.4942 [0.452, 0.535] | 0.5328 [0.490, 0.573] | 0.5772 [0.537, 0.618] |
| | Med-BLIP | 0.0370 [0.027, 0.050] | 0.0154 [0.006, 0.027] | 0.0386 [0.023, 0.056] | 0.0598 [0.041, 0.083] | 0.2838 [0.245, 0.324] | 0.3340 [0.295, 0.375] | 0.4402 [0.398, 0.483] |
| | Merlin | 0.0647 [0.051, 0.080] | 0.0212 [0.010, 0.035] | 0.1564 [0.127, 0.189] | 0.1776 [0.147, 0.210] | 0.2722 [0.237, 0.311] | 0.3784 [0.340, 0.421] | 0.5888 [0.546, 0.633] |
| | PubMedCLIP | 0.2355 [0.205, 0.265] | 0.1602 [0.129, 0.191] | 0.3764 [0.336, 0.417] | 0.4730 [0.430, 0.513] | 0.5135 [0.469, 0.552] | 0.5792 [0.537, 0.618] | 0.6969 [0.658, 0.732] |
| | **Ours** | **0.4241 [0.382, 0.464]** | **0.4112 [0.369, 0.452]** | **0.4266 [0.382, 0.467]** | 0.4363 [0.392, 0.477] | 0.5097 [0.465, 0.548] | 0.5405 [0.494, 0.583] | 0.5772 [0.535, 0.616] |
| | *p-value* | 0.0010 | 0.0005 | 0.0010 | 0.0007 | 0.2636 | 0.0028 | < 0.0001 |
| Out of Distribution 2 | CT-CLIP | 0.4751 [0.433, 0.518] | 0.4556 [0.411, 0.498] | 0.4819 [0.438, 0.526] | 0.5040 [0.462, 0.548] | 0.5988 [0.558, 0.645] | 0.6391 [0.599, 0.683] | 0.7036 [0.667, 0.746] |
| | Med-BLIP | 0.0247 [0.017, 0.034] | 0.0101 [0.002, 0.020] | 0.0161 [0.006, 0.028] | 0.0363 [0.022, 0.052] | 0.1391 [0.111, 0.171] | 0.2702 [0.234, 0.309] | 0.4859 [0.442, 0.532] |
| | Merlin | 0.0143 [0.013, 0.016] | 0.0000 [0.000, 0.000] | 0.0020 [0.000, 0.006] | 0.0181 [0.008, 0.032] | 0.1653 [0.133, 0.198] | 0.2964 [0.256, 0.335] | 0.4879 [0.444, 0.532] |
| | PubMedCLIP | 0.1684 [0.157, 0.180] | 0.0020 [0.000, 0.006] | 0.6270 [0.585, 0.673] | 0.6331 [0.591, 0.677] | 0.6915 [0.649, 0.734] | 0.8004 [0.764, 0.837] | 0.8730 [0.845, 0.903] |
| | **Ours** | **0.5727 [0.532, 0.617]** | **0.5565 [0.514, 0.601]** | 0.5847 [0.542, 0.631] | 0.5867 [0.544, 0.633] | 0.6371 [0.597, 0.679] | 0.6714 [0.633, 0.714] | 0.7198 [0.683, 0.760] |
| | *p-value* | 0.0005 | 0.0005 | < 0.0001 | < 0.0001 | < 0.0001 | < 0.0001 | < 0.0001 |
| Out of Distribution 3 | CT-CLIP | 0.5068 [0.464, 0.548] | 0.4906 [0.447, 0.532] | 0.5220 [0.478, 0.564] | 0.5346 [0.491, 0.577] | 0.5744 [0.532, 0.616] | 0.6562 [0.616, 0.700] | 0.6667 [0.625, 0.711] |
| | Med-BLIP | 0.0363 [0.030, 0.044] | 0.0042 [0.000, 0.010] | 0.0314 [0.017, 0.048] | 0.0692 [0.046, 0.092] | 0.3627 [0.319, 0.405] | 0.5891 [0.543, 0.631] | 0.6038 [0.560, 0.648] |
| | Merlin | 0.0330 [0.027, 0.040] | 0.0042 [0.000, 0.011] | 0.0147 [0.006, 0.027] | 0.0503 [0.031, 0.071] | 0.5136 [0.468, 0.560] | 0.6457 [0.600, 0.690] | 0.6457 [0.600, 0.690] |
| | PubMedCLIP | 0.1333 [0.122, 0.144] | 0.0021 [0.000, 0.006] | 0.4696 [0.428, 0.514] | 0.4885 [0.444, 0.532] | 0.6310 [0.591, 0.673] | 0.6876 [0.646, 0.725] | 0.8113 [0.776, 0.845] |
| | **Ours** | **0.6244 [0.584, 0.666]** | **0.6080 [0.564, 0.650]** | **0.6373 [0.597, 0.679]** | **0.6562 [0.614, 0.696]** | **0.6876 [0.650, 0.727]** | **0.7254 [0.686, 0.765]** | 0.7296 [0.688, 0.769] |
| | *p-value* | 0.0005 | 0.0005 | 0.0005 | 0.0005 | 0.0133 | < 0.0001 | < 0.0001 |
| Out of Distribution 4 | CT-CLIP | 0.1533 [0.123, 0.185] | 0.1420 [0.112, 0.174] | 0.1560 [0.124, 0.190] | 0.1560 [0.124, 0.190] | 0.1580 [0.126, 0.190] | 0.1620 [0.130, 0.194] | 0.1660 [0.134, 0.198] |
| | Med-BLIP | 0.0161 [0.013, 0.020] | 0.0000 [0.000, 0.000] | 0.0080 [0.002, 0.016] | 0.0340 [0.020, 0.050] | 0.1340 [0.106, 0.166] | 0.2460 [0.208, 0.284] | 0.4460 [0.402, 0.490] |
| | Merlin | 0.0150 [0.011, 0.019] | 0.0000 [0.000, 0.000] | 0.0100 [0.002, 0.020] | 0.0240 [0.010, 0.038] | 0.1200 [0.092, 0.150] | 0.2220 [0.184, 0.262] | 0.4260 [0.384, 0.472] |
| | PubMedCLIP | 0.0150 [0.011, 0.020] | 0.0020 [0.000, 0.006] | 0.0100 [0.002, 0.020] | 0.0200 [0.008, 0.034] | 0.1000 [0.074, 0.126] | 0.2000 [0.164, 0.236] | 0.4680 [0.424, 0.512] |
| | **Ours** | **0.2313 [0.193, 0.269]** | **0.2220 [0.184, 0.260]** | **0.2340 [0.194, 0.272]** | **0.2340 [0.194, 0.272]** | **0.2360 [0.196, 0.274]** | 0.2400 [0.200, 0.278] | **0.2420 [0.202, 0.282]** |
| | *p-value* | 0.0005 | 0.0005 | 0.0005 | 0.0005 | 0.0005 | 0.7896 | < 0.0001 |
| Out of Distribution 5 | CT-CLIP | 0.6944 [0.645, 0.747] | 0.6898 [0.639, 0.744] | 0.6958 [0.648, 0.747] | 0.6958 [0.648, 0.747] | 0.6988 [0.651, 0.750] | 0.7018 [0.654, 0.753] | 0.7048 [0.657, 0.756] |
| | Med-BLIP | 0.0301 [0.019, 0.043] | 0.0090 [0.000, 0.021] | 0.0331 [0.015, 0.054] | 0.0422 [0.021, 0.066] | 0.1687 [0.130, 0.211] | 0.3434 [0.292, 0.395] | 0.6416 [0.593, 0.699] |
| | Merlin | 0.0244 [0.018, 0.032] | 0.0030 [0.000, 0.009] | 0.0241 [0.009, 0.039] | 0.0512 [0.027, 0.075] | 0.2259 [0.181, 0.268] | 0.4277 [0.377, 0.482] | 0.7018 [0.648, 0.750] |
| | PubMedCLIP | 0.1173 [0.091, 0.145] | 0.0693 [0.042, 0.096] | 0.1205 [0.090, 0.157] | 0.1867 [0.145, 0.229] | 0.4066 [0.358, 0.461] | 0.6325 [0.581, 0.678] | **0.8283 [0.789, 0.868]** |
| | **Ours** | **0.7184 [0.672, 0.769]** | **0.7108 [0.663, 0.762]** | **0.7259 [0.678, 0.777]** | **0.7289 [0.684, 0.780]** | **0.7289 [0.684, 0.780]** | **0.7289 [0.684, 0.780]** | 0.7319 [0.684, 0.783] |
| | *p-value* | 0.0045 | 0.0085 | 0.0020 | 0.0020 | 0.0020 | 0.0050 | < 0.0001 |

**Supplementary Table 4 | Comparison of report retrieval performance measured by mean reciprocal rank and recall across internal and external cohorts.** Report retrieval performance is summarized for the proposed model and four existing medical vision-language models across one internal cohort and five independent external cohorts. The first block represents the internal evaluation cohort, and the remaining blocks represent external cohorts from institutions not used for model development. Performance was assessed using mean reciprocal rank and recall at 1, 5, 10, 50, 100 and 200, with higher values indicating better retrieval performance. Within each cohort, the highest value for a given metric is highlighted, the second highest value is underlined, and values for the proposed model are shown in bold. Values in brackets indicate confidence intervals. P values indicate the significance of the comparison between the proposed model and competing methods for the corresponding metric within each cohort. The results show variation in retrieval performance across centers, while the proposed model maintained strong performance in both internal and external evaluations.

| Data Source | Method | NDCG@1 | NDCG@5 | NDCG@10 | NDCG@50 | NDCG@100 | NDCG@200 |
|---|---|---|---|---|---|---|---|
| | CT-CLIP | 0.1267 [0.106, 0.147] | 0.1378 [0.117, 0.158] | 0.1422 [0.121, 0.163] | 0.1523 [0.132, 0.172] | 0.1568 [0.137, 0.177] | 0.1606 [0.141, 0.181] |
| | Med-BLIP | 0.0041 [0.001, 0.008] | 0.0078 [0.004, 0.013] | 0.0108 [0.006, 0.016] | 0.0284 [0.023, 0.035] | 0.0405 [0.034, 0.047] | 0.0560 [0.050, 0.063] |
| | Merlin | 0.0000 [0.000, 0.000] | 0.0020 [0.000, 0.004] | 0.0040 [0.002, 0.007] | 0.0177 [0.014, 0.022] | 0.0280 [0.024, 0.032] | 0.0437 [0.038, 0.049] |
| | PubMedCLIP | 0.0010 [0.000, 0.003] | 0.0030 [0.001, 0.006] | 0.0047 [0.002, 0.008] | 0.0152 [0.011, 0.019] | 0.0253 [0.021, 0.030] | 0.0388 [0.034, 0.044] |
| | **Ours** | **0.3223** [0.296, 0.352] | **0.3378** [0.311, 0.367] | **0.3392** [0.312, 0.369] | **0.3443** [0.317, 0.374] | **0.3462** [0.319, 0.376] | **0.3504** [0.323, 0.380] |
| | *p-value* | *0.0005* | *0.0005* | *0.0005* | *0.0005* | *0.0005* | *0.0005* |
| Out of Distribution 1 | CT-CLIP | 0.3764 [0.338, 0.417] | 0.3864 [0.347, 0.427] | 0.3933 [0.353, 0.434] | 0.4102 [0.372, 0.449] | 0.4164 [0.379, 0.454] | 0.4226 [0.386, 0.460] |
| | Med-BLIP | 0.0154 [0.006, 0.027] | 0.0267 [0.016, 0.041] | 0.0337 [0.022, 0.048] | 0.0779 [0.065, 0.092] | 0.0860 [0.073, 0.101] | 0.1005 [0.088, 0.115] |
| | Merlin | 0.0212 [0.010, 0.035] | 0.0770 [0.060, 0.095] | 0.0839 [0.066, 0.102] | 0.1047 [0.089, 0.123] | 0.1218 [0.106, 0.141] | 0.1509 [0.135, 0.168] |
| | PubMedCLIP | 0.1602 [0.129, 0.191] | 0.2565 [0.226, 0.288] | 0.2865 [0.256, 0.316] | 0.2951 [0.265, 0.325] | 0.3056 [0.276, 0.335] | 0.3220 [0.293, 0.350] |
| | **Ours** | **0.4112** [0.369, 0.452] | **0.4197** [0.377, 0.459] | **0.4228** [0.380, 0.462] | **0.4377** [0.396, 0.478] | **0.4428** [0.402, 0.483] | **0.4480** [0.407, 0.488] |
| | *p-value* | *0.0005* | *0.0005* | *0.0020* | *0.0010* | *0.0015* | *0.0010* |
| Out of Distribution 2 | CT-CLIP | 0.4556 [0.411, 0.498] | 0.4694 [0.425, 0.513] | 0.4768 [0.435, 0.519] | 0.4973 [0.457, 0.540] | 0.5037 [0.464, 0.546] | 0.5128 [0.474, 0.554] |
| | Med-BLIP | 0.0101 [0.002, 0.020] | 0.0131 [0.005, 0.024] | 0.0195 [0.011, 0.030] | 0.0411 [0.031, 0.053] | 0.0622 [0.052, 0.074] | 0.0924 [0.082, 0.104] |
| | Merlin | 0.0000 [0.000, 0.000] | 0.0009 [0.000, 0.003] | 0.0058 [0.002, 0.010] | 0.0372 [0.030, 0.044] | 0.0583 [0.050, 0.066] | 0.0851 [0.077, 0.094] |
| | PubMedCLIP | 0.0020 [0.000, 0.006] | 0.2747 [0.257, 0.295] | 0.2766 [0.258, 0.296] | 0.2887 [0.271, 0.307] | 0.3062 [0.291, 0.323] | 0.3162 [0.301, 0.332] |
| | **Ours** | **0.5565** [0.514, 0.601] | **0.5720** [0.531, 0.616] | **0.5726** [0.532, 0.617] | **0.5838** [0.543, 0.626] | **0.5894** [0.550, 0.632] | **0.5961** [0.559, 0.638] |
| | *p-value* | *0.0005* | *0.0005* | *0.0005* | *0.0005* | *0.0005* | *0.0005* |
| Out of Distribution 3 | CT-CLIP | 0.4906 [0.447, 0.532] | 0.5056 [0.463, 0.547] | 0.5094 [0.467, 0.551] | 0.5185 [0.476, 0.559] | 0.5319 [0.490, 0.570] | 0.5333 [0.492, 0.572] |
| | Med-BLIP | 0.0042 [0.000, 0.010] | 0.0162 [0.008, 0.026] | 0.0281 [0.019, 0.040] | 0.0936 [0.081, 0.107] | 0.1316 [0.119, 0.144] | 0.1337 [0.122, 0.146] |
| | Merlin | 0.0042 [0.000, 0.011] | 0.0093 [0.003, 0.017] | 0.0201 [0.012, 0.029] | 0.1161 [0.105, 0.128] | 0.1382 [0.127, 0.149] | 0.1382 [0.127, 0.149] |
| | PubMedCLIP | 0.0021 [0.000, 0.006] | 0.2053 [0.187, 0.225] | 0.2116 [0.192, 0.230] | 0.2429 [0.225, 0.260] | 0.2522 [0.235, 0.269] | 0.2698 [0.253, 0.285] |
| | **Ours** | **0.6080** [0.564, 0.650] | **0.6232** [0.582, 0.665] | **0.6294** [0.589, 0.671] | **0.6361** [0.596, 0.676] | **0.6423** [0.603, 0.682] | **0.6430** [0.604, 0.683] |
| | *p-value* | *0.0005* | *0.0005* | *0.0005* | *0.0005* | *0.0005* | *0.0005* |
| Out of Distribution 4 | CT-CLIP | 0.1420 [0.112, 0.174] | 0.1508 [0.120, 0.183] | 0.1508 [0.120, 0.183] | 0.1512 [0.121, 0.184] | 0.1519 [0.122, 0.184] | 0.1524 [0.122, 0.184] |
| | Med-BLIP | 0.0000 [0.000, 0.000] | 0.0042 [0.001, 0.009] | 0.0121 [0.007, 0.018] | 0.0337 [0.026, 0.042] | 0.0519 [0.043, 0.061] | 0.0796 [0.071, 0.088] |
| | Merlin | 0.0000 [0.000, 0.000] | 0.0054 [0.001, 0.011] | 0.0101 [0.004, 0.016] | 0.0296 [0.022, 0.038] | 0.0460 [0.037, 0.055] | 0.0742 [0.065, 0.083] |
| | PubMedCLIP | 0.0020 [0.000, 0.006] | 0.0059 [0.001, 0.012] | 0.0091 [0.004, 0.016] | 0.0258 [0.018, 0.034] | 0.0419 [0.033, 0.051] | 0.0796 [0.071, 0.089] |
| | **Ours** | **0.2220** [0.184, 0.260] | **0.2290** [0.190, 0.267] | **0.2290** [0.190, 0.267] | **0.2296** [0.191, 0.267] | **0.2302** [0.192, 0.268] | **0.2305** [0.192, 0.269] |
| | *p-value* | *0.0005* | *0.0005* | *0.0005* | *0.0005* | *0.0005* | *0.0005* |
| Out of Distribution 5 | CT-CLIP | 0.6898 [0.639, 0.744] | 0.6936 [0.644, 0.746] | 0.6936 [0.644, 0.746] | 0.6942 [0.645, 0.746] | 0.6947 [0.645, 0.747] | 0.6952 [0.645, 0.747] |
| | Med-BLIP | 0.0090 [0.000, 0.021] | 0.0215 [0.010, 0.036] | 0.0245 [0.012, 0.040] | 0.0507 [0.037, 0.067] | 0.0788 [0.064, 0.095] | 0.1203 [0.106, 0.137] |
| | Merlin | 0.0030 [0.000, 0.009] | 0.0125 [0.004, 0.022] | 0.0209 [0.011, 0.032] | 0.0562 [0.044, 0.069] | 0.0890 [0.077, 0.103] | 0.1272 [0.115, 0.139] |
| | PubMedCLIP | 0.0693 [0.042, 0.096] | 0.1010 [0.073, 0.132] | 0.1219 [0.094, 0.154] | 0.1688 [0.141, 0.200] | 0.2056 [0.180, 0.234] | 0.2330 [0.209, 0.261] |
| | **Ours** | **0.7108** [0.663, 0.762] | **0.7190** [0.671, 0.768] | **0.7200** [0.673, 0.769] | **0.7200** [0.673, 0.769] | **0.7200** [0.673, 0.769] | **0.7204** [0.673, 0.770] |
| | *p-value* | *0.0085* | *0.0030* | *0.0020* | *0.0025* | *0.0025* | *0.0025* |

**Supplementary Table 5 | Comparison of report retrieval performance measured by normalized discounted cumulative gain across internal and external cohorts.** Report retrieval performance is summarized for the proposed model and four existing medical vision-language models across one internal cohort and five independent external cohorts. The first block represents the internal evaluation cohort, and the remaining blocks represent external cohorts from institutions not used for model development. Performance was assessed using normalized discounted cumulative gain at 1, 5, 10, 50, 100 and 200, with higher values indicating better ranking quality of retrieved reports. In the present single-relevant-item setting with binary relevance, NDCG@1 is numerically equivalent to Recall@1 and is retained here only for completeness across NDCG cut-offs. Within each cohort, the highest value for a given metric is highlighted, the second highest value is underlined, and values for the proposed model are shown in bold. Values in brackets indicate confidence intervals. P values indicate the significance of the comparison between the proposed model and competing methods for the corresponding metric within each cohort. The results show variation in retrieval ranking quality across centres, while the proposed model remained stable in both internal and external evaluations.

| Evaluation set | Examinations | Labels | Evaluation level | AUROC | AP |
|---|---|---|---|---|---|
| CQ500 | 245 | 14 | Multilabel label-level detection | Macro: 0.8033 [0.7689–0.8425]; Micro: 0.8466 [0.8194–0.8750] | Macro: 0.3924 [0.3405–0.4789]; Micro: 0.4743 [0.3991–0.5536] |
| Matched cohort | 971 | 45 | Overall abnormality versus negative | 0.6255 [0.5880–0.6625] | 0.7465 [0.7092–0.7860] |
| | | | Fine-grained multilabel detection | Macro: 0.9472 [0.9362–0.9540]; Micro: 0.9698 [0.9667–0.9730] | Macro: 0.3973 [0.3296–0.4697]; Micro: 0.1600 [0.1411–0.1827] |

**Supplementary Table 6 | Summary of internal zero-shot abnormality detection and fine-tuned CQ500 multilabel classification.** The table summarises abnormality-detection performance for two evaluation settings. The internal matched cohort comprised 971 examinations and 45 report-derived abnormality labels and was evaluated in a zero-shot setting at both the overall abnormality-versus-negative level and the fine-grained multilabel level. CQ500 was evaluated as a fine-tuned downstream multilabel classification benchmark, with 1,019 training examinations and 245 validation examinations. CQ500 results are reported on the validation set across 14 haemorrhage- and fracture-related labels. AUROC and average precision (AP) are reported with 95% confidence intervals in brackets. Macro-averaged metrics summarise label-wise performance with equal weight across labels, whereas micro-averaged metrics were computed after pooling predictions across sample-label pairs.

| CQ500 label | Positive cases | AUROC | AP |
|---|---|---|---|
| ICH | 94/245 [38.4%] | 0.7502 [0.686-0.807] | 0.6726 [0.5725-0.7683] |
| IPH | 56/245 [22.9%] | 0.8063 [0.737-0.867] | 0.5275 [0.4112-0.6749] |
| IVH | 9/245 [3.7%] | 0.9557 [0.907-0.995] | 0.5269 [0.2415-0.9002] |
| SDH | 21/245 [8.6%] | 0.5851 [0.484-0.694] | 0.1036 [0.0649-0.1771] |
| EDH | 7/245 [2.9%] | 0.8914 [0.798-0.967] | 0.1792 [0.0643-0.4560] |
| SAH | 32/245 [13.1%] | 0.7566 [0.657-0.846] | 0.4333 [0.2644-0.6053] |
| Bleed Location-Left | 55/245 [22.4%] | 0.7886 [0.713-0.849] | 0.4980 [0.3770-0.6521] |
| Bleed Location-Right | 85/245 [34.7%] | 0.7619 [0.699-0.823] | 0.6421 [0.5437-0.7362] |
| Chronic Bleed | 6/245 [2.4%] | 0.8103 [0.611-0.951] | 0.1030 [0.0312-0.2624] |
| Fracture | 17/245 [6.9%] | 0.8976 [0.798-0.972] | 0.4815 [0.2820-0.7377] |
| Calvarial Fracture | 12/245 [4.9%] | 0.9396 [0.896-0.975] | 0.4818 [0.2450-0.7461] |
| Other Fracture | 5/245 [2.0%] | 0.8250 [0.653-0.969] | 0.1140 [0.0238-0.3363] |
| Mass Effect | 43/245 [17.6%] | 0.6595 [0.557-0.747] | 0.2973 [0.2028-0.4201] |
| Midline Shift | 27/245 [11.0%] | 0.8186 [0.720-0.898] | 0.4328 [0.2657-0.6362] |

**Supplementary Table 7 | Label-wise performance of fine-tuned CQ500 multilabel classification.** Label-wise validation performance of CHIEF is shown for 14 haemorrhage- and fracture-related CQ500 labels. Positive cases are reported as the number and percentage of positive examinations among the 245 validation examinations. AUROC and average precision (AP) are reported with 95% confidence intervals in brackets. ICH, intracranial haemorrhage; IPH, intraparenchymal haemorrhage; IVH, intraventricular haemorrhage; SDH, subdural haemorrhage; EDH, extradural/epidural haemorrhage; SAH, subarachnoid haemorrhage.

| Probe | Input and interface | Output | Evaluation |
|---|---|---|---|
| Report generation | 3D head CT volume encoded as the visual base latent and projected to continuous prefix embeddings for the autoregressive language model. | Generated Chinese radiology report. | BLEU, ROUGE-L, METEOR, CIDEr, BERTScore, expert review and blinded Turing study. |
| Emergency triage classification | Visual base latent with a lightweight classification head; generation-derived features could be added as a downstream enhancement. | Negative, non-emergency-positive or positive class. | ACC, BACC, precision, recall, F1, specificity, G-mean, AUROC and Cohen kappa. |
| Cross-modal retrieval | 3D head CT volume encoded as the image query; candidate radiology reports encoded as text latents and ranked by cosine similarity. | Ranked radiology reports. | Recall@K, MRR and NDCG@K for image-to-text retrieval, with representative top-ranked retrieval examples. |
| CQ500 multilabel classification | CQ500 head CT volume encoded by the visual branch and evaluated with a multilabel binary head without label text input. | Label probabilities for haemorrhage- and fracture-related abnormalities. | Macro/micro AUROC and AP with bootstrap confidence intervals and label-wise positive counts. |
| Zero-shot abnormality detection | 3D head CT volume with predefined abnormality-question templates; yes/no probabilities estimated using the image-conditioned generation branch with template ensembling and calibration. | Abnormality probability for each label. | Overall and label-wise AUROC/AP, thresholded metrics, prevalence and score-distribution summaries. |

**Supplementary Table 8 | Implementation summary of downstream probes.** The table summarises how each downstream task was connected to the pretrained CHIEF representation, including the input interface, output space and evaluation endpoints. Report generation, triage classification, cross-modal retrieval, CQ500 multilabel classification and zero-shot abnormality detection were implemented as probes of the same pretrained representation rather than as independent task-specific systems.

#### Supplementary Figures

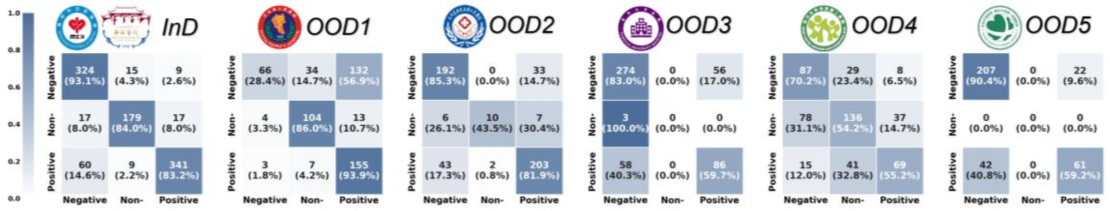

**Supplementary Fig. 1 | Class-wise performance of emergency triage classification across internal and external cohorts.** Confusion matrices and class-wise recall are shown for negative, non-emergency-positive and positive examinations across the internal cohort and five external cohorts. These panels illustrate how classification performance varies across centres and across clinically distinct triage categories.

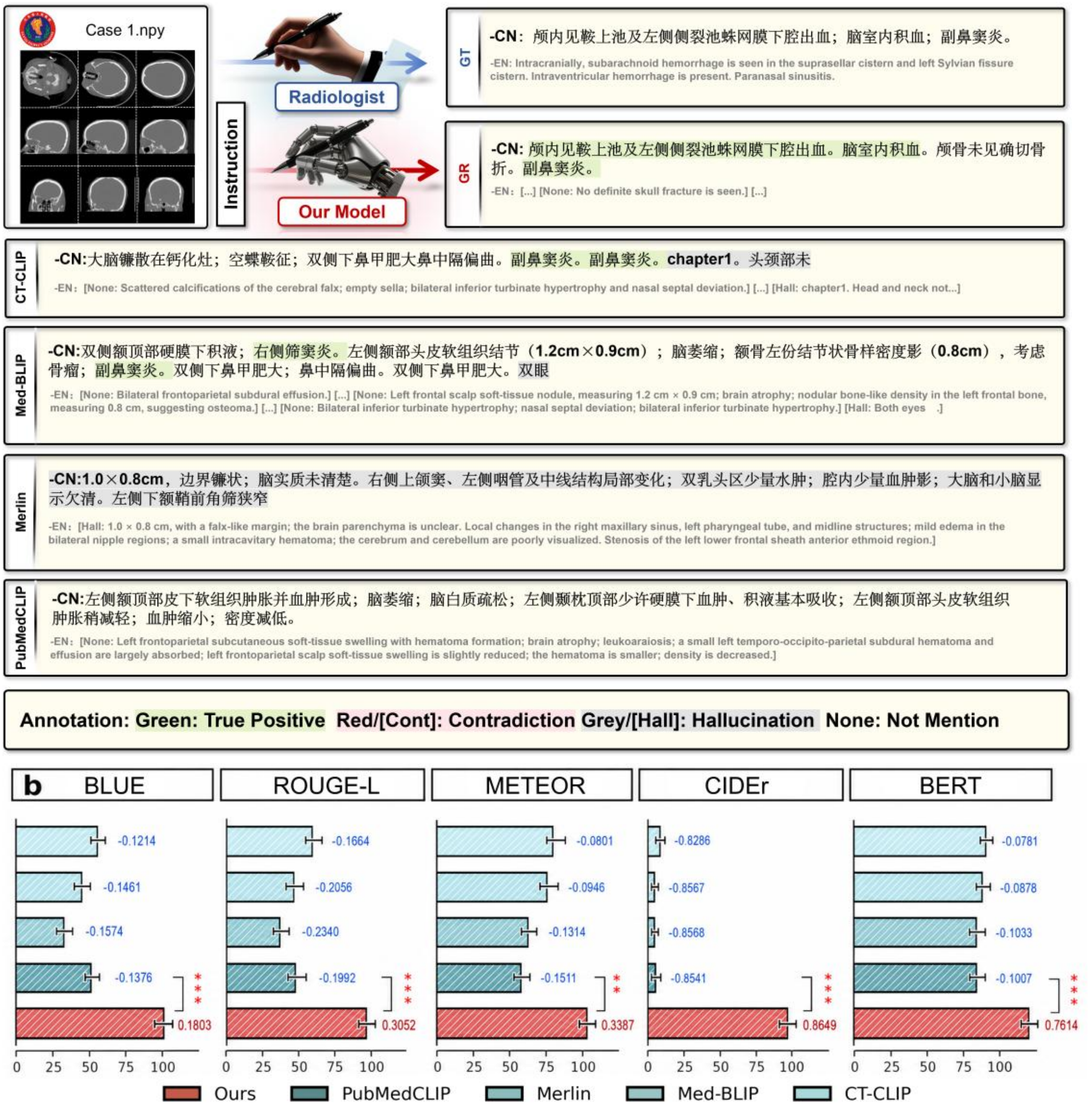

**Supplementary Fig. 2 | Additional report-generation example 1.** Representative head CT report-generation example from the internal or external cohort. a, Selected CT slices, the radiologist-written reference report, the CHIEF-generated report and reports generated by comparator medical vision-language models are shown. Expert annotations highlight correctly described findings, contradictions to the imaging findings, hallucinated content and abnormalities not mentioned. b, Automatic report-generation metrics, including BLEU, ROUGE-L, METEOR, CIDEr and BERTScore, are shown for CHIEF and comparator models.

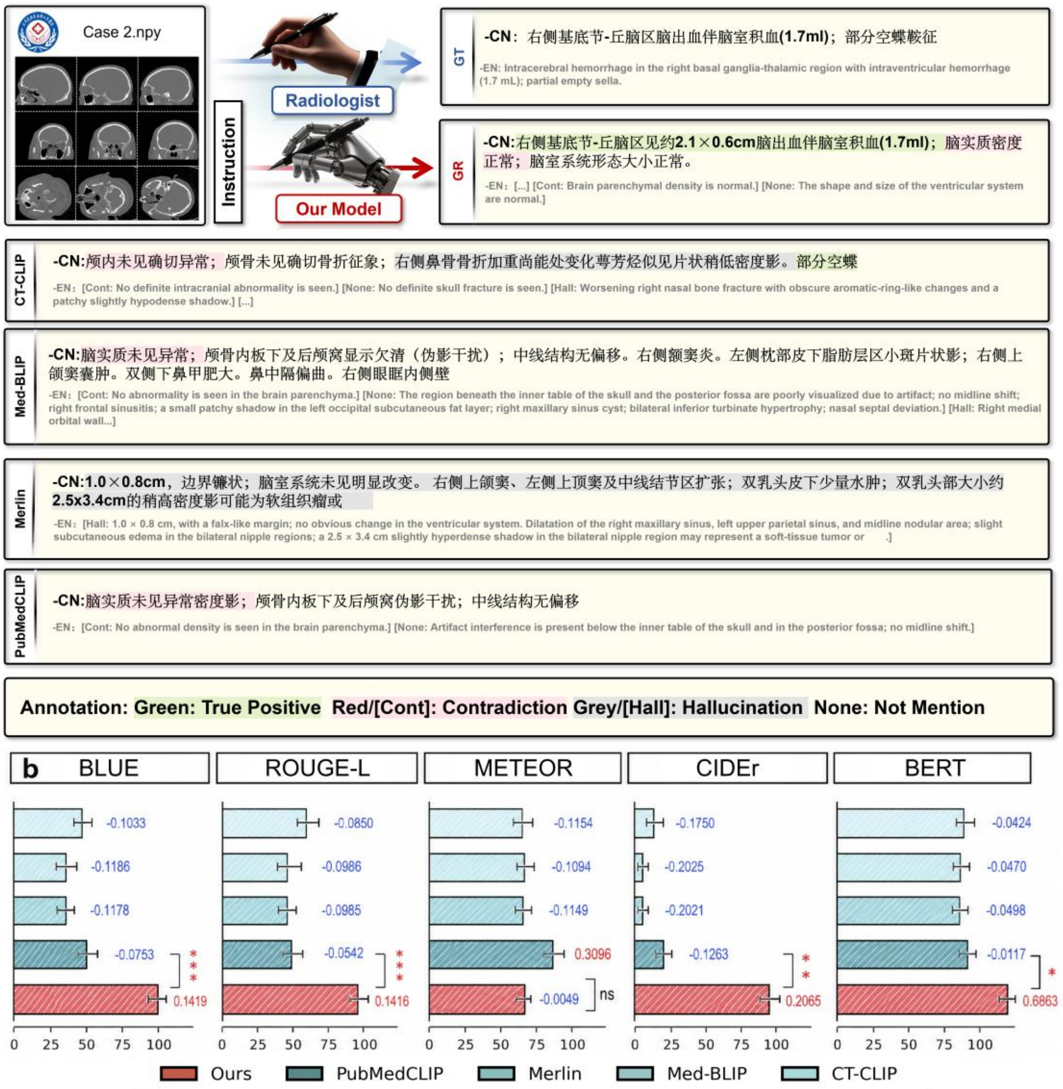

**Supplementary Fig. 3 | Additional report-generation example 2.** Representative head CT report-generation example from the internal or external cohort. a, Selected CT slices, the radiologist-written reference report, the CHIEF-generated report and reports generated by comparator medical vision-language models are shown. Expert annotations highlight correctly described findings, contradictions to the imaging findings, hallucinated content and abnormalities not mentioned. b, Automatic report-generation metrics, including BLEU, ROUGE-L, METEOR, CIDEr and BERTScore, are shown for CHIEF and comparator models.

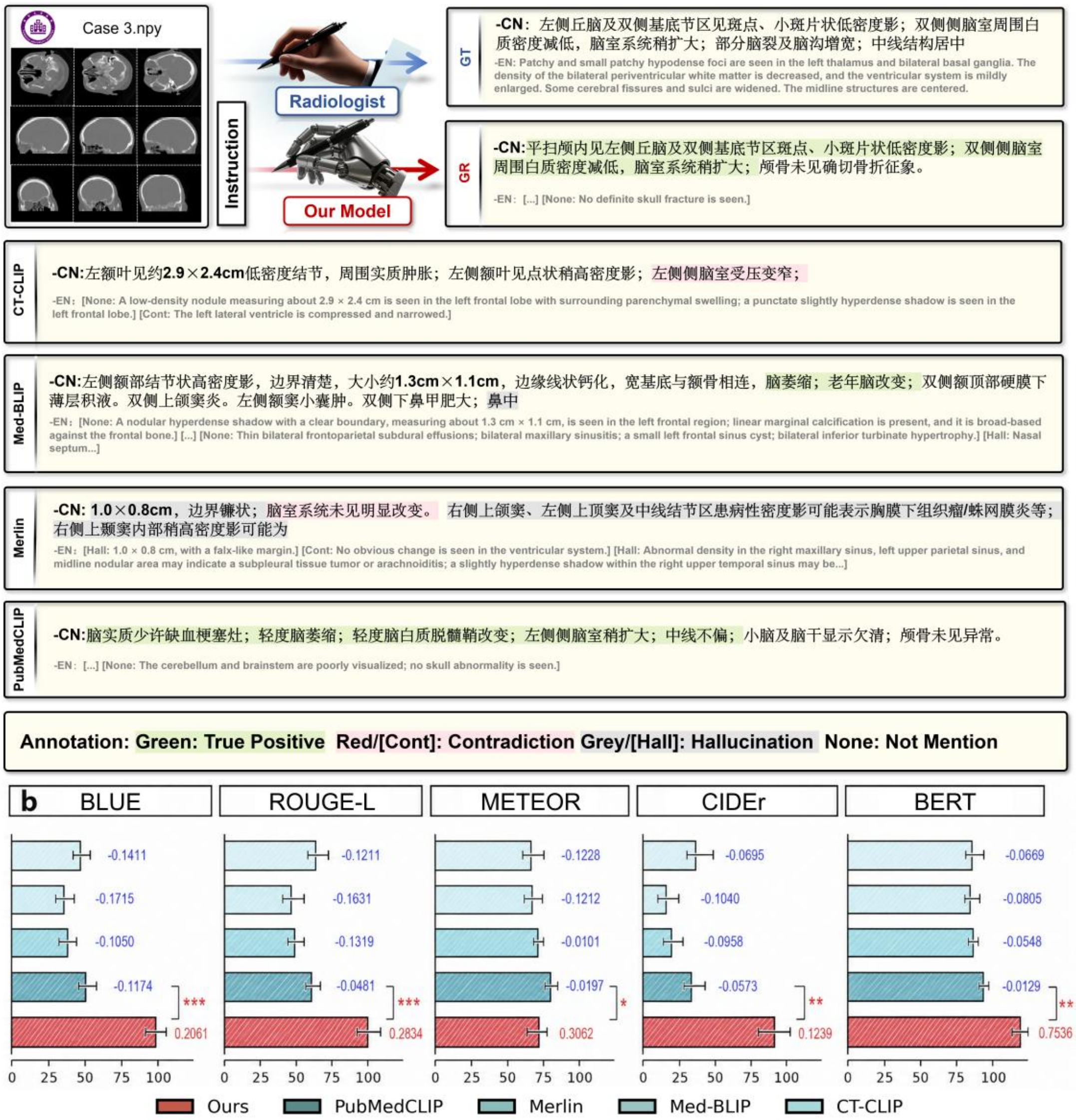

**Supplementary Fig. 4 | Additional report-generation example 3.** Representative head CT report-generation example from the internal or external cohort. a, Selected CT slices, the radiologist-written reference report, the CHIEF-generated report and reports generated by comparator medical vision-language models are shown. Expert annotations highlight correctly described findings, contradictions to the imaging findings, hallucinated content and abnormalities not mentioned. b, Automatic report-generation metrics, including BLEU, ROUGE-L, METEOR, CIDEr and BERTScore, are shown for CHIEF and comparator models.

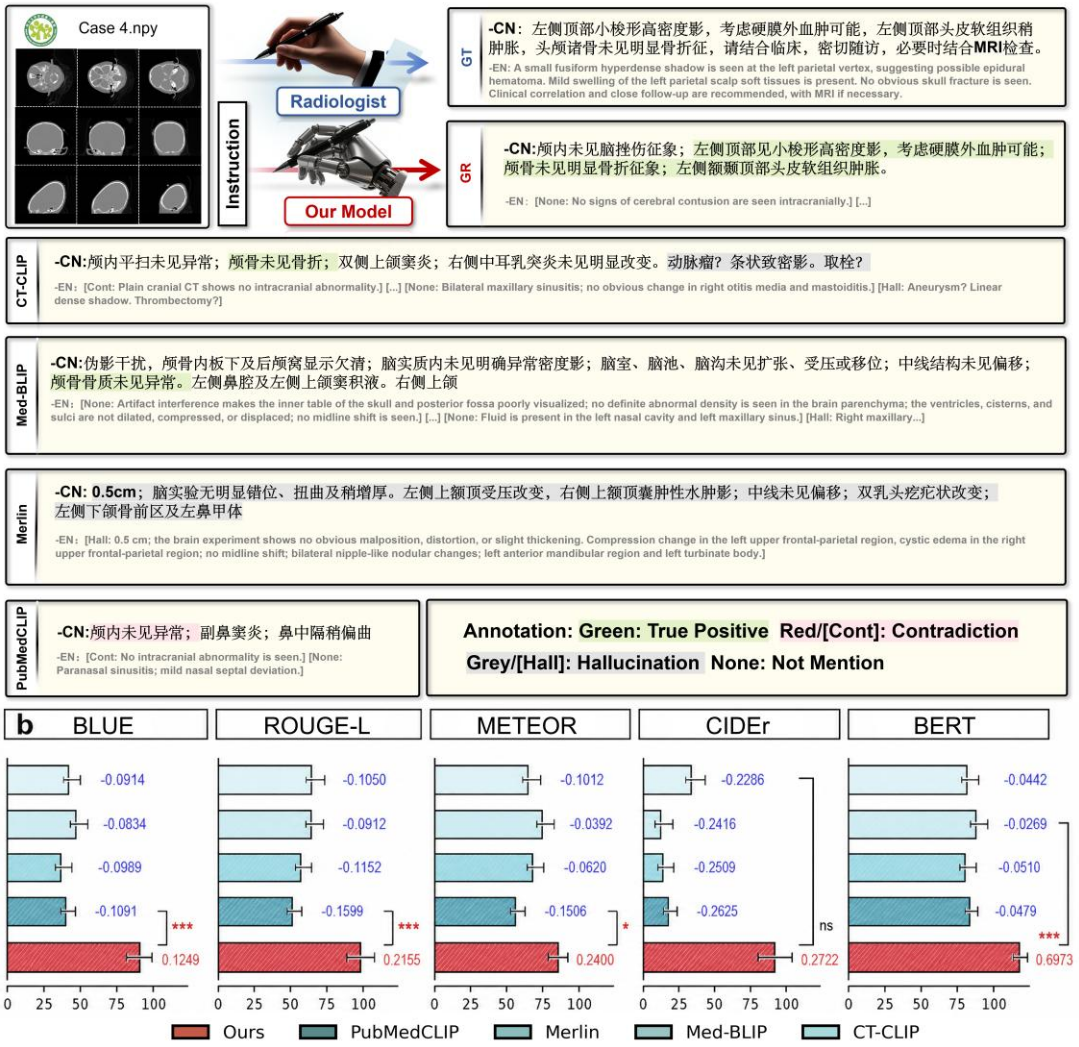

**Supplementary Fig. 5 | Additional report-generation example 4.** Representative head CT report-generation example from the internal or external cohort. a, Selected CT slices, the radiologist-written reference report, the CHIEF-generated report and reports generated by comparator medical vision-language models are shown. Expert annotations highlight correctly described findings, contradictions to the imaging findings, hallucinated content and abnormalities not mentioned. b, Automatic report-generation metrics, including BLEU, ROUGE-L, METEOR, CIDEr and BERTScore, are shown for CHIEF and comparator models.

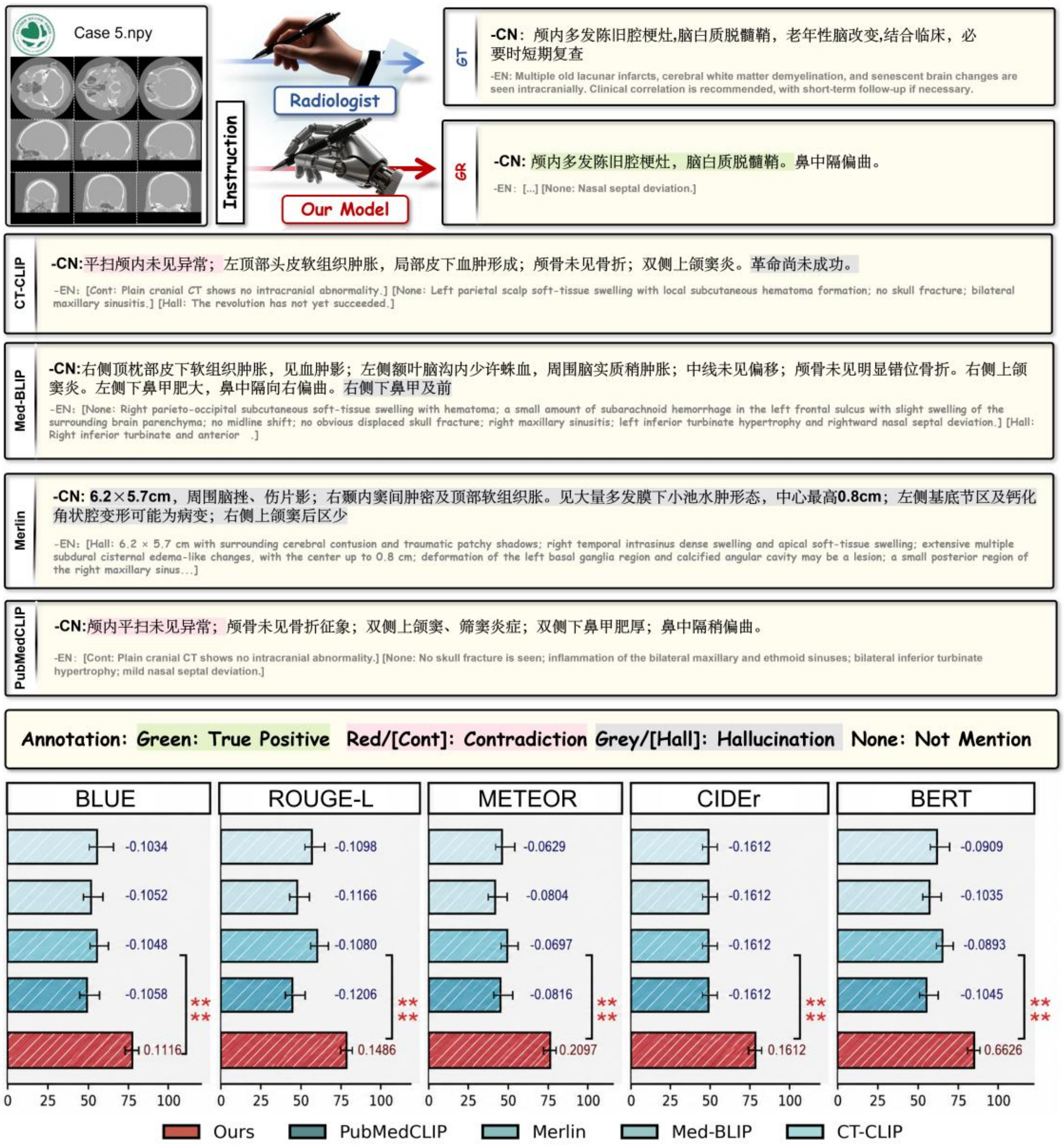

**Supplementary Fig. 6 | Additional report-generation example 5.** Representative head CT report-generation example from the internal or external cohort. a, Selected CT slices, the radiologist-written reference report, the CHIEF-generated report and reports generated by comparator medical vision-language models are shown. Expert annotations highlight correctly described findings, contradictions to the imaging findings, hallucinated content and abnormalities not mentioned. b, Automatic report-generation metrics, including BLEU, ROUGE-L, METEOR, CIDEr and BERTScore, are shown for CHIEF and comparator models.

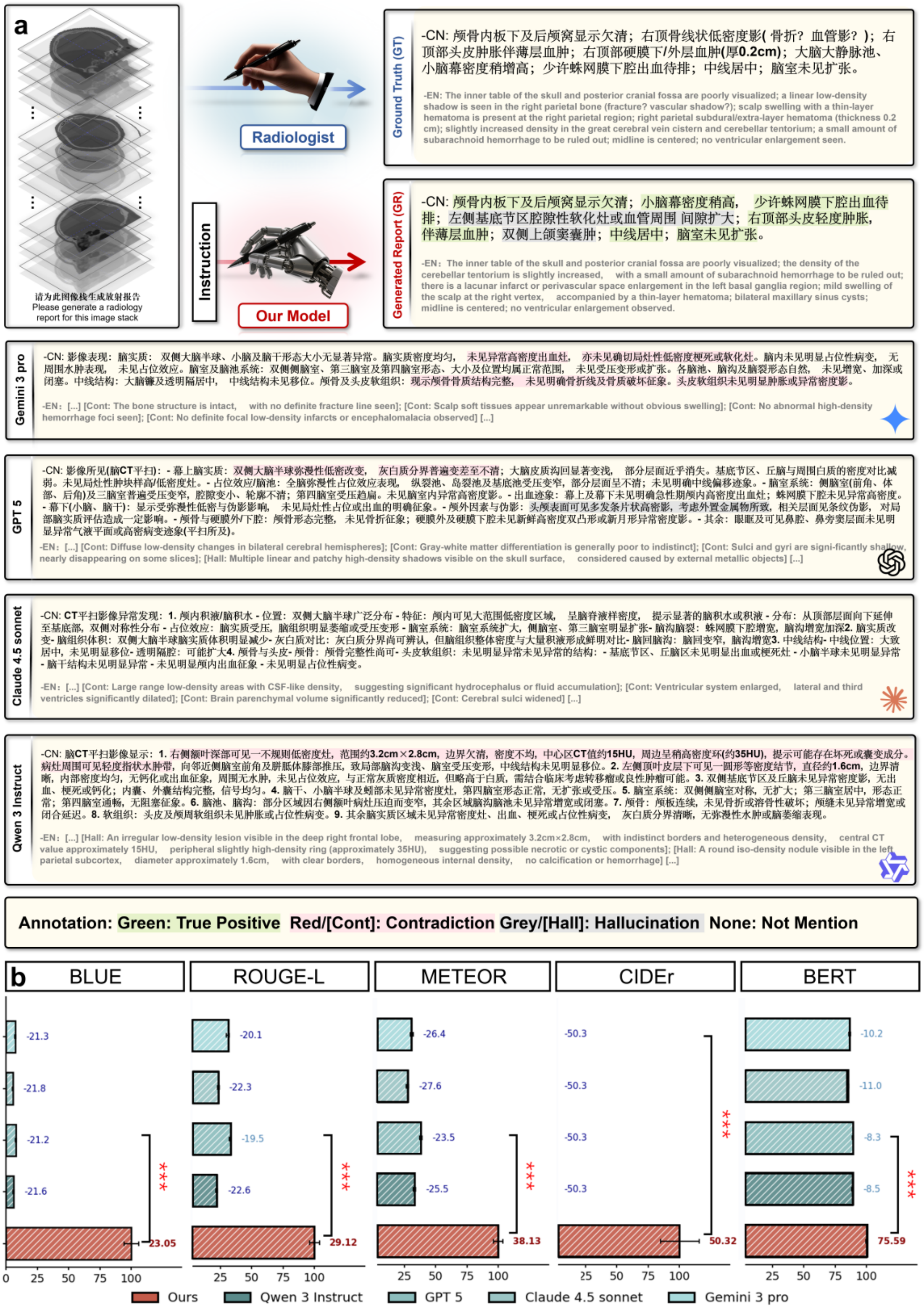
a
请为此图像栈生成放射报告
Please generate a radiology report for this image stack
Radiologist
Instruction
Our Model
Ground Truth (GT)
-CN: 颅骨内板下及后颅窝显示欠清；右顶骨线状低密度影( 骨折？血管影？)；右顶部头皮肿胀伴薄层血肿；右顶部硬膜下/外层血肿(厚0.2cm)；大脑大静脉池、小脑幕密度稍增高；少许蛛网膜下腔出血待排；中线居中；脑室未见扩张。
-EN: The inner table of the skull and posterior cranial fossa are poorly visualized; a linear low-density shadow is seen in the right parietal bone (fracture? vascular shadow?); scalp swelling with a thin-layer hematoma is present at the right parietal region; right parietal subdural/extra-layer hematoma (thickness 0.2 cm); slightly increased density in the great cerebral vein cistern and cerebellar tentorium; a small amount of subarachnoid hemorrhage to be ruled out; midline is centered; no ventricular enlargement seen.
Generated Report (GR)
-CN: 颅骨内板下及后颅窝显示欠清；小脑幕密度稍高，少许蛛网膜下腔出血待排；左侧基底节区腔隙性软化灶或血管周围 间隙扩大；右顶部头皮轻度肿胀，伴薄层血肿；双侧上颌窦囊肿；中线居中；脑室未见扩张。
-EN：The inner table of the skull and posterior cranial fossa are poorly visualized; the density of the cerebellar tentorium is slightly increased, with a small amount of subarachnoid hemorrhage to be ruled out; there is a lacunar infarct or perivascular space enlargement in the left basal ganglia region; mild swelling of the scalp at the right vertex, accompanied by a thin-layer hematoma; bilateral maxillary sinus cysts; midline is centered; no ventricular enlargement observed.
Gemini 3 pro
-EN：[...] [Cont: The bone structure is intact, with no definite fracture line seen]; [Cont: Scalp soft tissues appear unremarkable without obvious swelling]; [Cont: No abnormal high-density hemorrhage foci seen]; [Cont: No definite focal low-density infarcts or encephalomalacia observed] [...]
GPT 5
-EN：[...] [Cont: Diffuse low-density changes in bilateral cerebral hemispheres]; [Cont: Gray-white matter differentiation is generally poor to indistinct]; [Cont: Sulci and gyri are signi-ficantly shallow, nearly disappearing on some slices]; [Hall: Multiple linear and patchy high-density shadows visible on the skull surface, considered caused by external metallic objects] [...]
Claude 4.5 sonnet
-EN：[...] [Cont: Large range low-density areas with CSF-like density, suggesting significant hydrocephalus or fluid accumulation]; [Cont: Ventricular system enlarged, lateral and third ventricles significantly dilated]; [Cont: Brain parenchymal volume significantly reduced]; [Cont: Cerebral sulci widened] [...]
Qwen 3 Instruct
-EN：[...] [Hall: An irregular low-density lesion visible in the deep right frontal lobe, measuring approximately 3.2cm×2.8cm, with indistinct borders and heterogeneous density, central CT value approximately 15HU, peripheral slightly high-density ring (approximately 35HU), suggesting possible necrotic or cystic components]; [Hall: A round iso-density nodule visible in the left parietal subcortex, diameter approximately 1.6cm, with clear borders, homogeneous internal density, no calcification or hemorrhage] [...]
Annotation: Green: True Positive Red/[Cont]: Contradiction Grey/[Hall]: Hallucination None: Not Mention
b
BLUE
-21.3
-21.8
-21.2
-21.6
23.05
ROUGE-L
-20.1
-22.3
-19.5
-22.6
29.12
METEOR
-26.4
-27.6
-23.5
-25.5
38.13
CIDEr
-50.3
-50.3
-50.3
-50.3
50.32
BERT
-10.2
-11.0
-8.3
-8.5
75.59
***
0 25 50 75 100
Ours
Qwen 3 Instruct
GPT 5
Claude 4.5 sonnet
Gemini 3 pro

**Supplementary Fig. 7 | Report generation compared with commercial multimodal large language models.** a, Representative example of report generation for a head CT examination. For each examination, the radiologist-written reference report is shown alongside the report generated by CHIEF and reports generated by commercial large language models, including Gemini 3 Pro, GPT 5, Claude 4.5 Sonnet and Qwen 3 Instruct. Expert reviewers annotated report content as correctly described findings, contradictions to the imaging findings, hallucinated content or abnormalities not mentioned. The examples illustrate differences across models in factual accuracy, hallucination and omission. b, Comparison of CHIEF and commercial large language models using conventional automatic report generation metrics, including BLEU, ROUGE-L, METEOR, CIDEr and BERTScore. These metrics quantify textual overlap or semantic similarity with the reference report, but do not directly assess clinical correctness. The results therefore complement the expert evaluation shown in a. For visualization, all automatic metric values in b are multiplied by 100; values in the text and supplementary tables are reported on the original 0-1 scale.

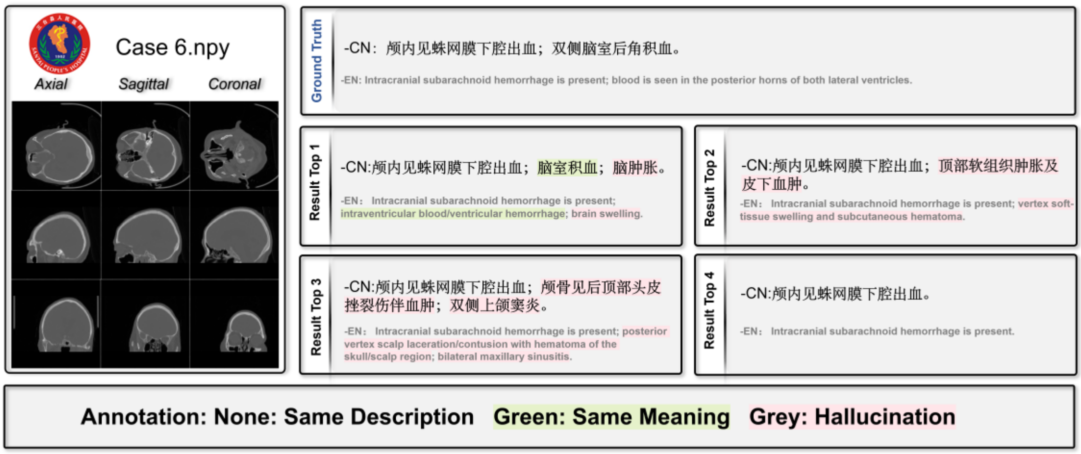

**Supplementary Fig. 8 | Image-to-text retrieval example from the OOD-1 cohort.** A representative query head CT examination from the OOD-1 cohort is shown together with the reference report and the top-ranked retrieved reports. Unhighlighted text denotes the same description as the reference report, green highlighting denotes semantically concordant content, and pink/grey highlighting denotes retrieved statements that are unsupported by or mismatched with the query reference report. This example illustrates retrieval of clinically concordant reports for subarachnoid haemorrhage and intraventricular blood under county-level cross-centre variation.

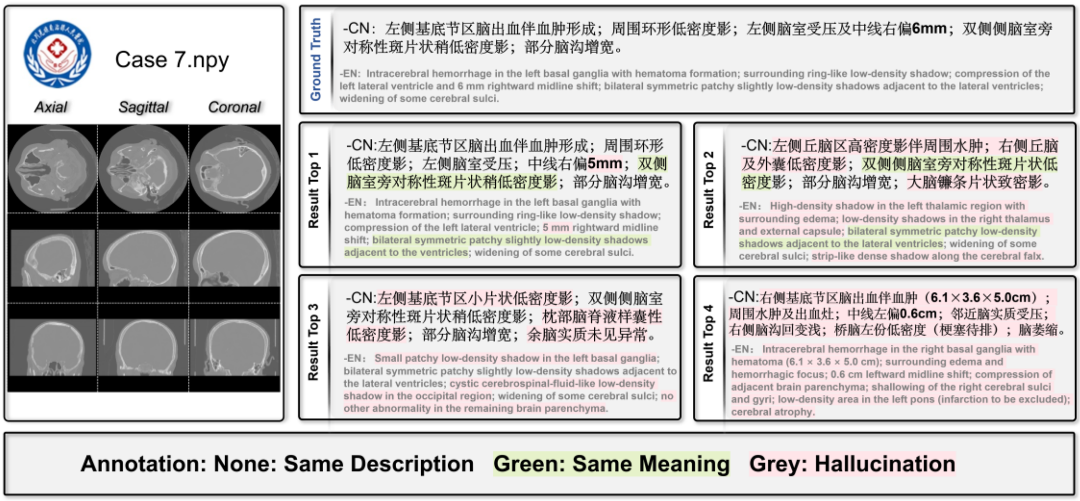

**Supplementary Fig. 9 | Image-to-text retrieval example from the OOD-2 cohort.** A representative query head CT examination from the OOD-2 cohort is shown together with the reference report and the top-ranked retrieved reports. Unhighlighted text denotes the same description as the reference report, green highlighting denotes semantically concordant content, and pink/grey highlighting denotes retrieved statements that are unsupported by or mismatched with the query reference report. This example illustrates retrieval of clinically similar reports for basal ganglia haemorrhage, ventricular compression and midline shift under external institutional variation.

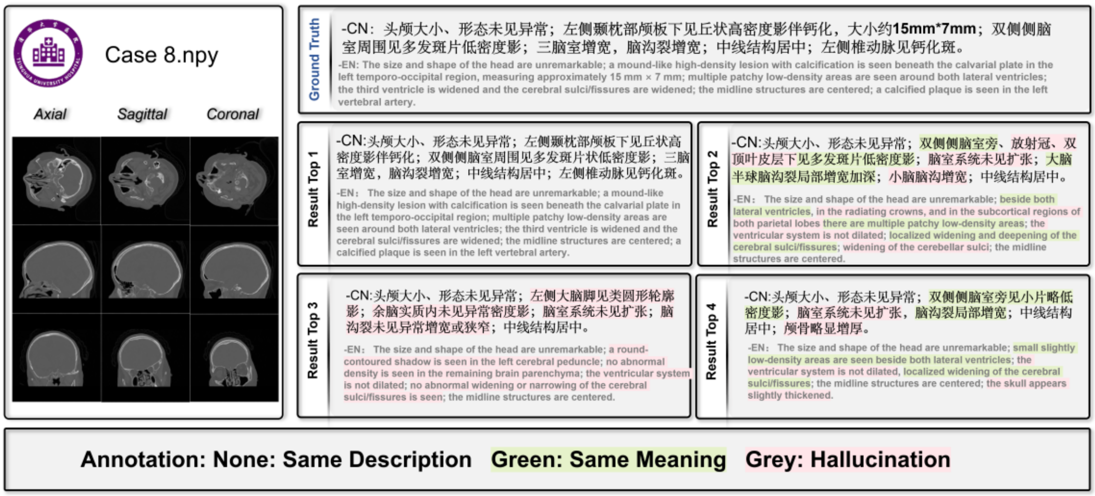

**Supplementary Fig. 10 | Image-to-text retrieval example from the OOD-3 cohort.** A representative query head CT examination from the OOD-3 cohort is shown together with the reference report and the top-ranked retrieved reports. Unhighlighted text denotes the same description as the reference report, green highlighting denotes semantically concordant content, and pink/grey highlighting denotes retrieved statements that are unsupported by or mismatched with the query reference report. This example illustrates retrieval of semantically similar reports for chronic or incidental findings, including calcification, periventricular low-density change and ventricular or sulcal enlargement.

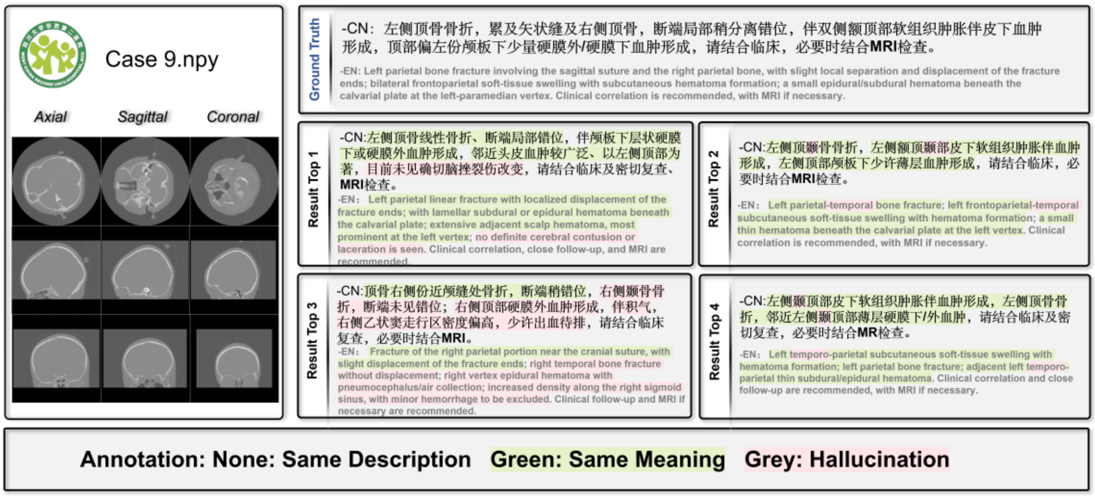

**Supplementary Fig. 11 | Image-to-text retrieval example from the OOD-4 cohort.** A representative query head CT examination from the OOD-4 cohort is shown together with the reference report and the top-ranked retrieved reports. Unhighlighted text denotes the same description as the reference report, green highlighting denotes semantically concordant content, and pink/grey highlighting denotes retrieved statements that are unsupported by or mismatched with the query reference report. This example illustrates retrieval of clinically similar reports for skull fracture, scalp soft-tissue swelling and small extra-axial haematoma in a paediatric-specialty external cohort.

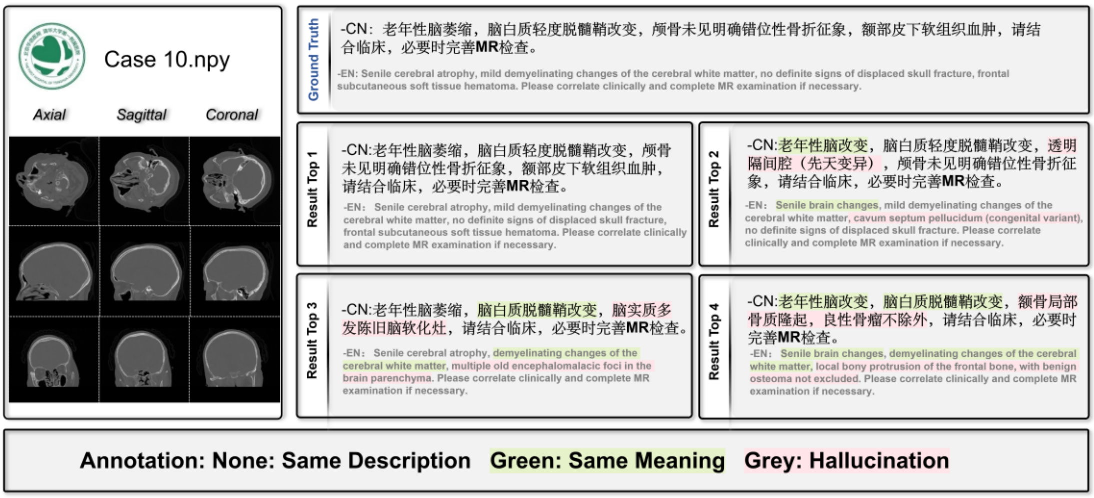

**Supplementary Fig. 12 | Image-to-text retrieval example from the OOD-5 cohort.** A representative query head CT examination from the OOD-5 cohort is shown together with the reference report and the top-ranked retrieved reports. Unhighlighted text denotes the same description as the reference report, green highlighting denotes semantically concordant content, and pink/grey highlighting denotes retrieved statements that are unsupported by or mismatched with the query reference report. This example illustrates retrieval of clinically similar reports for chronic brain changes, demyelinating white-matter change and frontal subcutaneous soft-tissue haematoma.

**Definition and mapping of abnormality labels used for zero-shot detection**

低密度(Low density)， 高密度(High density)， 占位(Space-occupying lesion)， 脑膜强化(Meningeal enhancement)，
水肿带(Edema zone)， 钙化(Calcification)， 囊状(Cystic)， 占位效应(Mass effect)， 出血(Hemorrhage)， 梗死(Infarction)，
腔隙性(Lacunar)， 萎缩(Atrophy)， 硬化(Sclerosis)， 移位(Displacement)， 脑室扩大(Ventricular enlargement)，
脑室受压(Ventricular compression)， 变窄(Narrowing)， 脑水肿(Cerebral edema)， 脑肿胀(Brain swelling)，
鼻甲肥大(Turbinate hypertrophy)， 中隔偏曲(Nasal septal deviation)， 鼻窦黏膜增厚(Sinus mucosal thickening)，
鼻腔积液(Nasal cavity fluid accumulation)， 急性脑梗死(Acute cerebral infarction)， 硬膜下血肿(Subdural hematoma)，
脑转移瘤(Brain metastasis)， 颅内动脉瘤(Intracranial aneurysm)， 蛛网膜下腔出血(Subarachnoid hemorrhage)，
真菌性上颌窦炎(Fungal maxillary sinusitis)， 脑膜瘤(Meningioma)， 脱髓鞘病变(Demyelinating lesion)，
颅骨凹陷性骨折(Depressed skull fracture)， 脑积水(Hydrocephalus)， 脑积液(Intracranial fluid collection)，
脑积血(Intracranial blood collection)， 脑萎缩(Cerebral atrophy)， 脑动脉硬化(Cerebral arteriosclerosis)，
轴索损伤(Axonal injury)， 筛窦炎(Ethmoid sinusitis)， 蝶窦炎(Sphenoid sinusitis)， 额窦炎(Frontal sinusitis)，
中耳乳突炎(Otitis media with mastoiditis)， 乳突积液(Mastoid effusion)， 胆脂瘤(Cholesteatoma)，
脑软化灶(Encephalomalacic focus)， 脑白质疏松(Leukoaraiosis)， 蛛网膜囊肿(Arachnoid cyst)， 大枕大池(Mega cisterna magna)， 眶脂体疝(Orbital fat herniation)， 中线移位(Midline shift)， 脑疝(Brain herniation)， 脑干受压(Brainstem compression)， 颅内压升高(Increased intracranial pressure)， 脑沟消失(Effacement of cerebral sulci)， 脑实质出血(Intraparenchymal hemorrhage)， 脑脊液漏(Cerebrospinal fluid leak)， 颅内积气(Pneumocephalus)， 脑镰下疝(Subfalcine herniation)， 脑室扩大伴受压(Ventricular enlargement with compression)， 脑膜脑膨出(Meningoencephalocele)，
脑干出血(Brainstem hemorrhage)， 颅内血肿(Intracranial hematoma)， 脑挫裂伤(Cerebral contusion and laceration)，
脑脊液外渗(Cerebrospinal fluid extravasation)， 脑室扩张(Ventricular dilatation)， 松果体区囊肿(Pineal region cyst)

**Supplementary Fig. 13 | Abnormality label bank used for internal zero-shot abnormality detection.** Chinese abnormality terms and their English mappings used for question-template-based zero-shot abnormality detection are listed. These labels were derived from the internal matched CT-report cohort and were used to construct abnormality-specific prompts for estimating yes/no probabilities from the image-conditioned language model. The label bank supports both overall abnormality-versus-negative evaluation and fine-grained label-level abnormality detection in the internal matched cohort.

**Negation expressions**

无(Absent)，未见(Not seen)，未发现(Not detected)，未显示(Not demonstrated)，未提示(Not suggestive of)，未见明显(No obvious evidence of)，无明显(No obvious)，未见明确(No definite evidence of)，未见确切(No definite evidence of)，排除(Rule out)，除外(Exclude)，不支持(Not supportive of)，阴性(Negative)，正常(Normal)，未见异常(No abnormality detected)，未见明显异常(No obvious abnormality detected)

**Supplementary Fig. 14 | Negation expressions used for report-derived abnormality-label construction.** Negation expressions were used to identify findings described as absent, not detected, not demonstrated, normal or unsupported when deriving abnormality labels from radiology reports in the internal matched cohort. These rules were applied before internal zero-shot abnormality detection to reduce false-positive report-derived labels caused by negated or normal statements.

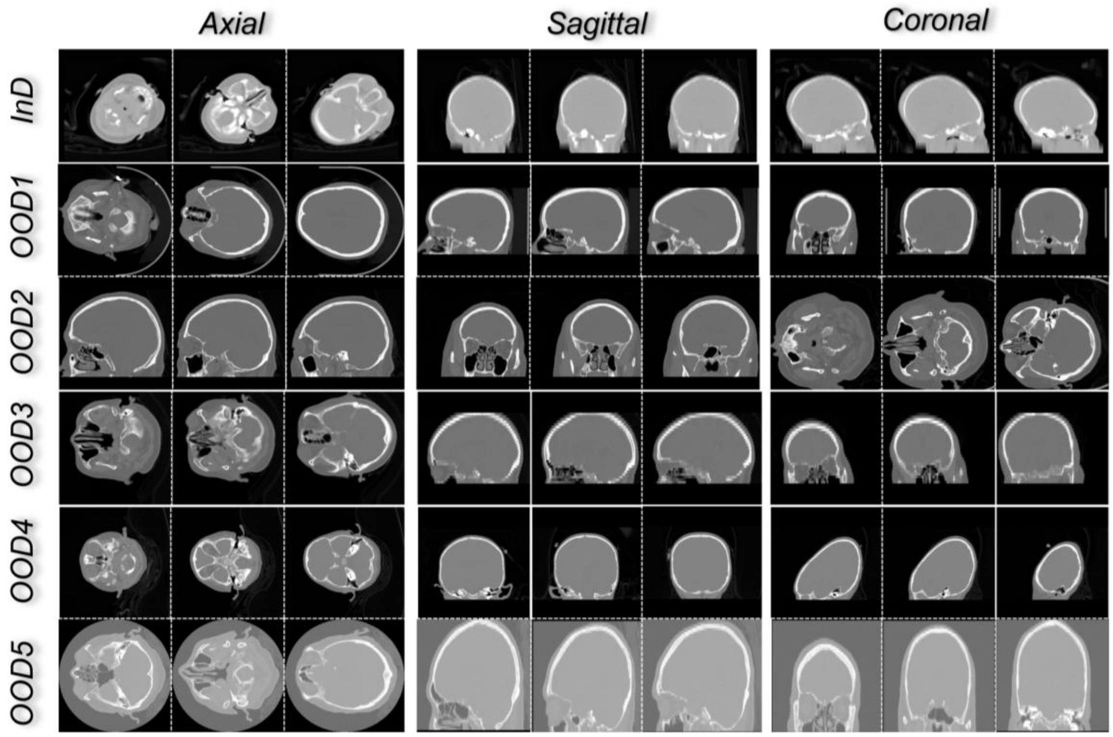

**Supplementary Fig. 15 | Representative CT examinations from the internal and five out-of-distribution cohorts.** Representative examinations from the internal cohort and each external institution are shown using selected axial, sagittal and coronal views. These examples illustrate cross-centre variation in acquisition appearance, patient population and anatomical presentation across the in-distribution and out-of-distribution cohorts.
